# Assessing the performance of ChatGPT in answering questions regarding cirrhosis and hepatocellular carcinoma

**DOI:** 10.1101/2023.02.06.23285449

**Authors:** Yee Hui Yeo, Jamil S. Samaan, Wee Han Ng, Peng-Sheng Ting, Hirsh Trivedi, Aarshi Vipani, Walid Ayoub, Ju Dong Yang, Omer Liran, Brennan Spiegel, Alexander Kuo

## Abstract

**Background:** Patients with cirrhosis and hepatocellular carcinoma (HCC) require extensive and personalized care to improve outcomes. ChatGPT (Generative Pre-trained Transformer), a natural language processing model, holds potential to provide professional yet patient-friendly support.

**Aim:** Examining the accuracy and reproducibility of ChatGPT in answering questions regarding knowledge, management, and emotional support for cirrhosis and HCC.

**Method:** ChatGPT’s responses to 164 questions were independently graded by two transplant hepatologists and resolved by a third reviewer. The performance of ChatGPT was compared to physicians or trainees in two validated questionnaires. ChatGPT’s knowledge on cirrhosis care was tested using 26 quality measures of cirrhosis management. Finally, its emotional support capacity was tested.

**Results:** ChatGPT regurgitated extensive knowledge of cirrhosis and HCC, but for questions with correct responses, only a small proportion was labelled as comprehensive. The performance was better in basic knowledge, lifestyle, and treatment than in the domains of diagnosis and preventive medicine. For the quality measures, the model answered 76.9% of questions correctly but failed to specify decision-making cut-offs and treatment durations. Compared to physicians/trainees, ChatGPT lacked knowledge of regional guidelines variations, such as HCC screening criteria. However, it provided practical and multifaceted advice to patients and caregivers regarding the next steps and adjusting to a new diagnosis.

**Conclusion:** In summary, we analyzed the areas of robustness and limitations of ChatGPT’s responses on the management of cirrhosis and HCC and relevant emotional support. ChatGPT may have a role as an adjunct informational tool for patients and physicians to improve outcomes.

**Conflict of Interest Disclosures:** None declared.

**Funding/Support:** None

**Ethics Approval:** Since all responses from ChatGPT were publicly available, approval from the institutional review board was not sought.

Guarantors of article:

Dr. Kuo and Spiegel

**Author Contributions:** Concept and design: Yeo, Samaan, Spiegel, Kuo

Acquisition of data: Yeo, Samaan, Ng, Vipani

Data review: Ting, Trivedi, Kuo

Statistical analysis: Yeo

Drafting of the manuscript: Yeo, Samaan, Ng

Critical revision of the manuscript: All authors

## Introduction

Cirrhosis is an end-stage liver disease that continues to be a rising global health threat. In 2017, the estimated global age-standardized prevalence of compensated and decompensated cirrhosis were 1395.0 and 132.5 per 100,000 population, respectively, with 2.4% of worldwide deaths related to cirrhosis.^1^ Cirrhosis is a major risk factor for hepatocellular carcinoma (HCC), which has also demonstrated increased incidence and mortality in recent decades.^2,3^ The incidence rate of HCC is projected to increase by 55% by 2040.^4^ With the substantial clinical and economic burden on patients, caregivers, and society, effective management of cirrhosis and HCC is pivotal.^5-7^

Optimal management of cirrhosis, particularly in those with decompensated cirrhosis, may be challenging. Complications such as ascites, hepatic encephalopathy, variceal bleeding, and malnutrition, are clinically complex and require significant medical attention.^8^ For instance, variceal bleeding poses a six-week mortality rate of more than 20%,^9^ and requires acute interventions as well as adherence to preventative measures such as medications, re-interventions, and regular outpatient follow-up.^10^ Similarly, HCC also requires proactive management and coordination of complex care for patients, especially when comorbid with cirrhosis.^11,12^ The complexity of care required for this patient population makes patient empowerment with knowledge about their disease crucial for optimal outcomes.

Patients who suffer from cirrhosis and HCC, as well as their caregivers, often have unmet needs and insufficient knowledge about managing and preventing complications of their disease. Prior studies have reported low health literacy in patients with cirrhosis and HCC.^13,14^ Although the internet may serve as a source of health information for patients, the complexity of the primary literature, and potential misinformation may result in confusion rather than clarity. A survey of 401 patients with chronic liver disease showed that only 15.7% of patients were aware of the safe dose of acetaminophen, and less than 20% reported awareness that medications such as Norco, Vicodin, and Percocet contain acetaminophen.^15^ Another study showed that online resources available to patients through health platforms and hepatology centers were lengthy and complex, highlighting the limited availability of easy-to-understand information for this patient population.^16^

ChatGPT (Generative Pre-trained Transformer) is a natural language processing (NLP) model developed by OpenAI.^17^ It generates human-like text for use in chatbot conversations.^17,18^ It has seen several potential applications in the medical field such as the ability to answer medical student examination questions,^19^ and write basic medical reports.^20^ However, there are reports raising concerns about the ability of ChatGPT to comprehend questions and the lack of in depth response.^21^ To date, there is no current literature examining the ability of ChatGPT to answer clinically oriented disease-specific questions correctly and comprehensively. In this study, we aim to assess the accuracy, completeness, and reproducibility of ChatGPT’s responses to frequently asked questions about the management and care of patients with cirrhosis and HCC. To further examine the knowledge base and problem-solving skills of ChatGPT, we also examined its performance compared to physicians and trainees on published knowledge questionnaires as reported in the individual studies.

## Methods

### Data source

First, we obtained frequently asked questions with cirrhosis or HCC posted by well-regarded professional societies and institutions. To enhance the inclusiveness and representation of patients and caregivers, questions about cirrhosis and HCC knowledge and management were also collected from posts in patient support groups on Facebook. Exclusion criteria included questions with similar meaning, questions with vague meaning (e.g., how does HCC affect my body?), questions that may vary from person to person (e.g., what is the chance that my cancer will come back?), and non-medical questions about the condition (e.g., what support groups are available?) (**Figure 1**). The wording and grammar of some questions was modified to ensure accuracy of the questions. A total of 73 and 91 questions were selected for HCC and cirrhosis, respectively.

**Figure 1.**
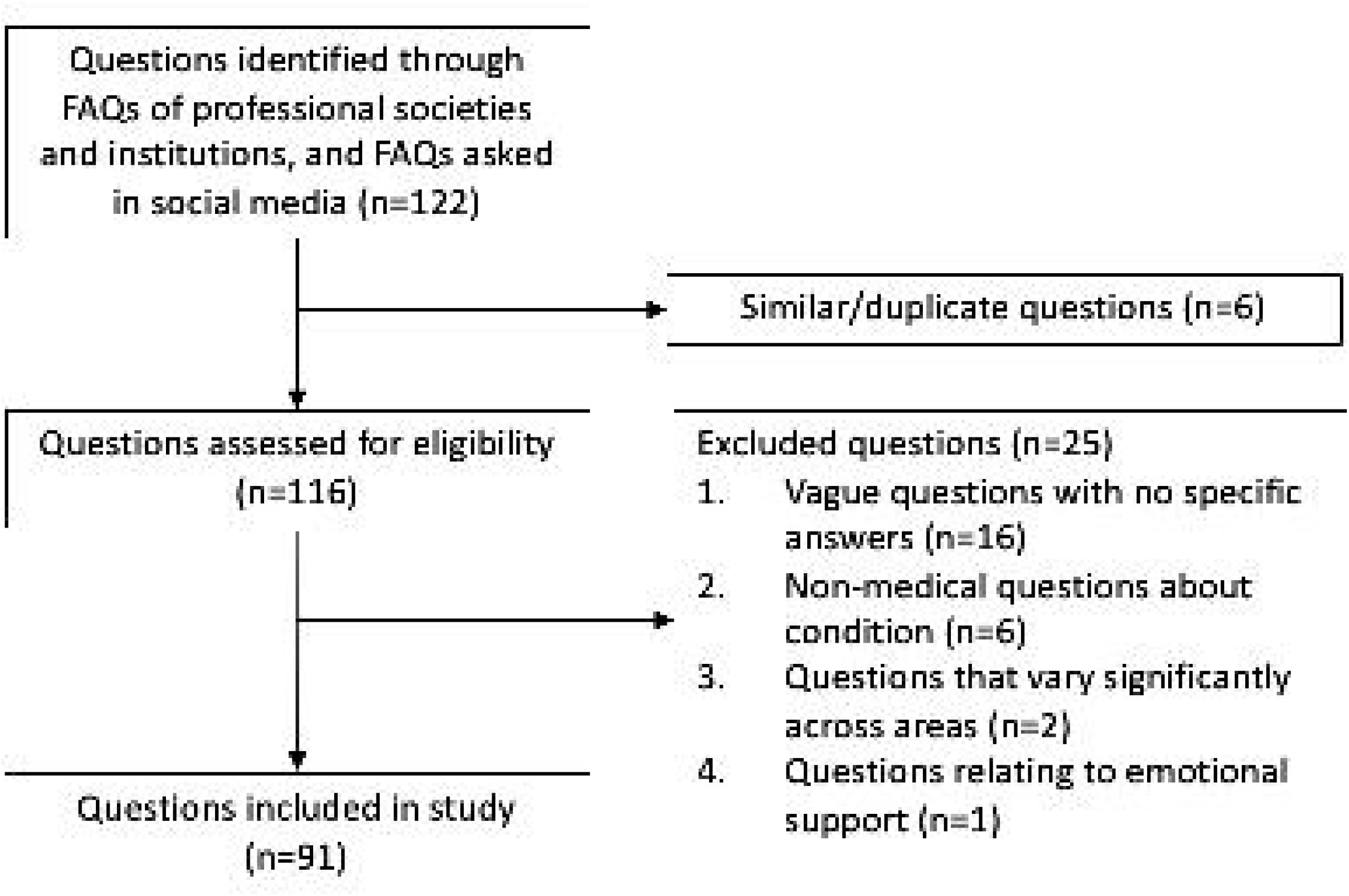
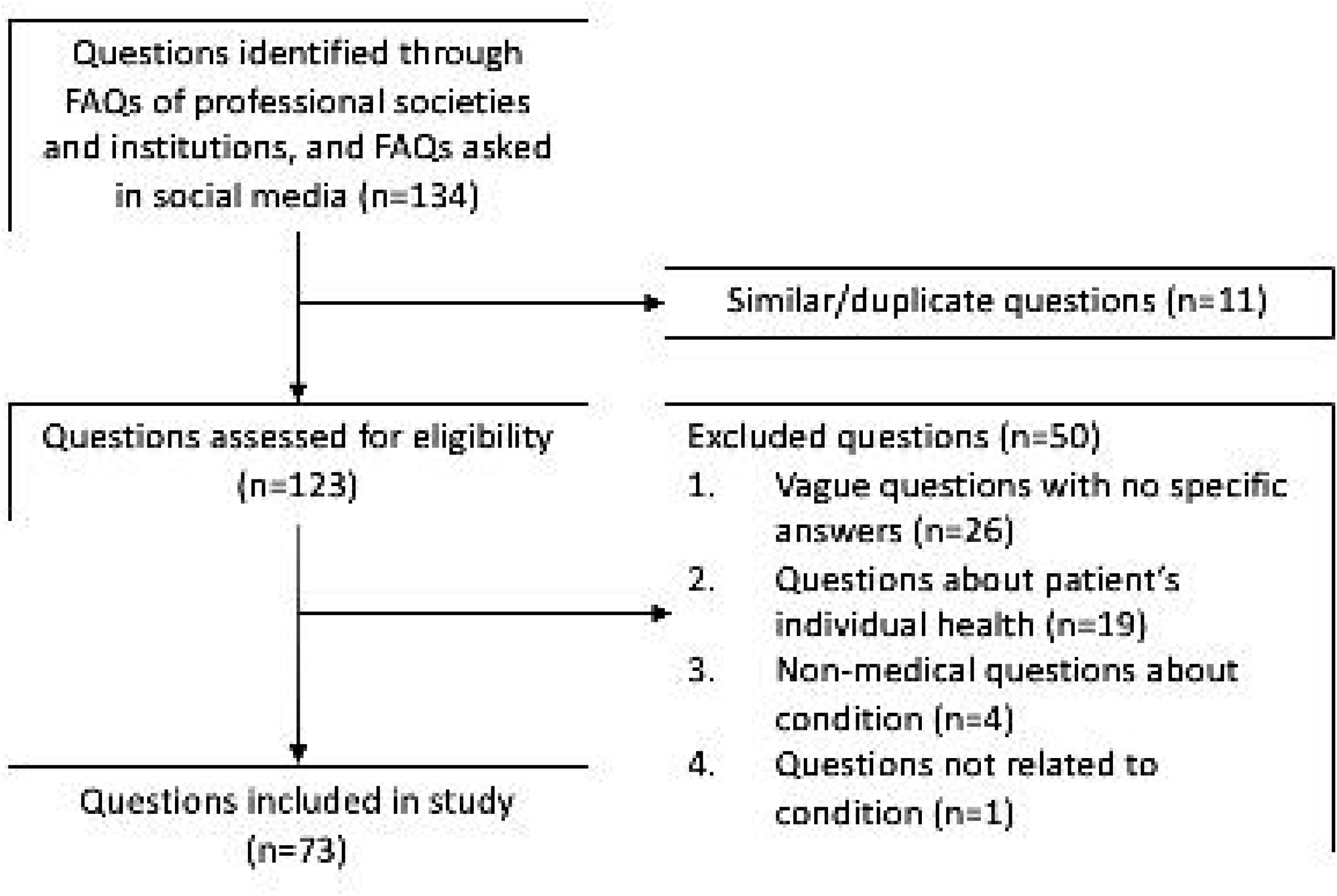
Flow chart of frequently asked question selection. (A) Cirrhosis (B) hepatocellular carcinoma. Frequently asked questions about the knowledge and management of cirrhosis or HCC were collected from patient support groups on Facebook and well-regarded professional societies and institutions.

### ChatGPT and response generation

ChatGPT is a natural language processing model (NLM), with the first research preview prototype released in November 2022.^17^ It is a variant of the GPT-3.5 LLM (Large Language Model) that is pre-trained on a large dataset of text obtained from online sources, including websites, books, and articles leading up to 2021. The exact amount of information has not been disclosed. ChatGPT was then trained to respond by incorporating feedback and correction from human input, allowing it to generate more coherent and contextually appropriate responses.^22^ This process is known as Reinforcement Learning from Human Feedback or Reinforcement Learning from Human Preference (RLHF/RLHP). Users can input any prompt, and ChatGPT will generate a response based on the information stored in its database. To prevent abuse, the developers of ChatGPT have multiple safeguards in place, such as its inability to generate any derogatory or dangerous responses.

Questions were entered into the ChatGPT Dec 15 version. Each question was entered as a separate, independent prompt using the “New Chat” function. Each question was entered into ChatGPT twice, and both responses were recorded to examine the reproducibility of ChatGPT’s responses.

### Grading

Review and grading of each response was done independently by two board certified/eligible transplant hepatologist reviewers. The accuracy of each response was graded using the following grading system: 1. Comprehensive, 2. Correct but inadequate, 3. Mixed with correct and incorrect/outdated data, and 4. Completely incorrect. Reproducibility was independently determined by each reviewer by assessing the similarity of the two responses to each individual question. For questions with different responses (e.g., contradictory information, varying levels of detail, etc.), both versions were separately evaluated by reviewers. Discrepancies in grading and assessment of reproducibility among the two reviewers were independently reviewed and resolved by a blinded third board certified senior hepatologist with more than twenty years of clinical experience in transplant hepatology.

### Performance of ChatGPT on published knowledge questionnaires

To further examine the ChatGPT’s knowledge base and clinical problem-solving, we compiled published questionnaires which tested physicians’ or trainees’ knowledge regarding HCC screening and surveillance in patients with cirrhosis or chronic hepatitis B infection. Each question was posed twice to ChatGPT, and the overall proportion of correct answers was calculated along with examining the provided explanations. We graded the responses according to the American Association for the Study of Liver Diseases (AASLD) guidelines. We compared the proportion of correct responses to that of interviewed physicians or trainees reported in each study, as well as commented on the incorrect responses.

### Understanding the quality measures in cirrhosis

We investigated the ability of ChatGPT to answer and explain standard quality measures in cirrhosis by formulating the 26 quality measures recommended by the practice metrics committee of the AASLD into questions. Using these practice metrics as the standard answers, we tested the performance of ChatGPT in the management of patients with cirrhosis.

### Response to questions requiring emotional support

We observed a number of queries posted on the patient support groups seeking emotional support in comparison to medical information related to each disease. To fill this unmet need, we assessed ChatGPT’s potential as a psychological support system for patients and their caregivers. As there was no established standard for the answers, two physician authors were involved in the process of appraising the responses and evaluating the model’s efficacy in providing emotional support.

### Statistical analysis

The proportions of each aforementioned grade among responses for each domain of cirrhosis and HCC were calculated and reported as percentages.

To assess the reproducibility of ChatGPT’s responses, a review was conducted to examine the similarity of the two responses generated by the model for each question posed. Reproducibility was defined as the consistency of two responses with comparable grading categorizations. For those responses that differed in content for an individual question, such as having different cut-off values or meanings, both were graded. The grading categorization was divided into two groups: grades 1 and 2 vs. grades 3 and 4. If the two responses were categorized into different groups, they were defined as being significantly different from each other. The analyses were calculated using Microsoft Excel (version 16.69.1).

## Results

### Frequently asked questions about cirrhosis

ChatGPT demonstrated high levels of accuracy in responding to 91 questions from a variety of domains (**Table 1**). The proportion of responses graded as comprehensive or correct but inadequate was 75% or higher for “basic knowledge,” “treatment,” “lifestyle,” and “others.” However, this proportion was 66.7% in the “diagnosis” domain and 50% in the “preventive medicine” domain (**Supplemental Table 1)**. No responses from ChatGPT were graded as completely incorrect. Overall, there were 19% of the questions graded differently (comprehensive or correct but inadequate versus mixed with correct and incorrect/outdated data or completely incorrect) by two reviewers and resolved by the third reviewer.

**Table 1.** Grade of responses by the ChatGPT language model to questions related to cirrhosis and hepatocellular carcinoma (HCC)

|  | **Cirrhosis** | **HCC** |
| --- | --- | --- |
| **Basic Knowledge (N= 36, 16)** |  |  |
| 1. Comprehensive | 47.2% | 50.0% |
| 2. Correct but inadequate | 30.6% | 25.0% |
| 3. Mixed with correct and incorrect/outdated data | 22.2% | 25.0% |
| 4. Completely incorrect | 0.0% | 0.0% |
| **Diagnosis (N= 3, 6)** |  |  |
| 1. Comprehensive | 0.0% | 0.0% |
| 2. Correct but inadequate | 66.7% | 16.7% |
| 3. Mixed with correct and incorrect/outdated data | 33.3% | 50.0% |
| 4. Completely incorrect | 0.0% | 33.3% |
| **Treatment (N= 16, 30)** |  |  |
| 1. Comprehensive | 31.2% | 40.0% |
| 2. Correct but inadequate | 43.8% | 36.7% |
| 3. Mixed with correct and incorrect/outdated data | 25.0% | 16.7% |
| 4. Completely incorrect | 0.0% | 6.7% |
| **Lifestyle (N= 22, 8)** |  |  |
| 1. Comprehensive | 50.0% | 50.0% |
| 2. Correct but inadequate | 31.8% | 37.5% |
| 3. Mixed with correct and incorrect/outdated data | 18.1% | 0.0% |
| 4. Completely incorrect | 0.0% | 12.5% |
| **Preventive medicine (N= 4, 0)** |  |  |
| 1. Comprehensive | 25.0% | NA |
| 2. Correct but inadequate | 25.0% | NA |
| 3. Mixed with correct and incorrect/outdated data | 50.0% | NA |
| 4. Completely incorrect | 0.0% | NA |
| **Others (N= 10, 13)** |  |  |
| 1. Comprehensive | 90.0% | 46.2% |
| 2. Correct but inadequate | 10.0% | 38.5% |
| 3. Mixed with correct and incorrect/outdated data | 0.0% | 15.4% |
| 4. Completely incorrect | 0.0% | 0.0% |

The model demonstrated an ability to provide comprehensive responses to basic knowledge- and lifestyle-related questions. It provided detailed explanations of the symptoms, etiology, and prognosis of compensated and decompensated cirrhosis, as well as the risk factors and lifestyle modifications that may impact outcomes. Although the model was able to answer questions in areas such as diagnosis, treatment, and preventive medicine correctly, the majority were graded as correct but inadequate.

The proportion of responses that were “mixed with correct and incorrect/outdated data” was 22.2%, 33.3%, 25.0%, 18.1%, and 50.0% in the “basic knowledge,” “diagnosis,” “treatment,” “lifestyle,” and “preventive medicine” domains, respectively. Reproducibility was high, with 90.48% of all questions producing two similar responses with similar grading. Similarly, the reproducibility of responses was high among all subgroups (**Table 2**).

**Table 2.** Percentage of questions with significantly different responses and difference in grading between the two responses.

|  | **Reproducibility** |  |
| --- | --- | --- |
|  | **Cirrhosis** | **HCC** |
| **Basic Knowledge (N= 36, 16)** | 2.78% | 0.00% |
| **Diagnosis (N= 3, 6)** | 0.00% | 0.00% |
| **Treatment (N= 16, 30)** | 6.25% | 0.00% |
| **Lifestyle (N= 22, 8)** | 4.54% | 16.70% |
| **Preventive medicine (N= 4, 0)** | 0% | 0% |
| **Others (N= 10, 13)** | 20% | 0.00% |

\*Difference between the two responses was assessed by the reviewers as a binary yes/no answer
\*Difference in grading between the two responses was defined as the difference in grading category (1 and 2 vs. 3 and 4)

### Frequently asked questions about HCC

The ChatGPT model was found to provide comprehensive and correct responses to more than 75% of the 73 questions in the categories of “basic knowledge,” “treatment,” “lifestyle,” and “others” (**Table 1**). However, in the “diagnosis” category, 50% of questions were graded as containing a mix of correct and incorrect/outdated information, and 33.3% were graded as incorrect (**Supplemental Table 2**). The model demonstrated a strong ability to provide detailed information on the background knowledge and potential side effects of various treatments for HCC, as well as scientific evidence for lifestyle-related questions. Overall, the two reviewers had 25% of the questions answered differently.

Of note, there were 6.7% of the questions in the “treatment” category in which the ChatGPT model used the TNM stage instead of the BCLC stage to infer survival rates. Additionally, 12.5% of questions in the “lifestyle” category were graded as completely incorrect. For example, the model suggested that diet may potentially reduce the size of HCC, however, there is currently a lack of strong evidence to support this claim. The model also suggested that HCC or its treatment may affect a person’s fertility, while it is actually the presence of cirrhosis that has the largest impact on fertility rather than the treatments used. For the second attempt to generate responses, the ChatGPT only provided responses with a significant difference for one question (**Table 2**).

### Quality measures related to the management of cirrhosis

To examine the knowledge of ChatGPT in cirrhosis care, we formulated the 26 quality measures recommended by the practice metrics committee of the AASLD into questions (**Supplemental Table 3**). The model was able to correctly answer 20 of these measures, resulting in an overall accuracy of 76.9%. One of the areas in which ChatGPT demonstrated proficiency was in accurately describing the algorithm of the initial workup for patients with liver disease, including diagnostic paracentesis, the administration of albumin in patients with a minimum of five liters of ascites removal, and the management of conditions such as spontaneous bacterial peritonitis, ascites, hepatic hydrothorax, esophageal variceal hemorrhage, and hepatic encephalopathy. The model was able to provide appropriate explanations for these topics of questions.

However, there were areas in which the model did not respond correctly or provided outdated information. For example, ChatGPT was not able to indicate the correct cutoff for certain situations, such as liver stiffness measurements indicating the need for upper endoscopy for variceal screening and MELD-Na score for liver transplant evaluation. Additionally, for patients who presented with upper gastrointestinal bleeding, the model failed to correctly indicate the maximum time window recommended for performing an upper endoscopy (12 hours) and the minimum antibiotic course duration for empiric gram negative coverage (5 days).

### Published HCC surveillance knowledge questionnaires

ChatGPT was tested using two published questionnaires that evaluated the knowledge of respondents in HCC and cirrhosis care (**Supplemental Table 4A-B**). In the first study, ChatGPT accurately responded to four of the eight questions, with the proportions of physicians providing correct answers for these questions being 42.9%, 45.4%, 28.8%, and 30.5%, respectively.

ChatGPT was not able to specify the age cut-off for HCC screening in patients with chronic hepatitis B. Although it pinpointed the use of abdominal ultrasound as the first-line screening tool, it failed to identify the use of MRI and CT scan for HCC surveillance in patients with ascites. The proportions of physicians answering correctly for these questions were 33.5% and 32.6%, respectively. ChatGPT correctly identified cirrhosis as an indication for HCC surveillance. However, it wrongly stated that all HBV/HCV patients required it when 47.8% and 42.4% of physicians had the correct responses. The second study contained questions about HCC screening. We showed that ChatGPT had limitations, with only one out of seven questions answered correctly, stating that HCC screening is recommended for patients with cirrhosis. For the additional questions, the model incorrectly emphasized HCC screening for all chronic hepatitis B patients.

### Emotional support to patients and caregivers

We tested ChatGPT’s response to questions regarding coping with psychological stress after receiving a diagnosis of HCC (**Table 3)**. First, it acknowledged the patient’s likely Bemotional response to their diagnosis. Second, ChatGPT provided clear and actionable starting points for a patient who has just been diagnosed with HCC. It provided motivational responses encouraging proactive steps to manage the diagnosis and treatment strategies. Finally, it emphasized the significance of both physical and mental well-being.

**Table 3.** ChatGPT provides emotional support to patients and caregivers.

|  |  |
| --- | --- |
| My family member was recently diagnosed with hepatocellular carcinoma. What can I do to help? | If your family member has been diagnosed with hepatocellular carcinoma, there are several ways you can support them:<br>- Offer practical support: Help with tasks like grocery shopping, cooking, and transportation to appointments.<br>- Be a listening ear: Encourage your family member to talk about their feelings and offer a supportive and understanding ear.<br>- Help them find information: Look into resources and support groups that may be helpful for your family member.<br>- Take care of yourself: It's important to remember to take care of your own physical and emotional health as well. It can be emotionally and physically draining to support a loved one through a serious health condition, so make sure to take breaks and seek support for yourself as needed.<br>- Encourage them to follow their treatment plan: It's important for your family member to follow their treatment plan as recommended by their medical team. Offer support and encouragement to help them stick to their treatment plan.<br>- It's also a good idea to talk to your family member's medical team about other ways you can support them. They may have specific recommendations or suggestions based on your loved one's individual needs. |
| I just received a diagnosis of hepatocellular carcinoma. I am anxious, sad, and confused! What should I do? | I'm sorry to hear about your diagnosis. It's completely natural to feel anxious, sad, and confused when faced with a serious health condition like hepatocellular carcinoma. The most important thing you can do is to talk to your doctor about your treatment options. They will be able to give you more information about your specific case and help you come up with a plan that is right for you. It may also be helpful to speak with a mental health professional, such as a therapist or counselor, to help you cope with your emotions. It's important to take care of your emotional well-being in addition to your physical health. |
| I was recently diagnosed with alcoholic hepatitis and cirrhosis, I am so stressed out and I don't know cope with all these, what should I do? | It can be overwhelming to receive a diagnosis of alcoholic hepatitis and cirrhosis, and it's important to take care of yourself both physically and emotionally. Here are a few things you can do to cope:<br>- Seek professional help: Talk to your doctor or a therapist about your diagnosis and treatment options. They can also help you manage stress and other emotions related to your diagnosis.<br>- Stop drinking: It is important to stop drinking alcohol in order to prevent further damage to your liver and overall health.<br>- Eat a healthy diet: Eating a balanced diet that is rich in fruits, vegetables, and lean protein can help your liver recover and improve your overall health.<br>- Exercise: Regular physical activity can help reduce stress and improve your overall health.<br>- Get enough rest: Make sure to get enough sleep and rest to help your body heal.<br>- Join support groups: Joining support groups can be very helpful in dealing with the emotional and practical aspects of the disease.<br>- Seek medical attention: If you are experiencing severe symptoms such as abdominal pain, jaundice, vomiting blood or a rapid decline in mental status, please seek medical attention.<br>- Remember that recovery takes time and it's important to be patient with yourself. It is also important to keep in touch with your healthcare provider and follow their advice. |
| My father was officially denied for liver transplantation because of his old age and debilitation. The whole family is really upset. What should we do to help him? | It is important to understand that transplant centers have strict guidelines in place to ensure that transplants are performed in the most effective and ethical manner possible. These guidelines take into account factors such as the patient's age, overall health, and likelihood of survival after the transplant.<br>That being said, there are still options for your father. One option would be to seek a second opinion from another transplant center, as different centers may have different guidelines. Additionally, your father may be a candidate for alternative treatments, such as a less invasive procedure or palliative care to help manage his symptoms.<br>It may also be beneficial for your family to seek support from a social worker or counselor to help cope with the difficult news and to discuss any other options that may be available for your father.<br>It's also important to have a good communication with his doctor, ask the reason why he was denied the transplant, and what other options are available for him. |

For caregivers, ChatGPT presented organized and multifaceted psychological and practical recommendations. It suggested caregivers to encourage patients to follow treatment strategies, offer emotional support, and recommend patients to seek sources such as support groups. Notably, the model’s recommendations also included the physical and emotional health of the caregiver in order to improve their resilience.

## Discussion

In this study, we examined the accuracy and reproducibility of the ChatGPT language model in responding to patient questions regarding cirrhosis and HCC. Reviewed by transplant hepatologists, ChatGPT regurgitated extensive knowledge about the two diseases, especially in basic knowledge, lifestyle, and treatment. The model also provided practical and multifaceted advice to patients and caregivers regarding the next steps and adjusting to a new diagnosis. On further examination, ChatGPT demonstrated a strong knowledge base in these two diseases by performing well on the cirrhosis quality measures recommended by the AASLD and previously published questionnaires administered to physicians and trainees. On the other hand, we also highlighted ChatGPT’s limitations in identifying specific cut-off values in the management of cirrhosis and guideline recommendations for surveillance/screening of HCC. The guidelines vary across global regions, and the model was not able to provide tailored recommendations according to the inquirers’ region. ChatGPT may serve as an adjunct information tool for patients with HCC and/or cirrhosis in order to improve outcomes.

Low health literacy can significantly impact care and outcomes in patients with cirrhosis and HCC. Valery et al. showed that poor health literacy was associated with increased medical expenses, cirrhosis-related admissions, and lower quality of life in patients with cirrhosis.^23^ This is particularly concerning given the already high cost of managing cirrhosis and its complications. A study by Farvardin et al. found that patients who knew cirrhosis was a risk factor for developing HCC had higher reported HCC surveillance rates compared to those unaware (odds ratio [OR], 3.09; 95% confidence interval [CI], 1.25–7.62).^24^ Increased regular surveillance of HCC has been shown to increase survival outcomes in patients.^25^ A prior study has shown an association between disease awareness and compliance with the treatment plan.^26^ Accurate and accessible medical information about HCC is essential in order to provide patients with the information they need to make informed decisions about their care and to improve their understanding of the disease and its management. A randomized controlled trial showed that a multifaceted pharmacist-led medication and disease education program was associated with significantly lower unplanned hospitalizations in patients with decompensated cirrhosis compared to usual care.^27^ Despite the importance and advantages of health education, patients are often not aware of resources to obtain accurate and personalized information about their condition. ChatGPT may provide a novel tool that provides increased access to potentially reliable and accurate health information for patients with cirrhosis and HCC. Future studies are needed to better examine the utility of ChatGPT in patient education as well as monitor improvements in accuracy and reproducibility of its responses to patient questions.

One of the strengths of ChatGPT is its ability to sift through massive amounts of information and produce responses in a manner that is conversational and easy to understand. Search results from current search engines can be overwhelming for patients, frequently filled with irrelevant or misleading information. ChatGPT may ameliorate this by providing an easy-to-understand and potentially reliable source of information. As ChatGPT is designed to provide conversational dialogue, the responses can be more comprehensible than professional guidelines or primary literature.

As of writing this article, ChatGPT is available free of charge to the public. This allows patients with financial restraints to access personalized and useful medical information about their condition. This is especially important given that financial difficulty has been associated with poor health outcomes.^28^ Enhancement of availability and ease of access to this resource could reduce health disparities, diminish disease burden, and improve the overall well-being of the population.^29^ Furthermore, this increased accessibility can also reduce anxiety among patients and caregivers by providing access to real-time information. Patients will be better informed about their condition, which could reduce unnecessary anxiety between doctor appointments.

ChatGPT can also enhance the workflow of healthcare providers in two important ways. First, it helps draft a framework for each tailored question asked by patients and caregivers, thereby increasing efficiency for providers. Given the high proportion of either comprehensive or correct but inadequate responses and expected continued improvement over time, we foresee that physicians would only need to revise ChatGPT’s responses to best answer patient queries. This may not only improve the efficiency of physicians but also decrease the overall cost and burden on the healthcare system. Second, ChatGPT empowers patients to be better informed about their care. This allows for patient-led care and facilitates efficient shared decision-making by providing patients with an additional source of information.

The major strengths of this study consist of the inclusive set of inquiries collected from both authorities (professional societies and well-regarded institutions) as well as patients and caregivers (patient support groups). To ensure a holistic grading process of ChatGPT’s responses, two independent hepatologists reviewed the responses, and a third blinded senior hepatologist resolved discrepancies. To our knowledge, this is the first article to examine the accuracy and reproducibility of ChatGPT’s medical knowledge base, along with its ability to answer patient questions.

ChatGPT has garnered interest in transforming the emotional support for patients with mental health disorders.^30^ While it is reasonable to argue that the algorithms used to design the ChatGPT language model may not be robust to completely comprehend and handle the complexities of human emotion, it may still be helpful to individuals who experience specific emotional distress such as psychological support for patients with a cancer diagnosis or their caregivers. In the current study, ChatGPT emulated empathetic responses to the patients and their caregivers and offered actionable recommendations. Studies are needed to assess this capacity from various perspectives, including a formal appraisal of the responses, an understanding of more complex questions, and the adaptation of the model to various cultural backgrounds.

Despite its impressive performance in answering patient questions, ChatGPT has several limitations. First, a small proportion of questions were graded as comprehensive, meaning ChatGPT may serve as an adjunct tool for patients rather than a replacement of care from a licensed healthcare provider. Given that ChatGPT is expected to be continuously improved with time, response quality and reproducibility are likely to improve. Notably, there were 20%-25% differences between the two reviewers. This implies that even specialists may have different thoughts in answering these questions. Second, the database that ChatGPT was trained on only extends to information up until 2021. Information may be outdated for certain subjects, thereby leading to incorrect responses. Third, the quality and accuracy of the dataset utilized to train ChatGPT is unknown, which may affect the reliability of ChatGPT’s responses. In this study, although ChatGPT provided correct or comprehensive answers to the majority of the questions, there was a significant limitation of the model in providing accurate cut-offs for laboratory tests or durations of treatment recommended by professional guidelines. Fourth, guidelines published by different professional societies may vary in recommendations globally. Unless specifically stated in the prompt, ChatGPT is unable to distinguish which guidelines to follow and may not generate the response according to the guidelines of the local area or country. This could generate potential confusion and danger should a patient follow recommendations not approved by local regulatory bodies. Further optimization is warranted to ensure the source of data and the specificity of the response. Perhaps, ChatGPT can be programmed to prompt clarifications from users to fine-tune the questions and provide responses with better accuracy in the future.

## Conclusion

Our study is the first to showcase the accuracy and reproducibility of ChatGPT’s responses to frequently asked questions regarding the management and care of patients with cirrhosis and HCC. ChatGPT often regurgitates correct and reproducible responses to commonly asked patient questions, although the majority of the correct responses were labeled as correct but inadequate. Recommendations that vary across regions, such as the indications and interval of HCC surveillance, should not be followed for the current version of ChatGPT. With the anticipation of the ongoing improvement of the model’s accuracy, ChatGPT may serve as a useful adjunct tool for patients in addition to the standard of care. We highly encourage future studies to examine the utility of ChatGPT in improving patient care and outcomes.

## Data Availability

All data produced in the present work are contained in the manuscript or supplemental material

