## Supplemental Table 1 for "Assessing the performance of ChatGPT in answering questions regarding cirrhosis and hepatocellular carcinoma"

Table of Content

### Supplemental Table 1. Grading of the responses from the ChatGPT Language Model for cirrhosis-related questions

| Questions | Responses 1 | Responses 2 | Reviewer 1 grades 1. Comprehensive  2. Correct but inadequate  3. Mixed with correct and incorrect/outdated data 4. Completely incorrect | Reviewer 2 grades 1. Comprehensive  2. Correct but inadequate  3. Mixed with correct and incorrect/outdated data 4. Completely incorrect | Reviewer 3 grades for the responses with discrepancies |
| --- | --- | --- | --- | --- | --- |
| **Basic Knowledge** | | | | | |
| 1. What is cirrhosis? | Cirrhosis is a type of chronic liver disease that occurs when scar tissue replaces healthy liver tissue. This scar tissue is the result of long-term damage to the liver, which can be caused by a variety of factors, including chronic alcohol abuse, viral hepatitis, and nonalcoholic fatty liver disease.  The liver is an essential organ that performs many important functions, including filtering toxins from the blood, producing bile to aid in digestion, and storing vitamins and minerals. When the liver is damaged, it becomes scarred and is unable to function properly. This can lead to a variety of complications, such as liver failure, liver cancer, and portal hypertension.  Symptoms of cirrhosis may include fatigue, weakness, weight loss, abdominal pain and swelling, and jaundice (a yellowing of the skin and whites of the eyes). Cirrhosis is a serious condition that can be life-threatening if left untreated. Treatment for cirrhosis may include lifestyle changes, medications, and surgery. | Similar | 2 | 2 | N/A |
| 2. What is decompensated cirrhosis? | Decompensated cirrhosis is a stage of liver disease in which the liver is unable to adequately perform its functions due to scarring (cirrhosis) and damage. When the liver is decompensated, it is unable to adequately filter toxins from the blood, produce proteins, and regulate the body's metabolism and blood clotting. This can lead to a range of complications, such as liver failure, bleeding, and infections.  Symptoms of decompensated cirrhosis may include fatigue, loss of appetite, weight loss, abdominal swelling, spider-like blood vessels on the skin, jaundice (yellowing of the skin and whites of the eyes), confusion or changes in mental status, and fluid accumulation in the abdomen (ascites).  Treatment for decompensated cirrhosis may include medications to control symptoms, lifestyle changes to reduce the strain on the liver, and, in some cases, liver transplantation. It is important for people with decompensated cirrhosis to receive timely and appropriate treatment to prevent further damage to the liver and to prevent complications. | Similar | 1 | 2 | 1 |
| 3. What are the symptoms of cirrhosis? | Cirrhosis is a progressive disease that can cause a variety of symptoms, including:  Fatigue: People with cirrhosis often feel tired and weak.  Loss of appetite: Cirrhosis can cause a loss of appetite, which can lead to weight loss.  Nausea and vomiting: These symptoms may be caused by the accumulation of toxins in the body due to liver damage.  Abdominal pain: Cirrhosis can cause abdominal pain due to the build-up of fluid in the abdomen (ascites).  Itching: Cirrhosis can cause itching due to the build-up of bile in the skin.  Yellowing of the skin and whites of the eyes (jaundice): This is caused by the accumulation of bilirubin in the body due to liver damage.  Swelling in the legs: This is caused by the build-up of fluid in the legs (edema) due to liver damage.  Confusion and difficulty thinking: Cirrhosis can cause confusion and difficulty thinking due to the accumulation of toxins in the brain.  Spider-like blood vessels on the skin: These are small, dilated blood vessels that may appear on the skin due to increased pressure in the veins.  Bruising and bleeding easily: Cirrhosis can cause bruising and bleeding easily due to a decrease in the production of clotting factors by the damaged liver.  It's important to note that some people with cirrhosis may not have any symptoms in the early stages of the disease. It's also possible for symptoms to come and go, or to vary in severity over time. If you have any concerns about your health, it's important to speak with a healthcare provider. | Similar | 1 | 2 | 1 |
| 4. How common is cirrhosis? | Cirrhosis is a serious and potentially life-threatening condition that occurs when scar tissue replaces healthy liver tissue, disrupting the normal structure and function of the liver. It can be caused by a variety of factors, including chronic alcohol abuse, viral hepatitis, and nonalcoholic fatty liver disease.  The prevalence of cirrhosis varies depending on the population being studied and the specific cause of the condition. According to the World Health Organization (WHO), cirrhosis is the 12th leading cause of death globally and is responsible for approximately 1.3 million deaths each year. In the United States, the prevalence of cirrhosis is estimated to be between 0.5% and 1% of the population, with the highest rates seen in middle-aged men. Nonalcoholic fatty liver disease is the most common cause of cirrhosis in the US, followed by hepatitis C and alcohol abuse. | Similar | 1 | 1 | N/A |
| 5. Who is more likely to get cirrhosis? | Cirrhosis is a serious liver condition that occurs when healthy liver tissue is replaced with scar tissue. This can lead to liver failure and other serious health problems. There are a number of factors that can increase a person's risk of developing cirrhosis, including:  Alcohol abuse: Excessive alcohol consumption is a major cause of cirrhosis.  Hepatitis: Chronic infection with the hepatitis B or C virus can cause cirrhosis.  Nonalcoholic fatty liver disease (NAFLD): This condition, which is caused by a buildup of excess fat in the liver, can lead to cirrhosis in some people.  Genetic factors: Some people may be more prone to developing cirrhosis due to genetic factors.  Other medical conditions: Certain medical conditions, such as cystic fibrosis and Wilson's disease, can increase the risk of cirrhosis.  It is important to note that anyone can develop cirrhosis, but some people may be more at risk due to the factors listed above. It is important to reduce your risk of cirrhosis by avoiding excessive alcohol consumption, getting vaccinated against hepatitis B, and taking steps to prevent or manage medical conditions that increase your risk. | Similar | 1 | 2 | 1 |
| 6. What vaccines are needed in people with cirrhosis? | Cirrhosis is a serious condition that is caused by long-term damage to the liver. People with cirrhosis are at increased risk of serious complications from certain infections, so it is important for them to be up to date on their vaccinations.  The specific vaccines that are recommended for people with cirrhosis will depend on several factors, including their age, overall health, and their risk of exposure to certain infections. However, some vaccines that are often recommended for people with cirrhosis include:  Influenza (flu) vaccine: People with cirrhosis are at increased risk of serious complications from the flu, so it is important for them to get vaccinated each year.  Pneumococcal vaccine: This vaccine helps to protect against pneumonia and other infections caused by the pneumococcus bacteria. It is usually recommended for people with cirrhosis, especially those who are over the age of 65 or have certain other risk factors.  Hepatitis A and B vaccines: These vaccines help to protect against hepatitis A and hepatitis B, which are viral infections that can cause serious liver damage. People with cirrhosis are at increased risk of complications from these infections, so it is important for them to be vaccinated.  Tetanus and diphtheria (Td) vaccine: This vaccine helps to protect against tetanus and diphtheria, which are serious bacterial infections. It is usually recommended for all adults, including those with cirrhosis.  Zoster (shingles) vaccine: This vaccine helps to protect against shingles, which is a painful skin rash caused by the herpes zoster virus. It is usually recommended for adults over the age of 50, including those with cirrhosis.  It is important to talk to your healthcare provider about which vaccines are recommended for you, as your specific vaccination needs may vary depending on your individual circumstances. | Similar | 3 | 2 | 2 |
| 7. Why are vaccinations important for cirrhosis? | Vaccinations are important for people with cirrhosis because they can help prevent infections that can be more serious or even life-threatening in people with weakened immune systems. Cirrhosis is a condition in which the liver becomes scarred and is unable to function properly. It can be caused by a number of things, including chronic alcohol abuse, viral hepatitis, and nonalcoholic fatty liver disease.  People with cirrhosis often have a weakened immune system, which means they are more susceptible to infections. Vaccinations can help protect against some of the most common and serious infections that can occur in people with cirrhosis, such as pneumonia, influenza, and hepatitis A and B.  It's important to talk to your healthcare provider about which vaccinations are recommended for you if you have cirrhosis. They will be able to provide you with information about the benefits and potential risks of each vaccination and help you make an informed decision about which ones to get. | Similar | 1 | 2 | 1 |
| 8. How much alcohol does it take before causing liver damage? | The amount of alcohol that can cause liver damage varies from person to person and depends on several factors, including:  The amount of alcohol consumed: Heavy drinking, defined as more than 3 drinks per day for women and more than 4 drinks per day for men, can lead to liver damage over time.  Body weight: Larger people generally have more body mass to dilute the alcohol, so they can tolerate more alcohol before it starts to cause damage.  Age: Older people may be more sensitive to the effects of alcohol and may be at increased risk of liver damage.  Gender: Women may be more susceptible to alcohol-related liver damage than men, even if they drink the same amount of alcohol.  Family history: People with a family history of alcohol abuse or liver disease may be more prone to liver damage from alcohol.  It's important to note that any amount of alcohol can be harmful to the liver in the long term, and it's best to drink in moderation or not at all. The Centers for Disease Control and Prevention (CDC) recommends that women limit their alcohol intake to one drink per day, and men to two drinks per day. One drink is defined as 12 ounces of beer, 5 ounces of wine, or 1.5 ounces of 80-proof distilled spirits. | It is difficult to determine exactly how much alcohol it takes to cause liver damage because it can vary depending on a number of factors, including the person's age, sex, weight, and overall health. In general, however, heavy alcohol consumption over a long period of time can lead to liver damage.  The Centers for Disease Control and Prevention (CDC) defines heavy alcohol use as more than 8 drinks per week for women and more than 15 drinks per week for men. This level of alcohol consumption may increase the risk of liver damage, as well as other serious health problems such as high blood pressure, pancreatitis, and certain types of cancer.  It is important to note that even moderate alcohol consumption can be harmful to the liver and other organs if it is not consumed responsibly. If you are concerned about your alcohol consumption and its potential impact on your health, it is a good idea to speak with a healthcare provider. | 3,1 | 3,2 | 3,1 |
| 9. What is Hepatitis? | Hepatitis is a general term that refers to an inflammation of the liver. The liver is a vital organ that plays a key role in many of the body's functions, including processing nutrients, producing bile, and removing toxins from the body. Hepatitis can be caused by a variety of factors, including viral infections, alcohol abuse, and certain medications.  There are several types of hepatitis, including hepatitis A, B, C, D, and E. Each type is caused by a different virus and can have different symptoms, modes of transmission, and treatment options.  Hepatitis A is caused by the hepatitis A virus (HAV) and is typically transmitted through contaminated food or water. It can also be transmitted through close personal contact with an infected person. Hepatitis A can be prevented through vaccination.  Hepatitis B is caused by the hepatitis B virus (HBV) and is transmitted through contact with blood or other bodily fluids from an infected person. This can occur through sexual contact, sharing needles or other injection drug use equipment, or exposure to contaminated blood or blood products. Hepatitis B can be prevented through vaccination.  Hepatitis C is caused by the hepatitis C virus (HCV) and is transmitted through contact with blood from an infected person. This can occur through sharing needles or other injection drug use equipment, or exposure to contaminated blood or blood products. Hepatitis C can be treated with antiviral medications.  Hepatitis D is caused by the hepatitis D virus (HDV) and can only occur in people who are already infected with hepatitis B. It is transmitted through contact with blood or other bodily fluids from an infected person.  Hepatitis E is caused by the hepatitis E virus (HEV) and is transmitted through contaminated food or water. It is more common in developing countries and is typically self-limiting, meaning it goes away on its own without treatment.  Symptoms of hepatitis can include fever, fatigue, loss of appetite, nausea, vomiting, abdominal pain, dark urine, clay-colored bowel movements, joint pain, and jaundice (yellowing of the skin and whites of the eyes). In some cases, hepatitis can lead to more serious complications, such as liver failure or liver cancer. If you think you may have hepatitis, it is important to see a healthcare provider for proper diagnosis and treatment. | Similar | 2 | 1 | 2 |
| 10. How to slow or reverse cirrhosis? | Cirrhosis is a serious and irreversible condition that occurs when scar tissue replaces healthy liver tissue. It is often the result of long-term damage to the liver from conditions such as alcohol abuse, hepatitis, or nonalcoholic fatty liver disease.  There is no cure for cirrhosis, and the damage it causes to the liver is permanent. However, it is possible to slow the progression of the disease and prevent further damage to the liver by making lifestyle changes and receiving appropriate medical treatment. Here are some steps that may help:  Stop drinking alcohol: If you have cirrhosis due to alcohol abuse, it is important to stop drinking alcohol completely. This will help to prevent further damage to your liver.  Eat a healthy diet: A healthy diet that is low in salt and fat can help to reduce the strain on your liver and improve your overall health.  Exercise regularly: Regular exercise can improve your overall health and help to reduce the risk of complications from cirrhosis.  Get vaccinated: Vaccines can protect you from certain types of hepatitis, which is a common cause of cirrhosis.  Take medications as prescribed: If you have been prescribed medications to treat your cirrhosis or any underlying condition, be sure to take them as directed by your healthcare provider.  Avoid exposure to toxins: Certain substances, such as pesticides and certain types of medications, can be harmful to the liver. Avoid exposure to these substances as much as possible.  Monitor your health: Regular check-ups with your healthcare provider can help to identify any complications or new problems that may be developing.  It is important to note that these steps may help to slow the progression of cirrhosis, but they cannot reverse the damage that has already been done to the liver. If you have cirrhosis, it is important to work closely with your healthcare provider to manage your condition and prevent complications. | Similar | 2 | 2 | N/A |
| 11. What are the complications of cirrhosis? | Cirrhosis is a serious and potentially life-threatening condition that occurs when the liver becomes scarred and damaged. It can lead to a number of complications, including:  Ascites: This is the accumulation of fluid in the abdomen, which can cause bloating and discomfort.  Hepatic encephalopathy: This is a condition in which toxins build up in the blood and affect brain function, leading to symptoms such as confusion, drowsiness, and tremors.  Variceal bleeding: Cirrhosis can cause the blood vessels in the esophagus and stomach to become enlarged and prone to bleeding.  Hepatorenal syndrome: This is a type of kidney failure that can occur in people with cirrhosis.  Infections: People with cirrhosis are more susceptible to infections, such as pneumonia and spontaneous bacterial peritonitis (an infection of the abdomen).  Hepatocellular carcinoma: This is a type of liver cancer that can occur in people with cirrhosis.  Liver failure: Cirrhosis can eventually lead to liver failure, which can be life-threatening.  It's important to manage cirrhosis through a combination of lifestyle changes, medication, and possibly a liver transplant to prevent these complications. | Similar | 1 | 2 | 2 |
| 12. Is cirrhosis a hereditary disease? | Cirrhosis is a condition in which the liver becomes scarred and damaged, leading to impaired function. It can be caused by a variety of factors, including viral hepatitis, alcohol abuse, and nonalcoholic fatty liver disease. While some forms of cirrhosis have a genetic component and may be more common in certain families, in most cases it is not a hereditary disease.  In some cases, certain genetic conditions can increase the risk of developing cirrhosis. For example, individuals with certain genetic mutations may be at an increased risk of developing nonalcoholic fatty liver disease, which can lead to cirrhosis. However, these genetic conditions are relatively rare and do not account for the majority of cases of cirrhosis.  It's important to note that while cirrhosis itself may not be hereditary, certain lifestyle factors that can contribute to its development, such as alcohol consumption and diet, can be influenced by family habits and may be passed down from one generation to the next. Therefore, it is important for individuals to be aware of their family history and to take steps to reduce their risk of developing cirrhosis or other liver diseases. This may include adopting a healthy lifestyle, including a balanced diet and avoiding or limiting alcohol consumption. | Similar | 2 | 1 | 2 |
| 13. Can cirrhosis kill? | Cirrhosis is a serious and potentially life-threatening condition that occurs when scar tissue replaces healthy liver tissue. The scarring that occurs in cirrhosis can interfere with the liver's ability to function properly, leading to a range of complications such as bleeding, infections, and liver failure.  In some cases, cirrhosis can be reversible if it is caught early and the underlying cause is treated. However, if the damage to the liver is severe and the person does not receive treatment, cirrhosis can be fatal. The prognosis for someone with cirrhosis will depend on the severity of the condition, the underlying cause, and whether the person receives treatment.  It is important for people with cirrhosis to receive medical care and follow their treatment plan to improve their chances of a good outcome. This may include medications, lifestyle changes, and in some cases, a liver transplant. | Similar | 1 | 2 | 1 |
| 14. Is cirrhosis painful? | Cirrhosis is a serious and often life-threatening condition that occurs when scar tissue replaces healthy liver tissue, leading to liver dysfunction and failure. While cirrhosis itself may not cause pain, many people with cirrhosis experience discomfort or pain due to the complications associated with the condition. For example, some people with cirrhosis may experience abdominal pain due to the build-up of fluid in the abdomen (known as ascites), or they may experience pain in the upper right part of the abdomen due to an enlarged liver. In addition, people with cirrhosis may develop liver cancer, which can cause pain. It is important for people with cirrhosis to seek medical attention if they experience any pain, as it could be a sign of a serious complication. | Similar | 1 | 1 | N/A |
| 15. Can cirrhosis cause cancer? | Cirrhosis, a condition in which scar tissue replaces healthy liver tissue, can increase the risk of liver cancer. This is because cirrhosis can cause changes in the liver that can lead to the development of cancer cells.  The exact cause of liver cancer is not fully understood, but several factors can increase the risk, including chronic viral hepatitis (such as hepatitis B or C), heavy alcohol consumption, and cirrhosis. People with cirrhosis have an increased risk of developing liver cancer because the scar tissue that forms in the liver as a result of cirrhosis can cause changes in the liver cells that can lead to the development of cancer.  It's important to note that not everyone with cirrhosis will develop liver cancer. However, it's important for people with cirrhosis to be monitored regularly for any signs or symptoms of liver cancer, such as abdominal pain, a lump or mass in the abdomen, unexplained weight loss, or yellowing of the skin and whites of the eyes (jaundice). Early detection and treatment of liver cancer can improve the chances of a successful outcome. | Similar | 1 | 1 | N/A |
| 16. How to prevent cirrhosis? | Cirrhosis is a serious and potentially life-threatening condition that occurs when scar tissue replaces healthy liver tissue, leading to liver dysfunction. It is often the result of chronic damage to the liver, such as from alcohol abuse, hepatitis, or other chronic liver diseases.  To prevent cirrhosis, it is important to take steps to protect your liver and avoid activities that can damage it. Some ways to do this include:  Avoiding excessive alcohol consumption: Excessive alcohol consumption is a leading cause of cirrhosis. It is important to limit your alcohol intake or abstain from alcohol completely to reduce your risk of developing cirrhosis.  Getting vaccinated against hepatitis: Hepatitis B and C are viral infections that can cause chronic liver disease and cirrhosis. Getting vaccinated against hepatitis B and getting tested for hepatitis C can help protect against these infections and reduce your risk of cirrhosis.  Managing other health conditions: Some health conditions, such as obesity, type 2 diabetes, and nonalcoholic fatty liver disease, can increase your risk of cirrhosis. It is important to manage these conditions and maintain a healthy lifestyle to reduce your risk.  Avoiding risky behaviors: Engaging in risky behaviors, such as sharing needles or engaging in unprotected sex, can increase your risk of hepatitis and other infections that can lead to cirrhosis. It is important to practice safe behaviors to reduce your risk.  Getting regular check-ups: Regular check-ups with your healthcare provider can help identify any potential liver issues early on, allowing for timely treatment and management to reduce the risk of cirrhosis. | Similar | 1 | 1 | N/A |
| 17. Is cirrhosis reversible? | Cirrhosis is a type of liver disease that is characterized by scarring of the liver tissue. In advanced stages, cirrhosis can lead to serious complications such as liver failure, portal hypertension, and liver cancer.  In some cases, cirrhosis may be reversible if it is caught early and the underlying cause is treated effectively. For example, if the cause of cirrhosis is alcohol abuse, abstaining from alcohol and getting treatment for alcohol dependence may halt the progression of the disease and potentially reverse it. However, if the disease has progressed to advanced stages, it may not be reversible.  It is important to seek medical attention if you are experiencing symptoms of cirrhosis or if you are at risk for developing the disease due to certain lifestyle factors or underlying medical conditions. Early diagnosis and treatment can improve your chances of a successful outcome. | Cirrhosis is a condition in which the liver becomes scarred and damaged, leading to a loss of its normal structure and function. It is generally considered to be a progressive and irreversible condition, meaning that it typically gets worse over time and cannot be reversed. However, it is possible to slow the progression of cirrhosis and improve symptoms through early diagnosis and treatment of the underlying cause, such as by treating hepatitis or alcohol abuse.  In some cases, cirrhosis may be reversible if it is caused by a condition that can be treated or resolved. For example, if cirrhosis is caused by excessive alcohol consumption, it may be reversible if the person stops drinking alcohol and receives proper medical treatment. Similarly, if cirrhosis is caused by a condition such as nonalcoholic fatty liver disease, it may be reversible if the person makes lifestyle changes such as losing weight, exercising, and eating a healthy diet.  It is important to note that cirrhosis is a serious condition that can have serious complications, such as liver failure, and it can be life-threatening if not treated properly. If you have been diagnosed with cirrhosis or are at risk of developing the condition, it is important to follow your doctor's recommendations for treatment and to make any necessary lifestyle changes to help manage the condition. | 1,2 | 2,1 | 1,2 |
| 18. What is hepatic encephalopathy? | Hepatic encephalopathy is a condition that occurs when the liver is not able to function properly and remove toxic substances from the blood. These toxic substances can build up in the brain and cause a range of symptoms, including changes in mental function, behavior, and consciousness.  The symptoms of hepatic encephalopathy can range from mild to severe, and may include changes in mood, memory, and concentration; tremors or tremors; disorientation; confusion; slurred speech; changes in sleep patterns; and changes in motor function. In severe cases, hepatic encephalopathy can lead to coma or death.  There are several potential causes of hepatic encephalopathy, including liver failure, cirrhosis (chronic scarring of the liver), hepatitis (inflammation of the liver), and toxic substances that damage the liver. The condition can also be triggered by certain medications, infections, and bleeding in the digestive tract.  Treatment for hepatic encephalopathy typically involves addressing the underlying cause of the condition and controlling the symptoms. This may involve medications to remove toxic substances from the blood, changes to diet and nutrition, and supportive care such as artificial ventilation. In some cases, a liver transplant may be necessary. | Similar | 1 | 2 | 2 |
| 19. What are the symptoms of hepatic encephalopathy in patients with cirrhosis? | Hepatic encephalopathy is a condition that occurs when there is damage to the liver, resulting in the accumulation of toxins in the body that can affect brain function. In patients with cirrhosis, the most common symptoms of hepatic encephalopathy include:  Confusion and disorientation: This can range from mild forgetfulness to severe confusion and loss of awareness of surroundings.  Changes in behavior and personality: Patients may become more irritable, aggressive, or withdrawn, and may exhibit unusual or bizarre behavior.  Changes in mental function: Patients may have difficulty thinking, concentrating, or making decisions. They may also experience difficulty speaking or writing.  Changes in physical functioning: Patients may have difficulty walking or moving, and may exhibit tremors or twitching.  Changes in sleep patterns: Patients may have difficulty falling asleep or may sleep more than usual.  Changes in appetite: Patients may lose their appetite or may have a sudden increase in appetite.  It's important to note that these symptoms may vary in severity and may come and go. If you are experiencing any of these symptoms, it's important to speak with a healthcare professional as soon as possible. | Similar | 1 | 2 | 1 |
| 20. How to prevent hepatic encephalopathy? | Hepatic encephalopathy is a condition that can occur as a result of liver disease or damage. It is characterized by changes in brain function, including confusion, personality changes, and difficulty with muscle control and coordination. There are several steps that can be taken to help prevent hepatic encephalopathy or reduce the risk of developing it:  Control and treat the underlying cause of liver disease: This may include taking medications as prescribed, following a healthy diet, avoiding alcohol, and managing other health conditions such as viral hepatitis or cirrhosis.  Monitor and manage ammonia levels: Ammonia is a substance that is produced by the breakdown of protein in the body. High levels of ammonia can lead to hepatic encephalopathy. Your healthcare provider may recommend medications or other treatments to help control ammonia levels.  Manage constipation: Constipation can lead to an accumulation of toxins in the body, which can worsen hepatic encephalopathy. Staying hydrated and eating a high-fiber diet can help prevent constipation.  Avoid certain medications: Some medications can worsen hepatic encephalopathy, so it is important to discuss all medications with your healthcare provider.  Follow a low-protein diet: A low-protein diet may help reduce the production of ammonia and reduce the risk of hepatic encephalopathy. Your healthcare provider or a dietitian can help you develop a low-protein diet plan that meets your nutritional needs.  Manage fluid and electrolyte balance: It is important to maintain a healthy balance of fluids and electrolytes in the body to help prevent hepatic encephalopathy. Your healthcare provider may recommend medications or other treatments to help manage fluid and electrolyte balance.  It is important to work closely with your healthcare provider to develop a treatment plan that is appropriate for your specific needs. | Similar | 3 | 3 | N/A |
| 21. What is the criteria for liver transplant? | The criteria for liver transplantation vary depending on the specific situation and the transplant center, but there are some general factors that are typically considered when determining whether a person is a candidate for liver transplantation.  Some of the factors that may be considered in the evaluation for liver transplantation include:  Severity of liver disease: People with severe liver disease that is causing symptoms or complications, or that is likely to progress to end-stage liver disease, may be candidates for liver transplantation.  Other medical conditions: In order to be a candidate for liver transplantation, a person must be generally healthy enough to undergo the procedure and to recover from it. This means that other medical conditions, such as heart disease or uncontrolled infections, may need to be treated or controlled before transplantation can be considered.  Ability to comply with post-transplant care: A person must be able to follow the prescribed treatment and care regimen after transplantation in order to have the best chance of a successful outcome. This includes taking medications as prescribed and attending follow-up appointments.  Age: While there is no strict age limit for liver transplantation, older people may be less likely to be considered candidates due to the increased risks and challenges of surgery and recovery at an advanced age.  Availability of a suitable organ: The availability of a suitable donor liver is also a factor in the decision to undergo transplantation. In some cases, people may be placed on a waiting list for a liver transplant, and the transplant may be performed when a suitable organ becomes available.  It is important to note that the specific criteria for liver transplantation can vary from one transplant center to another, and the decision to proceed with transplantation is ultimately made on a case-by-case basis. | Similar | 1 | 2 | 2 |
| 22. What are the symptoms of spontaneous bacterial peritonitis? | Spontaneous bacterial peritonitis (SBP) is a serious infection of the abdomen that can cause symptoms such as:  Abdominal pain: This is often severe and may be felt in the upper right or upper left part of the abdomen.  Nausea and vomiting: These may be present if the abdominal pain is severe.  Fever: A high fever may be present, often above 101°F (38.3°C).  Decreased urine output: The person may urinate less frequently or produce less urine than usual.  Confusion or altered mental status: The person may be confused or disoriented, or may have difficulty thinking clearly.  Rapid breathing: The person may breathe faster than normal.  Rapid heart rate: The person's heart may beat faster than normal.  Swelling of the abdomen: The abdomen may appear swollen or distended.  If you are experiencing any of these symptoms, it is important to seek medical attention as soon as possible. SBP can be a life-threatening condition if not treated promptly. | Similar | 1 | 2 | 2 |
| 23. What is spontaneous bacterial peritonitis? | Spontaneous bacterial peritonitis (SBP) is a serious and potentially life-threatening condition that occurs when bacteria enter and infect the peritoneum, the thin layer of tissue that lines the inner wall of the abdomen and covers the abdominal organs. SBP typically develops in people who have cirrhosis, a chronic liver disease that leads to scarring and damage of the liver. However, it can also occur in people with other conditions that cause fluid to accumulate in the abdomen, such as kidney failure, pancreatitis, and peritoneal dialysis.  The symptoms of SBP can be severe and may include abdominal pain, fever, nausea, vomiting, and rapid breathing. Left untreated, SBP can lead to sepsis, a serious and potentially life-threatening condition that occurs when the body's immune system overreacts to an infection, leading to inflammation and damage to the body's own tissues and organs.  Treatment of SBP usually involves antibiotics and fluid replacement, and may also include surgery to remove any infected or necrotic (dead) tissue. In severe cases, hospitalization may be necessary. It is important to seek medical attention as soon as possible if you suspect that you or someone you know may have SBP. Early diagnosis and treatment can improve the chances of a full recovery. | Similar | 1 | 2 | 3 |
| 24. How to prevent and manage spontaneous bacterial peritonitis in patients with cirrhosis? | Spontaneous bacterial peritonitis (SBP) is a serious complication that can occur in patients with cirrhosis, a condition in which the liver is damaged and scarred. SBP is caused by bacteria entering the abdomen and causing inflammation of the lining of the abdomen (peritoneum). It can lead to serious complications and can be life-threatening if not treated promptly.  To prevent SBP, it is important for patients with cirrhosis to receive proper medical care and follow their treatment plan. This may include taking medications to reduce the risk of infection, such as antibiotics or antiviral drugs. It is also important for patients to maintain good hygiene, including washing their hands regularly and avoiding contact with people who are sick.  If SBP is suspected, it is important to seek medical attention immediately. Treatment may include intravenous antibiotics, fluid replacement, and other supportive care. In severe cases, surgery may be necessary to remove infected tissue or to repair any damage to the abdomen.  It is also important for patients with cirrhosis to receive regular check-ups with their healthcare provider to monitor their condition and to identify any potential complications early on. This can help prevent serious complications and improve the chances of a successful outcome. | Similar | 1 | 2 | 3 |
| 25. What are varices? | Varices are dilated or enlarged veins that are usually found in the esophagus or the lower part of the stomach. They are often seen in people with liver cirrhosis, a condition in which the liver is damaged and scarred and is unable to function properly. Varices can also be found in the legs, but they are more commonly found in the upper part of the body.  Varices can cause a number of problems, including bleeding, which can be life-threatening. When varices bleed, they can cause a person to vomit blood or have black, tarry stools. If you have varices and experience any of these symptoms, it is important to seek medical attention immediately.  Treatment for varices often involves medications to reduce the risk of bleeding, as well as procedures to close or remove the dilated veins. In some cases, a person with varices may need a liver transplant to repair the damaged liver. | Similar | 3 | 3 | N/A |
| 26. What are the symptoms and complications of varices in cirrhosis? | Varices are dilated, swollen veins that often occur in the esophagus or stomach of people with cirrhosis, a type of liver disease. The main symptoms of varices in cirrhosis are:  Hematemesis (vomiting of blood): This is the most common symptom of varices and is usually the result of a variceal bleed, which occurs when a varix ruptures and bleeds. Hematemesis can also be accompanied by hematochezia (passage of blood through the rectum).  Dysphagia (difficulty swallowing): Varices can cause dysphagia by narrowing the esophagus and making it difficult for food and liquids to pass through.  Abdominal pain: Varices can cause abdominal pain due to the presence of blood in the abdomen, which can lead to irritation and inflammation.  Weakness: Varices can cause weakness due to blood loss, which can lead to anemia.  Complications of varices in cirrhosis include:  Variceal bleeding: This is the most serious complication of varices and can be life-threatening if not treated promptly.  Esophageal stricture: Varices can cause scarring of the esophagus, leading to a narrowing of the esophagus (esophageal stricture). This can cause difficulty swallowing and lead to malnutrition.  Portal hypertension: Varices can cause increased pressure in the portal vein, which can lead to the development of portal hypertension. Portal hypertension can cause a number of complications, including ascites (build-up of fluid in the abdomen) and hepatic encephalopathy (a condition that affects brain function).  It is important to seek medical attention if you have cirrhosis and experience any of the symptoms or complications associated with varices. Early diagnosis and treatment can help prevent serious complications and improve your chances of recovery. | Similar | 3 | 3 | N/A |
| 27. How to prevent and manage esophageal varices in patients with cirrhosis? | Esophageal varices are enlarged veins in the lining of the esophagus (the tube that carries food from the mouth to the stomach). They can occur in people with cirrhosis, which is a condition in which the liver is damaged and scarred and can't function properly.  There are several steps that can be taken to prevent and manage esophageal varices in people with cirrhosis:  Treat the underlying cause of cirrhosis: The most effective way to prevent esophageal varices is to treat the underlying cause of cirrhosis. This may involve medications to control conditions such as hepatitis or alcohol abuse, or lifestyle changes such as eating a healthy diet and exercising.  Lower the pressure in the portal vein: The portal vein carries blood from the digestive organs to the liver. In people with cirrhosis, the pressure in the portal vein can be high, which can cause esophageal varices. Medications called beta blockers can be used to lower the pressure in the portal vein and reduce the risk of esophageal varices.  Endoscopic band ligation: This is a procedure in which a rubber band is placed around the base of an esophageal varix to cut off its blood supply. This can help to shrink the varix and prevent it from bleeding.  Sclerotherapy: In this procedure, a chemical solution is injected into the varix to shrink it.  TIPS (transjugular intrahepatic portosystemic shunt): This is a procedure in which a small tube is inserted into the liver to create a new pathway for blood to flow around the damaged liver. This can help to lower the pressure in the portal vein and reduce the risk of esophageal varices.  It's important to follow your doctor's recommendations for managing esophageal varices and to attend regular follow-up appointments to monitor for any changes in the varices. | Similar | 1 | 2 | 1 |
| 28. What are the symptoms and complications of gastrointestinal bleeding in cirrhosis? | Gastrointestinal (GI) bleeding is a common complication of cirrhosis, a chronic liver disease in which scar tissue replaces healthy liver tissue and impairs the liver's ability to function properly. GI bleeding can occur anywhere in the digestive tract from the mouth to the rectum, but it is most common in the esophagus and stomach.  Symptoms of GI bleeding in cirrhosis may include:  Vomiting blood or coffee-ground material (a sign of bleeding in the upper digestive tract)  Black, tarry stools (a sign of bleeding in the lower digestive tract)  Weakness and dizziness due to blood loss  Swelling in the abdomen due to fluid accumulation (ascites)  Abdominal pain  Fever  Complications of GI bleeding in cirrhosis may include:  Anemia (low red blood cell count) due to blood loss  Shock, a life-threatening condition that occurs when the body doesn't have enough blood and oxygen  Liver failure, which can occur if the bleeding is severe or if the liver is already severely damaged  Death, which is more likely in people with advanced cirrhosis and those who have experienced multiple episodes of GI bleeding.  If you have cirrhosis and experience any of the above symptoms, it is important to seek medical attention immediately as GI bleeding can be life-threatening. Your doctor will likely recommend tests to determine the location and cause of the bleeding and may recommend treatment options such as blood transfusions, medications to control bleeding, or endoscopic procedures to stop the bleeding. | Similar | 1 | 2 | 2 |
| 29. How to prevent and manage gastrointestinal bleeding in patients with cirrhosis? | Gastrointestinal bleeding is a serious complication that can occur in patients with cirrhosis, a condition in which the liver is scarred and no longer functions properly. The following steps can help prevent and manage gastrointestinal bleeding in patients with cirrhosis:  Control blood pressure: High blood pressure in the veins that supply blood to the liver (portal hypertension) is a common cause of gastrointestinal bleeding in patients with cirrhosis. Therefore, it is important to control blood pressure to reduce the risk of bleeding. This can be done through medications such as beta blockers, which can help lower blood pressure in the veins.  Treat underlying causes of bleeding: If the bleeding is caused by a specific problem, such as a bleeding ulcer or a tumor, it is important to treat the underlying cause. This may involve medications or surgery.  Use proton pump inhibitors: Proton pump inhibitors (PPIs) are medications that can help reduce the risk of bleeding by decreasing the production of acid in the stomach.  Endoscopy: Endoscopy is a procedure in which a flexible tube with a camera on the end is inserted through the mouth to visualize the inside of the digestive tract. This can help identify the cause of bleeding and allow for treatment, such as applying heat or using a laser to stop the bleeding.  Transfusions: If the patient is losing a significant amount of blood, a transfusion may be necessary to replace lost blood and prevent further complications.  Surgery: In some cases, surgery may be necessary to stop the bleeding or repair damage caused by the bleeding.  It is important for patients with cirrhosis to work closely with their healthcare team to identify the cause of gastrointestinal bleeding and implement appropriate treatment measures. | Similar | 1 | 3 | 3 |
| 30. What is ascites in cirrhosis? | Ascites is the accumulation of fluid in the abdomen. It is a common complication of cirrhosis, a progressive liver disease that occurs when scar tissue replaces healthy liver tissue, blocking the flow of blood and hindering the liver's ability to function properly. The scar tissue interferes with the liver's ability to produce proteins, process hormones and drugs, and remove toxins from the blood.  Ascites is caused by an imbalance in the production and removal of fluid in the body. In cirrhosis, this is often due to high levels of pressure in the blood vessels of the liver and reduced function of the liver cells. The pressure in the blood vessels can cause fluid to leak out of the blood vessels and into the abdominal cavity, leading to the accumulation of fluid.  Symptoms of ascites in cirrhosis may include abdominal bloating and swelling, difficulty breathing, and a feeling of fullness or pressure in the abdomen. The condition can be uncomfortable and can cause difficulties with daily activities. Treatment may involve medications to reduce fluid buildup and remove excess fluid through a procedure called paracentesis, as well as lifestyle changes and treatment for the underlying liver disease. | Similar | 1 | 2 | 1 |
| 31. What are the symptoms and complications of ascites in cirrhosis? | Ascites is the accumulation of fluid in the abdomen, and it is a common complication of cirrhosis, a progressive liver disease. Symptoms of ascites may include:  Abdominal swelling: The abdomen may appear swollen or distended due to the accumulation of fluid.  Abdominal discomfort or pain: The increased pressure in the abdomen caused by the fluid accumulation can cause discomfort or pain.  Difficulty breathing: The fluid accumulation can put pressure on the diaphragm, making it difficult to breathe.  Changes in bowel movements: The fluid accumulation can cause constipation or diarrhea.  Weight gain: The fluid accumulation can cause rapid weight gain.  Complications of ascites can include:  Infection: The fluid accumulation can create a breeding ground for bacteria, leading to infections such as spontaneous bacterial peritonitis.  Hepatorenal syndrome: This is a serious complication that occurs when the kidneys stop working properly due to cirrhosis. It can lead to kidney failure.  Hepatopulmonary syndrome: This occurs when there is an abnormal connection between the blood vessels in the liver and the lungs, leading to difficulty breathing.  Gastrointestinal bleeding: The fluid accumulation can put pressure on blood vessels in the gastrointestinal tract, leading to bleeding.  It's important to seek medical attention if you have cirrhosis and experience symptoms of ascites. Treatment may include medications to remove excess fluid, a low-sodium diet, and possibly a procedure to remove the fluid. In some cases, a liver transplant may be necessary. | Ascites is the accumulation of fluid in the abdomen. It is a common complication of cirrhosis, a chronic liver disease. Symptoms of ascites can include:  Abdominal bloating and discomfort  Feeling full after eating only a small amount of food  Difficulty breathing due to pressure on the diaphragm  Swelling in the legs and ankles  Weight gain  Complications of ascites can include:  Spontaneous bacterial peritonitis (SBP): This is an infection of the fluid in the abdomen. Symptoms include fever, abdominal pain, and nausea. SBP can be life-threatening and requires immediate treatment.  Hepatorenal syndrome: This is a type of kidney failure that can occur in people with cirrhosis. Symptoms include fatigue, confusion, and changes in urine output.  Hepatopulmonary syndrome: This is a condition in which the liver is not functioning properly and there is an abnormal expansion of the blood vessels in the lungs. Symptoms include shortness of breath and a bluish tint to the skin and mucous membranes (cyanosis).  Portal hypertension: This is high blood pressure in the vein that carries blood from the intestine to the liver (the portal vein). It can lead to the formation of abnormal blood vessels (varices) in the esophagus or stomach, which can rupture and cause bleeding.  If you are experiencing symptoms of ascites or any other complications of cirrhosis, it is important to seek medical attention as soon as possible. Early treatment can help prevent or delay the progression of the disease and reduce the risk of complications. | 3,3 | 3,3 | N/A |
| 32. How to prevent and manage ascites in patients with cirrhosis? | Ascites is a common complication of cirrhosis, a condition in which scarring of the liver interferes with its ability to function properly. Ascites occurs when fluid builds up in the abdominal cavity, causing discomfort, abdominal distention, and difficulty breathing. It can also lead to other complications such as infection and malnutrition.  To prevent and manage ascites in patients with cirrhosis, it is important to follow a healthy lifestyle and to manage any underlying medical conditions that may be contributing to the development of ascites. This may include:  Maintaining a healthy diet: A low-sodium diet can help reduce the amount of fluid that accumulates in the body. It is important to avoid salty foods and to limit the amount of sodium in your diet to 2,000 milligrams per day or less. Your doctor or a nutritionist can help you plan a healthy, low-sodium diet.  Exercising regularly: Regular physical activity can help improve your overall health and reduce the risk of developing ascites.  Avoiding alcohol: Drinking alcohol can worsen liver damage and increase the risk of developing ascites. If you have cirrhosis, it is important to avoid alcohol altogether.  Managing other medical conditions: If you have other medical conditions such as high blood pressure or heart disease, it is important to manage these conditions properly as they can contribute to the development of ascites.  If you already have ascites, your doctor may recommend one or more of the following treatments:  Diuretics: These medications help your body get rid of excess fluid by increasing urine production. They can be taken orally or injected directly into a vein.  Paracentesis: This is a procedure in which a needle is inserted into the abdominal cavity to remove excess fluid.  Transjugular intrahepatic portosystemic shunt (TIPS): This is a procedure in which a small tube is inserted into the liver to help drain excess fluid from the abdominal cavity.  Liver transplant: In severe cases of cirrhosis, a liver transplant may be necessary to treat ascites.  It is important to work closely with your healthcare team to determine the best treatment plan for you. Proper management of ascites can help improve your quality of life and reduce the risk of complications. | Similar | 1 | 2 | 1 |
| 33. what is the life expectancy for patients with cirrhosis? | The life expectancy for patients with cirrhosis, a condition in which scar tissue replaces healthy liver tissue, can vary significantly depending on the cause of the cirrhosis, the severity of the disease, and the presence of complications such as ascites (accumulation of fluid in the abdomen), bleeding from esophageal varices (enlarged veins in the esophagus), and hepatic encephalopathy (a decline in brain function due to liver failure).  In general, the prognosis for patients with cirrhosis is poor, and the average life expectancy is about 5-10 years after diagnosis. However, with proper treatment and lifestyle changes, some patients with cirrhosis can live for many years and even achieve long-term survival.  Factors that can influence life expectancy in patients with cirrhosis include:  The underlying cause of the cirrhosis: Different types of cirrhosis have different prognoses. For example, cirrhosis caused by alcohol abuse is generally less severe and has a better prognosis than cirrhosis caused by viral hepatitis or autoimmune liver disease.  The stage of the disease: The severity of cirrhosis is usually classified into four stages (A, B, C, and D) based on the presence and severity of complications. Patients in the early stages of cirrhosis (A and B) generally have a better prognosis than those in the later stages (C and D).  The presence of complications: Complications such as ascites, bleeding from esophageal varices, and hepatic encephalopathy can significantly worsen the prognosis for patients with cirrhosis.  The presence of other medical conditions: Patients with cirrhosis who also have other medical conditions, such as diabetes or heart disease, may have a poorer prognosis.  It is important to note that life expectancy for patients with cirrhosis can vary widely, and some patients may live for many years with proper treatment and lifestyle changes. It is important for patients with cirrhosis to work closely with their healthcare team to manage their condition and minimize the risk of complications. | Similar | 3 | 2 | 3 |
| 34. Why do patients with cirrhosis develop red spots on chest/face? | Cirrhosis is a condition in which the liver becomes scarred and damaged, leading to a reduction in its ability to function properly. It is often the result of long-term alcohol abuse, but can also be caused by other factors such as viral hepatitis, autoimmune diseases, and fatty liver disease.  One of the complications of cirrhosis is the development of red spots on the skin, particularly on the chest and face. These spots, known as spider angiomas or angiomas, are small clusters of dilated blood vessels that appear as reddish or purplish lesions on the skin. They are often found in people with liver disease, and are thought to be caused by increased levels of estrogen in the body. High levels of estrogen can occur in people with cirrhosis due to the liver's decreased ability to metabolize and clear hormones from the body.  Spider angiomas are generally benign and do not cause any symptoms, but they may be a sign of underlying liver disease. If you have red spots on your chest or face and are concerned about your liver health, it is important to speak with a healthcare provider for further evaluation and treatment. | Similar | 1 | 1 | N/A |
| 35.  Can a patient with cirrhosis get liver cancer? How to prevent liver cancer in cirrhosis? | Yes, it is possible for a patient with cirrhosis to develop liver cancer. Cirrhosis is a condition in which the liver is damaged and scarred, and it is a major risk factor for the development of liver cancer.  There are several ways that people with cirrhosis can reduce their risk of developing liver cancer:  Avoiding alcohol: Excessive alcohol consumption is a major risk factor for the development of both cirrhosis and liver cancer. If you have cirrhosis, it is important to avoid alcohol or to consume it in moderation.  Getting vaccinated: Some viruses, such as hepatitis B and C, can cause cirrhosis and increase the risk of liver cancer. Getting vaccinated against these viruses can help protect against both conditions.  Controlling other risk factors: Other risk factors for liver cancer include being overweight or obese, having diabetes, and having high levels of iron in the blood. It is important to manage these risk factors through a healthy diet, exercise, and medication if necessary.  Regular monitoring: Regular monitoring by a healthcare provider can help identify early signs of liver cancer in people with cirrhosis. This may include imaging tests, blood tests, and liver biopsies.  It is important to note that while these measures may help reduce the risk of liver cancer in people with cirrhosis, they cannot completely eliminate the risk. If you have cirrhosis, it is important to follow the recommendations of your healthcare provider and to seek medical attention if you experience any unusual symptoms. | Similar | 1 | 2 | 1 |
| 36. What is the risk of liver cancer in patients with cirrhosis? | Cirrhosis is a serious and potentially life-threatening condition that can lead to an increased risk of developing liver cancer. The risk of developing liver cancer is higher in patients with cirrhosis than in those without the condition.  There are several factors that can increase the risk of developing liver cancer in patients with cirrhosis, including:  The severity of the cirrhosis: The more advanced the cirrhosis, the higher the risk of developing liver cancer.  The underlying cause of the cirrhosis: Certain causes of cirrhosis, such as chronic infection with the hepatitis B or C virus, are more likely to lead to liver cancer than others.  The presence of other risk factors: Other factors that can increase the risk of developing liver cancer in patients with cirrhosis include a family history of liver cancer, heavy alcohol consumption, and exposure to certain toxins or chemicals.  Overall, the risk of developing liver cancer in patients with cirrhosis is relatively low, but it is still important for these patients to undergo regular surveillance and screenings to detect any potential problems early. | Similar | 2 | 2 | N/A |
| **Diagnosis** | | | | | |
| 37. How is cirrhosis diagnosed? | Cirrhosis is a serious and potentially life-threatening condition that occurs when the liver is damaged and scarred over time. It is often the result of long-term alcohol abuse or chronic hepatitis, but it can also be caused by other conditions such as nonalcoholic fatty liver disease, autoimmune hepatitis, and primary biliary cholangitis.  There are several tests and procedures that may be used to diagnose cirrhosis, including:  Physical examination: During a physical examination, a healthcare provider will check for signs of liver damage, such as jaundice (yellowing of the skin and whites of the eyes), spider angiomas (small red spots on the skin), and ascites (abdominal swelling).  Blood tests: Blood tests can help to detect liver damage, such as an elevation in liver enzymes, and can also help to identify any underlying conditions that may be causing cirrhosis.  Liver function tests: These tests measure the level of certain substances in the blood, such as bilirubin and albumin, which can help to determine how well the liver is functioning.  Ultrasound: An ultrasound uses sound waves to create a detailed image of the liver and can help to identify scarring or other abnormalities in the liver.  CT scan or MRI: A CT scan or MRI can provide detailed images of the liver and can help to confirm the presence of cirrhosis.  Liver biopsy: A liver biopsy involves taking a small sample of liver tissue for examination under a microscope. This can help to confirm the diagnosis of cirrhosis and can also help to determine the extent and severity of the damage.  It's important to note that cirrhosis is a serious and potentially life-threatening condition that requires prompt treatment. If you have any symptoms that suggest you may have cirrhosis, it is important to see a healthcare provider for a proper diagnosis and treatment. | Similar | 1 | 2 | 2 |
| 38. What is a Child-Turcotte-Pugh score? | "The Child-Turcotte-Pugh (CTP) score is a system used to assess the severity of liver disease in patients with cirrhosis. It is a widely used tool that helps doctors determine the best course of treatment for patients with cirrhosis and helps predict the likelihood of survival.  The CTP score is calculated based on three factors:  The severity of liver dysfunction, as measured by the presence of certain clinical signs and symptoms, such as jaundice, ascites, and encephalopathy.  The degree of hepatic reserve, as measured by the patient's albumin level, prothrombin time, and bilirubin level.  The patient's functional status, as determined by their ability to perform activities of daily living.  Based on these three factors, the CTP score is calculated as follows:  Class A: Score 5-6 (mild liver disease)  Class B: Score 7-9 (moderate liver disease)  Class C: Score 10-15 (severe liver disease)  The CTP score is useful in helping doctors determine the best course of treatment for patients with cirrhosis and in predicting the likelihood of survival. It is important to note that the CTP score is only one tool that doctors use to assess liver disease, and it should be used in conjunction with other clinical and laboratory findings to get a complete picture of a patient's health. | Similar | 3 | 1 | 3 |
| 39. What is a MELD score? What is a MELD-Na score? | The Model for End-Stage Liver Disease (MELD) score is a system that is used to predict the likelihood of mortality within the next three months in patients with end-stage liver disease. It is calculated based on the values of three laboratory tests: serum bilirubin, serum creatinine, and international normalized ratio (INR). These tests are used to assess the severity of liver damage, kidney function, and blood clotting ability, respectively. The MELD score is calculated using the following formula:  MELD score = 3.78 ln(serum bilirubin (mg/dL)) + 11.2 ln(serum creatinine (mg/dL)) + 9.57 ln(INR) + 6.43  The MELD score ranges from 6 to 40, with a higher score indicating a higher risk of mortality. The MELD score is used to prioritize liver transplantation for patients with end-stage liver disease.  The MELD-Na score is a modified version of the MELD score that includes sodium level in the calculation. Sodium level is an important factor in the development of liver disease and can affect a patient's prognosis. The MELD-Na score is calculated using the following formula:  MELD-Na score = MELD score + 0.379 × ln(sodium level (mEq/L))  Like the MELD score, the MELD-Na score ranges from 6 to 40, with a higher score indicating a higher risk of mortality. The MELD-Na score is used to prioritize liver transplantation for patients with end-stage liver disease, particularly in cases where sodium levels are abnormal. | The Model for End-Stage Liver Disease (MELD) score is a numerical system used to predict the likelihood of mortality in patients with end-stage liver disease. It is used to prioritize the allocation of organs for transplantation, with higher MELD scores indicating a higher risk of death and a greater need for a transplant.  The MELD score is calculated using three laboratory values: the serum creatinine level, the serum bilirubin level, and the international normalized ratio (INR) for prothrombin time. These values are used to predict the likelihood of death within the next three months.  The MELD-Na score is similar to the MELD score, but it also includes a measure of sodium levels in the blood. The MELD-Na score is used to predict the likelihood of death in patients with end-stage liver disease who have ascites, or a build-up of fluid in the abdomen. It is used to prioritize the allocation of organs for transplantation in these patients, with higher MELD-Na scores indicating a higher risk of death and a greater need for a transplant. | 3,1 | 2,2 | 2,2 |
| **Treatment** | | | | | |
| 40. How is cirrhosis treated? | Cirrhosis is a serious and often progressive liver disease that is caused by long-term damage to the liver. It is characterized by scarring of the liver tissue, which can interfere with the organ's ability to function properly. There is no cure for cirrhosis, but treatment can help to slow the progression of the disease and improve symptoms.  Treatment for cirrhosis may include:  Lifestyle changes: Making lifestyle changes, such as quitting smoking and drinking alcohol, can help to slow the progression of cirrhosis.  Medications: Your doctor may prescribe medications to treat the underlying cause of cirrhosis, such as medications to control hepatitis or to reduce inflammation in the liver.  Nutrition: A healthy diet can help to support liver function and reduce the risk of complications. Your doctor may recommend a low-salt, low-fat, and high-carbohydrate diet.  Transplant: In severe cases of cirrhosis, a liver transplant may be necessary. A liver transplant involves surgically removing the damaged liver and replacing it with a healthy donor liver.  It is important to work closely with your doctor to develop a treatment plan that is right for you. Cirrhosis is a serious and potentially life-threatening condition, and it is important to seek medical treatment as soon as possible. | Similar | 1 | 2 | 2 |
| 41. Is there a cure for cirrhosis? | Cirrhosis is a chronic condition that can lead to serious health problems, including liver failure. While there is no cure for cirrhosis, treatment can help manage the symptoms and slow the progression of the disease. Early detection and treatment of the underlying cause of cirrhosis, such as chronic alcoholism or hepatitis, is the key to managing the condition.  Treatment options for cirrhosis may include:  Medications: There are several medications that can help manage the symptoms of cirrhosis and prevent further damage to the liver. These may include diuretics to reduce fluid accumulation, medications to control bleeding and blood pressure, and antibiotics to treat or prevent infections.  Lifestyle changes: Making healthy lifestyle choices, such as quitting smoking and reducing alcohol consumption, can help manage the symptoms of cirrhosis and slow the progression of the disease.  Nutrition: Eating a healthy, balanced diet can help improve overall health and provide the nutrients necessary for healing.  Surgery: In some cases, surgery may be necessary to treat complications of cirrhosis, such as a bleeding esophageal varix or hepatocellular carcinoma (liver cancer).  Liver transplant: In advanced cases of cirrhosis, a liver transplant may be the best option. A liver transplant involves replacing the damaged liver with a healthy one from a donor. While a liver transplant can be a life-saving treatment, it is a complex and risky procedure.  It is important to work closely with a healthcare provider to determine the best treatment plan for managing cirrhosis. | Similar | 1 | 2 | 1 |
| 42. When do doctors consider a liver transplant for cirrhosis? | Doctors usually consider a liver transplant for cirrhosis when the disease has progressed to a severe stage and the patient is experiencing significant symptoms or complications that are significantly impacting their quality of life. This may include ascites (buildup of fluid in the abdomen), hepatic encephalopathy (a brain disorder caused by liver failure), variceal bleeding (bleeding from enlarged veins in the esophagus or stomach), or severe malnutrition.  In general, liver transplantation is reserved for patients with end-stage liver disease who are not responding to other treatments and who have a good chance of surviving the procedure and recovering after the transplant. The decision to recommend a liver transplant is typically made by a team of specialists that includes liver disease experts, surgeons, and other healthcare professionals.  It's important to note that a liver transplant is a major surgery with significant risks and complications, and it's not suitable for everyone. It's also important to note that there is a shortage of available organs for transplant, so the process of getting a liver transplant can be very competitive. | Liver transplantation is typically considered for patients with cirrhosis when the condition has progressed to a stage known as end-stage liver disease (ESLD). This is a serious and advanced stage of liver damage, characterized by the presence of severe symptoms and complications that significantly impact a person's quality of life and survival.  There are several criteria that doctors use to determine if a liver transplant is appropriate for a patient with cirrhosis. These criteria may include:  The severity of the patient's liver disease: A patient with cirrhosis may be considered for a liver transplant if they have developed severe complications such as hepatic encephalopathy (a brain disorder caused by liver failure), ascites (abnormal accumulation of fluid in the abdomen), or variceal bleeding (bleeding from dilated veins in the esophagus or stomach).  The patient's overall health: A liver transplant candidate must be in good overall health and have no other serious medical conditions that would make them ineligible for transplant surgery.  The availability of a suitable donor organ: The availability of a suitable donor organ is a key factor in determining whether a patient with cirrhosis is a candidate for transplantation. The organ must be a good match for the patient and be in good condition.  The patient's willingness and ability to adhere to the post-transplant regimen: A liver transplant involves a significant commitment to ongoing medical care, including taking medications and following a special diet. The patient must be willing and able to adhere to this regimen in order to be considered for a transplant.  It's important to note that liver transplantation is a major surgery that carries risks and complications, and it is not always the best treatment option for every patient with cirrhosis. The decision to undergo a liver transplant is made on a case-by-case basis, after a thorough evaluation by a team of medical professionals. | 1 | 2 | 2 |
| 43. How much ascites can be removed each time for patients with cirrhosis? | Ascites, or the accumulation of fluid in the abdominal cavity, can be a common complication of cirrhosis, a type of chronic liver disease. The amount of ascites that can be removed during a procedure, such as paracentesis, will depend on various factors including the patient's overall health, the underlying cause of the ascites, and the presence of any other complications.  In general, it is generally safe to remove up to 5 liters of ascites at a time, although some patients may be able to tolerate more. The amount of ascites that can be removed in a single procedure may also be limited by the size of the patient's abdominal cavity and the presence of any adhesions or other abnormalities.  It is important to note that paracentesis, or the removal of ascites, is typically only used to relieve symptoms and is not a cure for the underlying condition. Other treatments, such as medications and lifestyle changes, may be necessary to manage the cirrhosis and prevent further accumulation of ascites. | Ascites, or the accumulation of fluid in the abdomen, is a common complication of cirrhosis, a chronic liver disease. The amount of ascites that can be removed during a procedure called paracentesis will depend on several factors, including the underlying cause of the ascites, the patient's overall health, and the presence of any other medical conditions.  In general, the maximum amount of fluid that can be safely removed during a single paracentesis procedure is usually between 5 and 25 liters, depending on the patient's size and condition. However, removing large amounts of fluid at once can be challenging and may lead to complications such as hypovolemia (low blood volume) or electrolyte imbalances. Therefore, it is generally recommended to remove smaller amounts of fluid at a time, with the goal of gradually reducing the overall amount of ascites.  It is important to note that paracentesis is not a cure for ascites, and the fluid will usually return unless the underlying cause of the ascites is treated. Therefore, patients with cirrhosis and ascites may need to undergo repeat paracentesis procedures or receive other forms of treatment, such as medications or liver transplantation, to manage their condition. | 1,3 | 2,3 | 1,2 |
| 44. Should a patient with cirrhosis get albumin with a paracentesis? | It is generally recommended that patients with cirrhosis who are undergoing a paracentesis procedure receive albumin replacement. This is because patients with cirrhosis often have low levels of albumin due to liver dysfunction, and losing large volumes of fluid during a paracentesis can further decrease their albumin levels. Low albumin levels can lead to complications such as edema (fluid accumulation in the tissue) and ascites (fluid accumulation in the abdominal cavity).  Albumin replacement can help to maintain or restore normal albumin levels and prevent or treat these complications. It is usually given intravenously after the paracentesis procedure. The dose and duration of albumin replacement will depend on the patient's individual circumstances and should be determined by a healthcare provider.  It is important to note that albumin replacement is not always necessary in every patient with cirrhosis undergoing a paracentesis. The decision to use albumin replacement should be made on a case-by-case basis, taking into account the patient's overall clinical status, the amount of fluid removed during the paracentesis, and any other relevant factors. | Similar | 1 | 2 | 2 |
| 45. How to treat ascites in patients with cirrhosis that is not responsive to high dose waterpills? | Ascites, or the accumulation of fluid in the abdomen, is a common complication of cirrhosis. It can be caused by a variety of factors, including high blood pressure in the veins that carry blood from the intestines, liver, and spleen to the heart (hepatic venous pressure gradient, or HVPG), kidney problems, and high levels of aldosterone, a hormone that regulates electrolyte balance and blood pressure.  Treatment for ascites in patients with cirrhosis typically involves a combination of medications and lifestyle changes. If high-dose diuretics (water pills) are not sufficient to control ascites, other options may include:  Increasing the dose of diuretics or switching to a different type of diuretic: Your healthcare provider may recommend increasing the dose of your current diuretic or switching to a different type of diuretic, such as spironolactone or eplerenone, which can help reduce fluid accumulation.  Adding a vasoconstrictor: Vasoconstrictors, such as midodrine or octreotide, can help constrict blood vessels and reduce fluid accumulation.  Using albumin infusions: Albumin is a protein that helps maintain fluid balance in the body. Infusions of albumin can help prevent or reduce the risk of developing ascites.  Paracentesis: If the ascites is severe and not responding to other treatments, your healthcare provider may recommend a procedure called paracentesis to remove the excess fluid from your abdomen.  Liver transplant: In some cases, liver transplantation may be recommended for patients with cirrhosis and refractory ascites.  It's important to work closely with your healthcare provider to determine the best treatment approach for your specific situation. | Ascites, or the accumulation of fluid in the abdomen, is a common complication of cirrhosis, a condition in which scar tissue replaces healthy liver tissue and impairs its function. If ascites is not responsive to high-dose diuretics (also known as water pills), there are several other treatment options that may be considered.  Sodium restriction: Limiting sodium intake can help to reduce the amount of fluid in the body. This may involve following a low-sodium diet and avoiding salty foods and drinks.  Paracentesis: This procedure involves removing fluid from the abdomen using a needle or catheter. It can provide relief from the symptoms of ascites, but it is typically not a long-term solution.  Transjugular intrahepatic portosystemic shunt (TIPS): This procedure involves creating a connection between the portal vein and the hepatic vein in the liver, allowing blood to bypass the damaged liver tissue. This can help to reduce pressure in the portal vein and decrease the amount of fluid that accumulates in the abdomen.  Liver transplant: In severe cases of cirrhosis, a liver transplant may be necessary. This involves replacing the damaged liver with a healthy one from a donor.  It is important to work closely with a healthcare provider to determine the most appropriate treatment for ascites in patients with cirrhosis. In some cases, a combination of these approaches may be necessary to manage the condition effectively. | 3, 1 | 2,1 | 3,2 |
| 46. What are the side effects of diuretics in cirrhosis? | Diuretics, also known as water pills, are medications that increase the amount of urine produced by the kidneys and help to remove excess fluid from the body. They are often used in the treatment of cirrhosis, a condition in which the liver is damaged and scarred, to help reduce fluid buildup in the abdomen and legs.  There are several potential side effects of diuretics in people with cirrhosis, including:  Electrolyte imbalances: Diuretics can cause imbalances in electrolytes such as potassium, sodium, and chloride, which are essential for proper body function. Low potassium levels can cause muscle weakness and cramping, while low sodium levels can cause fatigue and confusion.  Dehydration: Diuretics can cause dehydration, especially if they are taken without adequate fluid intake. This can lead to dizziness, fatigue, and muscle cramps.  Kidney damage: Diuretics can cause kidney damage in some people, especially those with pre-existing kidney problems or those taking high doses of the medication.  Low blood pressure: Diuretics can lower blood pressure, which can cause dizziness or fainting when standing up too quickly.  Hypotension: Diuretics can cause hypotension, or low blood pressure, which can lead to symptoms such as dizziness, lightheadedness, and fainting.  It is important to talk to your doctor about the potential side effects of diuretics and to follow their instructions for taking the medication. If you experience any side effects, it is important to report them to your doctor so that they can be properly managed. | Similar | 1 | 2 | 1 |
| 47. Can transjugular intrahepatic portosystemic shunt helps with ascites? | Transjugular intrahepatic portosystemic shunt (TIPS) is a procedure that involves creating a connection between the portal vein and the hepatic vein within the liver. This connection, or shunt, can help to reduce the pressure in the portal vein, which can in turn help to reduce the accumulation of fluid in the abdomen, or ascites.  Ascites is a common complication of liver cirrhosis and other conditions that affect the liver and can cause fluid to build up in the abdomen. TIPS can be used as a treatment option for ascites in people who have not responded to other treatments, such as diuretics or paracentesis (removal of fluid from the abdomen using a needle).  TIPS is a minimally invasive procedure that is typically performed using imaging guidance, such as ultrasound or computed tomography (CT). The procedure involves inserting a thin tube, or catheter, through a small incision in the neck or groin and threading it through the blood vessels to the liver. Once the catheter is in place, a stent is inserted through the catheter to create the shunt.  TIPS can be effective at reducing ascites in many people, but it is not a cure for the underlying cause of the fluid accumulation. It may be necessary to repeat the procedure if the ascites returns or to consider other treatment options, such as liver transplantation, if the underlying liver disease progresses. | Similar | 1 | 2 | 2 |
| 48. Are there any side effects of transjugular intrahepatic portosystemic shunt? | Transjugular intrahepatic portosystemic shunt (TIPS) is a procedure that is used to treat portal hypertension, which is a condition in which the blood pressure in the portal vein (which carries blood from the intestines to the liver) is higher than normal. The procedure involves creating a shunt, or small passage, between the portal vein and the hepatic vein (which carries blood from the liver to the heart).  As with any medical procedure, there are potential risks and side effects associated with TIPS. Some of the possible side effects of TIPS include:  Bleeding: TIPS involves making a small incision in the skin and then inserting a catheter into a vein. There is a risk of bleeding during or after the procedure.  Infection: There is a risk of infection at the incision site or at the insertion site of the catheter.  Reactivation of hepatitis: TIPS may cause hepatitis to reactivate in some people who have a history of hepatitis.  Liver failure: TIPS may cause liver failure in some cases.  Portal vein thrombosis: TIPS may cause a blood clot to form in the portal vein, which can be life-threatening.  Stenosis: TIPS may cause the shunt to become narrowed or blocked over time, which can lead to portal hypertension.  Abdominal pain: Some people may experience abdominal pain after the procedure.  It's important to discuss the potential risks and benefits of TIPS with a healthcare provider before the procedure. | Transjugular intrahepatic portosystemic shunt (TIPS) is a procedure used to treat portal hypertension, which is a condition that occurs when the blood pressure in the portal vein (which carries blood from the digestive organs to the liver) is too high. The procedure involves creating a shunt, or small channel, between the portal vein and the hepatic vein (which carries blood from the liver to the heart).  TIPS can be effective in reducing portal hypertension and relieving symptoms such as abdominal swelling and ascites (fluid accumulation in the abdomen). However, like any medical procedure, TIPS carries some risks and potential side effects. These can include:  Bleeding: There is a risk of bleeding during the procedure, which may require a blood transfusion.  Infection: There is a risk of infection at the site of the shunt.  Hepatic encephalopathy: TIPS may cause hepatic encephalopathy, a condition in which toxins build up in the brain due to liver dysfunction. Symptoms may include confusion, tremors, and difficulty speaking.  Shunt stenosis: The shunt may become narrowed, which can decrease its effectiveness. This may require further intervention.  Renal failure: TIPS may cause kidney failure in some cases.  Shunt dysfunction: The shunt may become blocked or may not work properly, which can lead to an increased risk of bleeding or other complications.  It's important to discuss the potential risks and benefits of TIPS with a healthcare provider before undergoing the procedure. | 2,2 | 2,2 | N/A |
| 49. What can be done to treat nonalcoholic steatohepatitis-related cirrhosis? | Nonalcoholic steatohepatitis (NASH) is a type of liver disease characterized by the accumulation of fat in the liver and inflammation, which can eventually lead to cirrhosis, a severe form of liver damage. There is no specific treatment for NASH-related cirrhosis, but there are several measures that can be taken to slow the progression of the disease and improve symptoms.  The most important step in managing NASH-related cirrhosis is to address the underlying cause of the liver damage, which is often related to unhealthy lifestyle factors such as being overweight or obese, having high levels of cholesterol or triglycerides in the blood, or having uncontrolled diabetes.  Treatment may involve a combination of the following measures:  Lifestyle changes: Making healthy lifestyle changes, such as losing weight, exercising regularly, and eating a healthy diet, can help improve liver function and slow the progression of NASH.  Medications: Certain medications, such as statins, can be used to lower cholesterol and triglyceride levels, which may help reduce the amount of fat in the liver.  Vitamin supplements: Some studies have shown that taking supplements of antioxidants, such as vitamin E, may help reduce liver inflammation and improve liver function in people with NASH.  Liver transplant: In advanced cases of NASH-related cirrhosis, a liver transplant may be necessary. This involves replacing the damaged liver with a healthy one from a donor.  It's important to work closely with a healthcare provider to determine the best course of treatment for NASH-related cirrhosis. Prompt treatment is crucial to prevent further liver damage and potentially life-threatening complications. | Similar | 1 | 3 | 1 |
| 50. why should beta blockers be taken for cirrhosis? | Beta blockers are medications that block the effects of the hormone adrenaline on the body's cardiovascular system, which can help lower blood pressure and reduce the risk of heart problems. In people with cirrhosis, a type of liver disease, beta blockers may be used to help reduce the risk of complications such as variceal bleeding, a type of bleeding that occurs when the blood vessels in the esophagus or stomach become dilated and rupture.  Cirrhosis can cause an increase in blood pressure in the veins leading to the liver, which can lead to the development of varices. Beta blockers can help reduce the blood pressure in these veins and decrease the risk of variceal bleeding. They may also be used to help reduce the risk of other complications of cirrhosis, such as heart failure and portal hypertension.  It is important to note that beta blockers should be used with caution in people with cirrhosis, as they can cause side effects such as fatigue and impaired breathing. It is important to discuss the use of beta blockers with a healthcare provider, who can determine the appropriate dosage and monitoring for individual patients. | Similar | 1 | 2 | 2 |
| 51. How long is Rifaximin taken for hepatic encephalopathy? | Rifaximin is a medication that is used to treat hepatic encephalopathy, a condition that can occur in people with liver disease or cirrhosis. The usual recommended dose of rifaximin for the treatment of hepatic encephalopathy is 550 mg taken orally twice daily for 7 to 14 days. The duration of treatment with rifaximin will depend on the severity of the hepatic encephalopathy and the response to treatment. Your healthcare provider will determine the appropriate duration of treatment for you based on your individual needs. It is important to follow your healthcare provider's instructions and take rifaximin as directed. Do not stop taking rifaximin or change your dose without first talking to your healthcare provider. | Rifaximin is a medication that is used to treat hepatic encephalopathy, which is a condition that occurs when there is a buildup of toxins in the blood that can lead to changes in mental status and other neurological symptoms. Rifaximin is usually taken by mouth, and the usual dosage is 200 mg to 400 mg three times a day. The duration of treatment with rifaximin will depend on the severity of the hepatic encephalopathy and the response to treatment. It is important to follow the dosing instructions provided by your healthcare provider and to continue taking the medication as prescribed, even if you start to feel better, to help prevent the hepatic encephalopathy from recurring. | 3,3 | 3,3 | N/A |
| 52. What are the benefits and risk of taking rifaximin in patients with cirrhosis? | Rifaximin is an antibiotic that is commonly used to treat certain types of gastrointestinal infections and is sometimes used in the treatment of hepatic encephalopathy in patients with cirrhosis.  The benefits of taking rifaximin in patients with cirrhosis include:  Reducing the risk of recurrent episodes of hepatic encephalopathy, which is a condition that can cause confusion, difficulty with coordination and balance, and even coma in severe cases.  Improving quality of life for patients with cirrhosis by reducing symptoms associated with hepatic encephalopathy.  Decreasing the need for hospitalizations and other medical interventions in patients with cirrhosis.  Some of the potential risks and side effects of taking rifaximin in patients with cirrhosis include:  Diarrhea: Rifaximin can cause diarrhea, which can be a particular concern for patients with cirrhosis, as they may be more prone to dehydration.  Allergic reactions: Rifaximin can cause allergic reactions in some people, including rash, itching, and difficulty breathing.  Other gastrointestinal side effects: Rifaximin can also cause other gastrointestinal side effects, such as nausea, abdominal pain, and bloating.  Antibiotic resistance: The use of antibiotics, including rifaximin, can lead to the development of antibiotic-resistant bacteria. This can be a concern for patients with cirrhosis, as they may be more vulnerable to infection.  It is important to discuss the potential risks and benefits of taking rifaximin with your healthcare provider, as well as any other medications you are taking, before starting treatment. Your healthcare provider will be able to determine the most appropriate treatment for your specific situation. | Rifaximin is a broad-spectrum antibiotic that is commonly used to treat bacterial infections in the gut. In patients with cirrhosis, rifaximin may be used to treat infections or to reduce the risk of gastrointestinal bleeding.  The potential benefits of taking rifaximin in patients with cirrhosis include:  Reducing the risk of gastrointestinal bleeding: Cirrhosis is a condition in which the liver is scarred and damaged, leading to impaired function. One of the complications of cirrhosis is an increased risk of gastrointestinal bleeding, which can be life-threatening. Rifaximin may help reduce this risk by decreasing the amount of bacteria in the gut, which can contribute to the development of bleeding.  Treating infections: Cirrhosis can weaken the immune system, making patients more prone to infections. Rifaximin may be effective in treating infections caused by bacteria in the gut, such as traveler's diarrhea or diarrhea associated with antibiotic use.  However, there are also risks associated with taking rifaximin in patients with cirrhosis. These may include:  Drug interactions: Rifaximin may interact with other medications that a patient with cirrhosis is taking, such as blood thinners or medications for high blood pressure. It is important for patients to inform their healthcare provider about all medications they are taking to avoid potential drug interactions.  Allergic reactions: Some patients may have an allergic reaction to rifaximin, which can range from mild skin rash to a life-threatening reaction called anaphylaxis.  Antibiotic resistance: Overuse or improper use of antibiotics, including rifaximin, can lead to the development of antibiotic-resistant bacteria. This can make it more difficult to treat infections in the future.  In summary, the benefits of taking rifaximin in patients with cirrhosis may include reducing the risk of gastrointestinal bleeding and treating infections. However, there are also risks to consider, including drug interactions, allergic reactions, and the potential for antibiotic resistance. It is important for patients to discuss the risks and benefits of rifaximin with their healthcare provider before starting treatment. | 1,3 | 3,3 | 3,3 |
| 53. What are complications of paracentesis in cirrhosis? | Paracentesis is a procedure in which a needle or catheter is inserted into the abdomen to remove excess fluid that has accumulated there, a condition called ascites. In people with cirrhosis, a liver condition that is often caused by chronic alcoholism or viral hepatitis, paracentesis can be a useful treatment to relieve abdominal discomfort and improve quality of life. However, there are several potential complications that can occur during or after the procedure, including:  Infection: The risk of infection is generally low, but there is a risk of infection at the insertion site or the possibility of introducing bacteria into the abdomen.  Bleeding: There is a risk of bleeding during the procedure, especially in people with bleeding disorders or who are taking blood-thinning medications.  Perforation: There is a risk of perforation, or puncturing, of the abdominal wall or other organs during the procedure.  Hypovolemia: Removing a large amount of fluid from the abdomen can lead to low blood volume (hypovolemia), which can cause lightheadedness, dizziness, and fainting.  Recurrent ascites: In some cases, the fluid may return after the procedure, and additional paracentesis may be needed.  Electrolyte imbalances: Removing fluid from the abdomen can also lead to imbalances in electrolytes, such as sodium and potassium, which can cause muscle weakness and other symptoms.  Overall, the risks of paracentesis are generally low, and the procedure is generally well tolerated. However, it is important to discuss the potential risks and benefits of the procedure with a healthcare provider before deciding whether it is appropriate for you. | Similar | 1 | 2 | 1 |
| 54. What should be done to prevent complications of paracentesis in cirrhosis patients? | Paracentesis is a procedure in which fluid is removed from the abdominal cavity using a needle or a small tube. It is often performed in patients with cirrhosis, a chronic liver disease, to remove excess fluid that has accumulated in the abdomen, a condition known as ascites.  There are several steps that can be taken to prevent complications during and after paracentesis in patients with cirrhosis:  Use proper sterile technique: It is important to follow proper sterilization techniques to reduce the risk of infection.  Administer antibiotics: Antibiotics may be given before and after the procedure to reduce the risk of infection.  Monitor fluid balance: It is important to closely monitor the patient's fluid balance to ensure that they do not become dehydrated or overhydrated after the procedure.  Monitor for bleeding: Bleeding can occur during or after paracentesis. It is important to closely monitor the patient for signs of bleeding, such as blood in the drainage fluid or a drop in blood pressure.  Use a small needle: Using a smaller needle for the procedure may reduce the risk of complications, such as bleeding or infection.  Avoid over-draining: It is important to avoid removing too much fluid during the procedure to prevent complications, such as low blood pressure or dehydration.  Avoid large volumes of fluid: Removing large volumes of fluid at once can cause complications, such as low blood pressure or electrolyte imbalances. It is generally recommended to remove smaller volumes of fluid at a time.  It is important to follow the guidelines and recommendations of the healthcare provider to prevent complications during and after paracentesis in patients with cirrhosis. | Similar | 2 | 3 | 3 |
| 55. Does eating salt help increase blood sodium levels in cirrhosis? | In cirrhosis, the liver is damaged and may not be able to function properly. This can lead to a variety of complications, including an imbalance of electrolytes, including sodium, in the body. Consuming more salt can help increase blood sodium levels, but it is important to be cautious when doing so, as excessive salt intake can be harmful to people with cirrhosis.  In cirrhosis, the liver is unable to remove excess water and electrolytes from the body as efficiently as it should. As a result, people with cirrhosis may develop a condition called hyponatremia, which is characterized by low levels of sodium in the blood. Low sodium levels can cause symptoms such as fatigue, weakness, muscle cramps, and dizziness.  If you have cirrhosis and are experiencing low blood sodium levels, it is important to talk to your doctor about the best way to manage your condition. Your doctor may recommend a low-sodium diet or may prescribe medications to help manage your electrolyte levels. In some cases, your doctor may recommend increasing your salt intake, but this should be done under medical supervision to ensure that you are not consuming too much salt.  In general, it is important to maintain a balanced diet and to consume a healthy amount of salt. Too much salt can lead to high blood pressure, which can be harmful to people with cirrhosis. It is always best to consult with a healthcare professional before making any changes to your diet or medication regimen. | Similar | 3 | 3 | N/A |
| **Lifestyle** | | | | | |
| 56. Can cirrhosis patients still drink alcohol? | It is generally not recommended for people with cirrhosis to drink alcohol. Cirrhosis is a serious condition in which the liver is severely damaged, and alcohol can further damage the liver and worsen the symptoms of cirrhosis.  The liver is responsible for breaking down and removing toxins from the body, and alcohol is a toxic substance that puts a lot of strain on the liver. In people with cirrhosis, the liver is already impaired and may not be able to effectively process alcohol, which can lead to further damage and complications.  In addition to damaging the liver, alcohol can also interfere with medications that people with cirrhosis may be taking and can worsen other health conditions that may be present.  It is important for people with cirrhosis to follow the recommendations of their healthcare provider and to avoid alcohol in order to protect their liver and maintain their overall health. | Similar | 1 | 1 | N/A |
| 57. What pain medications are safe in cirrhosis? | Cirrhosis is a condition in which the liver is severely damaged and scarred, and it can cause a range of symptoms including abdominal pain, fatigue, and weakness. The safe use of pain medications in people with cirrhosis can be challenging because the liver plays a crucial role in metabolizing and eliminating many medications from the body. As a result, people with cirrhosis may be at increased risk of side effects or adverse reactions to certain medications.  It is important to consult a healthcare professional before starting any new medications, including pain medications, if you have cirrhosis. Your healthcare provider will consider your specific situation and medical history, as well as the potential risks and benefits of different treatment options. They may recommend a pain medication that is safe for use in people with cirrhosis or recommend a different treatment approach.  In general, non-steroidal anti-inflammatory drugs (NSAIDs) such as ibuprofen and naproxen are not recommended for use in people with cirrhosis because they can cause gastrointestinal bleeding and worsen liver function. Acetaminophen (also known as paracetamol) is generally considered to be safer for use in people with cirrhosis, although it is important to use it at the lowest effective dose for the shortest possible duration and to avoid taking more than the recommended maximum daily dose.  Other options for managing pain in people with cirrhosis may include medications such as opioids (e.g., morphine, oxycodone), gabapentin, or tricyclic antidepressants. Again, it is important to consult a healthcare professional to determine the most appropriate treatment approach for you. They will consider factors such as the severity of your pain, the underlying cause of your pain, and any other medical conditions you may have. | Similar | 3 | 2 | 2 |
| 58. Can I take Tylenol in cirrhosis? How much Tylenol is allowed in patients with cirrhosis? | It is generally not recommended to take Tylenol (acetaminophen) if you have cirrhosis. This is because Tylenol is metabolized by the liver, and the liver may not function properly in people with cirrhosis. If the liver is not able to properly metabolize Tylenol, it can accumulate in the body and cause serious side effects, including liver damage.  In people with cirrhosis, the recommended daily maximum dose of Tylenol is generally considered to be 2 grams (2000 milligrams) or less. This is because the risk of liver damage increases as the dose of Tylenol increases. It is important to talk to your doctor or pharmacist before taking any medication, including Tylenol, if you have cirrhosis. They can help you determine the appropriate dose and frequency of Tylenol based on your specific circumstances. | Similar | 1 | 1 | N/A |
| 59. What medications should be avoided for cirrhosis patients? | Cirrhosis is a serious condition that affects the liver and can lead to complications such as hepatic encephalopathy, bleeding, and infections. It is important for people with cirrhosis to be careful about the medications they take, as some medications can be harmful to the liver or may interact with other medications that they are taking.  Some medications that should be avoided in people with cirrhosis include:  Acetaminophen (Tylenol): This is a commonly used pain reliever that is generally safe for most people, but it can be harmful to the liver in large amounts or if it is taken for a long period of time. People with cirrhosis should avoid taking acetaminophen or should use it with caution and only as directed by their healthcare provider.  Nonsteroidal anti-inflammatory drugs (NSAIDs): These medications, which include ibuprofen and naproxen, can cause stomach bleeding and ulcers, and they may also increase the risk of bleeding in people with cirrhosis.  Corticosteroids: These medications, which are used to treat a variety of conditions, can cause fluid retention and increase the risk of bleeding in people with cirrhosis.  Amiodarone: This medication, which is used to treat certain heart conditions, can be harmful to the liver and should be avoided in people with cirrhosis.  It is important to talk to your healthcare provider about all of the medications you are taking, including over-the-counter medications and supplements, to make sure they are safe for you to use. Your healthcare provider can help you determine which medications are safe for you to take and can recommend alternative medications if necessary. | Cirrhosis is a serious condition in which the liver becomes scarred and is unable to function properly. It is important for individuals with cirrhosis to be careful about the medications they take, as some medications can be harmful to the liver or may interact with other medications in a way that could be harmful.  Some common medications that may be harmful for individuals with cirrhosis include:  Nonsteroidal anti-inflammatory drugs (NSAIDs): These include over-the-counter painkillers such as ibuprofen (Advil) and naproxen (Aleve). NSAIDs can cause gastrointestinal bleeding and may be harmful to the liver.  Acetaminophen (Tylenol): While acetaminophen is generally considered safe when taken at recommended doses, it can be harmful to the liver when taken in large amounts or when combined with alcohol.  Statins: These medications, which are used to lower cholesterol, can cause muscle damage and may be harmful to the liver.  Antifungal medications: Some antifungal medications, such as ketoconazole, can be harmful to the liver.  It is important for individuals with cirrhosis to speak with their healthcare provider about all medications they are taking, including over-the-counter medications and supplements. The healthcare provider can help determine which medications are safe to take and which should be avoided. | 3,1 | 3,3 | 3,1 |
| 60. What diet is good for cirrhosis? | Cirrhosis is a serious condition in which scar tissue replaces healthy liver tissue, leading to a loss of function in the liver. It is usually caused by long-term damage from conditions such as alcohol abuse, hepatitis, and nonalcoholic fatty liver disease.  A healthy diet can help manage the symptoms of cirrhosis and support overall health. It is important to follow the recommendations of a healthcare professional and a registered dietitian, as the specific dietary needs may vary depending on the individual's overall health and the stage of their cirrhosis.  Generally, a diet for cirrhosis should include:  Adequate calories and protein to help maintain strength and prevent malnutrition.  Foods that are high in nutrients but low in sodium, as a high-sodium diet can worsen swelling in the abdomen and legs. This may include fresh fruits and vegetables, whole grains, and lean proteins such as chicken, fish, and beans.  Limited amounts of fat, as the liver may have difficulty processing large amounts of fat.  Adequate fluids to prevent dehydration and help flush toxins out of the body.  It is also important to avoid alcohol, as it can further damage the liver.  It is important to note that a healthy diet is just one part of the treatment plan for cirrhosis. It is crucial to follow the recommendations of a healthcare professional and to take any prescribed medications as directed. | Similar | 1 | 2 | 1 |
| 61. Will taking supplements help with cirrhosis? | Cirrhosis is a serious condition in which scar tissue replaces healthy liver tissue, leading to impaired liver function. While certain supplements may be helpful in managing some of the symptoms associated with cirrhosis, it is important to note that supplements are not a substitute for medical treatment and should not be used in place of prescribed medications.  If you have cirrhosis, it is important to work closely with your healthcare provider to develop a treatment plan that is appropriate for your specific needs. This may include medications, lifestyle changes, and other therapies to help manage your condition and prevent further liver damage.  Some supplements that may be helpful in managing symptoms associated with cirrhosis include:  Vitamin K: This vitamin is important for blood clotting and may be recommended for people with cirrhosis who are at risk of bleeding due to low levels of clotting factors produced by the liver.  Folic acid: This vitamin is important for cell growth and may be recommended for people with cirrhosis to help prevent anemia.  L-carnitine: This amino acid may be helpful in improving muscle strength and endurance in people with cirrhosis.  It is important to talk to your healthcare provider before taking any supplements, as they may interact with other medications you are taking or have potential side effects. Your healthcare provider can help you determine the best course of treatment for your specific needs. | Similar | 1 | 3 | 3 |
| 62. Can cirrhosis affect sexual function? | Yes, cirrhosis can affect sexual function in both men and women. In men, cirrhosis can lead to a decrease in testosterone production, which can cause reduced sex drive and erectile dysfunction. In women, cirrhosis can lead to a decrease in estrogen production, which can cause menopausal symptoms such as vaginal dryness and reduced sex drive.  Cirrhosis can also lead to physical changes that can affect sexual function. For example, people with cirrhosis may experience swelling in the legs and abdomen, which can make physical activity, including sexual activity, more difficult. In addition, cirrhosis can cause fatigue and muscle weakness, which can also affect sexual function.  It is important to talk to a healthcare provider if you are experiencing sexual dysfunction as a result of cirrhosis. There may be treatments available to help manage these symptoms. | Similar | 1 | 2 | 1 |
| 63. Can cirrhosis affect fertility? | Yes, cirrhosis can affect fertility in both men and women. In men, cirrhosis can cause low testosterone levels, which can lead to decreased fertility. In women, cirrhosis can cause irregular periods, which can make it difficult to become pregnant. Additionally, cirrhosis can cause scarring of the reproductive organs, which can also affect fertility.  It's important to note that the effects of cirrhosis on fertility can vary depending on the severity of the liver damage and the underlying cause of the cirrhosis. If you have cirrhosis and are concerned about your fertility, it's a good idea to discuss your concerns with a healthcare professional. They can provide more specific information and guidance based on your individual situation. | Similar | 3 | 2 | 3 |
| 64. Is exercise safe in cirrhosis? | Exercise can be safe and beneficial for people with cirrhosis, a condition in which scar tissue replaces healthy liver tissue, causing the liver to lose its ability to function properly. However, it is important for people with cirrhosis to talk to their healthcare provider before starting or making changes to an exercise routine.  Exercise can help improve physical function and quality of life in people with cirrhosis, but it is important to start slowly and gradually increase the intensity and duration of exercise as tolerated. It is also important to monitor for any signs of worsening liver function or other complications during exercise.  People with cirrhosis may experience fatigue, muscle weakness, and difficulty breathing, which can make exercise more challenging. It is important to listen to your body and stop exercising if you experience any discomfort or difficulty. It may also be helpful to work with a physical therapist or exercise specialist who can help develop a safe and effective exercise plan.  In general, it is important for people with cirrhosis to avoid activities that are excessively strenuous or involve heavy lifting, as these activities can put extra strain on the liver and other organs. Low-impact activities such as walking, swimming, and cycling may be safer and easier to tolerate.  Again, it is important to consult with a healthcare provider before starting or changing an exercise routine if you have cirrhosis. They can help you develop a safe and effective exercise plan that is tailored to your specific needs and abilities. | Similar | 1 | 2 | 1 |
| 65. What can be done to improve muscle loss in cirrhosis? | Cirrhosis is a condition in which the liver becomes scarred and damaged, leading to impaired function and a range of symptoms including muscle loss. There are several potential strategies that may help to improve muscle loss in people with cirrhosis:  Exercise: Exercise can help to build and maintain muscle mass. It is important to consult with a healthcare professional before starting any exercise program, as people with cirrhosis may have limitations on the types and intensity of exercise they can do.  Nutritional support: Proper nutrition is important for maintaining muscle mass. People with cirrhosis may need to consume a higher-calorie diet to support muscle growth and repair. A registered dietitian can help develop a nutrition plan that meets an individual's needs.  Anabolic steroids: Anabolic steroids are medications that can help to stimulate muscle growth. They may be prescribed to people with cirrhosis who have significant muscle loss. However, these medications can have serious side effects and should be used with caution.  Physical therapy: Physical therapy can help to strengthen muscles and improve mobility. A physical therapist can work with an individual to develop a customized exercise plan.  Liver transplant: In some cases, a liver transplant may be necessary to improve muscle loss in people with cirrhosis. This is typically reserved for those with advanced liver disease who have not responded to other treatments.  It is important to work closely with a healthcare team to determine the best course of treatment for muscle loss in cirrhosis. | Similar | 1 | 3 | 3 |
| 66. What can be done to improve fatigue in cirrhosis? | Fatigue is a common symptom in individuals with cirrhosis, a progressive liver disease that can lead to serious complications such as liver failure. There are several things that can be done to help improve fatigue in people with cirrhosis:  Get enough rest: It is important to get enough sleep and rest to help manage fatigue. It may be helpful to create a sleep schedule and stick to it.  Exercise regularly: Exercise can help improve energy levels and reduce fatigue. However, it is important to talk to a healthcare provider before starting any new exercise program.  Eat a healthy diet: A healthy diet that is rich in nutrients can help improve energy levels and reduce fatigue. It is important to eat a balanced diet that includes plenty of fruits, vegetables, whole grains, and protein.  Avoid alcohol and drugs: Alcohol and drugs can worsen fatigue and other symptoms of cirrhosis. It is important to avoid these substances to improve overall health.  Manage stress: Stress can worsen fatigue and other symptoms of cirrhosis. It may be helpful to try stress-management techniques such as relaxation techniques, meditation, or exercise.  Get support: It can be helpful to connect with others who are also dealing with cirrhosis. Support groups can provide a sense of community and can be a source of support and encouragement.  Talk to a healthcare provider: It is important to speak with a healthcare provider about fatigue and other symptoms of cirrhosis. They can help determine the cause of fatigue and recommend treatment options. | Similar | 1 | 2 | 1 |
| 67. What can be done to improve pruritus in cirrhosis? | Pruritus, or itching, is a common symptom in cirrhosis, a late stage of liver disease. It can be caused by a variety of factors, including increased levels of certain chemicals in the blood, dry skin, and changes in nerve function.  There are several strategies that may be helpful in improving pruritus in cirrhosis:  Hydration: Keeping the skin well-moisturized can help alleviate itching. Using a humidifier and taking frequent lukewarm baths or showers can help.  Avoiding triggers: Certain substances and activities may worsen itching. These can include hot water, scratchy clothing, and certain medications. Avoiding these triggers can help improve pruritus.  Medications: There are several medications that may be helpful in reducing itching in cirrhosis. These include antihistamines, opioids, and medications that block the actions of certain chemicals in the body.  Lifestyle changes: Making lifestyle changes, such as quitting smoking and reducing alcohol consumption, may help improve pruritus in cirrhosis.  Light therapy: Phototherapy, or light therapy, may be helpful in reducing itching in some people with cirrhosis.  It is important to speak with a healthcare professional about the best treatment approach for pruritus in cirrhosis. | Similar | 1 | 2 | 2 |
| 68. What can be done to improve brain fog in cirrhosis? | Brain fog, or cognitive impairment, is a common symptom in people with cirrhosis, a severe liver disease. It can be caused by several factors, including the accumulation of toxins in the body, inflammation, malnutrition, and anemia.  To improve brain fog in cirrhosis, it is important to address the underlying cause. Some steps that may be helpful include:  Managing the progression of cirrhosis: This may involve treating the underlying cause of the liver disease, such as viral hepatitis, alcohol abuse, or nonalcoholic fatty liver disease.  Treating complications of cirrhosis: This may include addressing complications such as ascites (buildup of fluid in the abdomen), encephalopathy (a brain disorder that can cause confusion and loss of consciousness), or bleeding from esophageal varices (enlarged veins in the esophagus).  Managing nutrition: Malnutrition is common in people with cirrhosis, and can contribute to brain fog. It is important to eat a balanced diet and ensure that you are getting enough protein, vitamins, and minerals.  Treating anemia: Anemia, or a deficiency of red blood cells, can cause brain fog and other symptoms. It can be treated with iron supplements, erythropoietin (a medication that stimulates the production of red blood cells), or, in severe cases, blood transfusions.  Avoiding or limiting alcohol: Alcohol can worsen brain fog and other symptoms of cirrhosis. If you have cirrhosis, it is important to limit your alcohol intake or avoid it altogether.  Getting enough sleep: Adequate sleep is important for overall brain health. Try to get at least 7-8 hours of sleep per night.  Staying hydrated: Dehydration can contribute to brain fog. It is important to drink plenty of fluids, especially if you have cirrhosis and are at risk of developing ascites.  Managing stress: Stress can exacerbate brain fog and other symptoms of cirrhosis. It is important to find healthy ways to manage stress, such as through relaxation techniques, exercise, or talking to a mental health professional.  It is important to work with your healthcare team to determine the best approach for managing brain fog in cirrhosis. In some cases, medication may be necessary to manage brain fog and other symptoms. | Similar | 1 | 2 | 2 |
| 69. what can be done to improve insomnia in cirrhosis? | Insomnia can be a common symptom in people with cirrhosis, a condition in which the liver is damaged and cannot function properly. There are several things that can be done to improve insomnia in people with cirrhosis:  Maintain a consistent sleep schedule: Try to go to bed and wake up at the same time every day, even on weekends.  Create a relaxing bedtime routine: Engage in activities that help you relax before bed, such as reading or listening to soothing music.  Avoid stimulating activities before bed: Avoid activities that stimulate the brain, such as watching TV or using electronic devices, in the hours leading up to bedtime.  Avoid caffeine and alcohol: These substances can disrupt sleep and should be avoided close to bedtime.  Avoid napping during the day: While it may be tempting to take a nap during the day to make up for lost sleep, this can actually disrupt your sleep-wake cycle and make it more difficult to fall asleep at night.  Get regular exercise: Regular physical activity can help improve sleep quality, but be sure to avoid vigorous exercise close to bedtime.  Consider medication: If lifestyle changes are not sufficient to improve your sleep, your doctor may recommend medication to help you sleep. It's important to discuss the risks and benefits of any medication with your doctor.  It's also important to work with your doctor to manage the underlying cause of your cirrhosis, as this can help improve your overall health and sleep quality. | Similar | 1 | 2 | 2 |
| 70. What can be done to improve leg swelling in cirrhosis? | Leg swelling, or edema, can be a common symptom in people with cirrhosis, a liver disease that can cause fluid to build up in the body. To improve leg swelling in cirrhosis, some treatment options include:  Diuretics: These medications, also known as water pills, can help your body get rid of excess fluid by increasing urine production.  Sodium restriction: Limiting your intake of salt can help reduce fluid retention.  Compression stockings: Wearing compression stockings can help improve circulation and reduce swelling in your legs.  Elevation: Keeping your legs elevated above your heart can help reduce swelling.  Exercise: Gentle exercises, such as walking or swimming, can help improve circulation and reduce swelling.  Treatment of underlying cause: Treating the underlying cause of your cirrhosis, such as by managing your alcohol intake or taking medications to control hepatitis, can help improve your overall health and reduce swelling.  It's important to talk to your doctor about the best treatment options for your specific situation. In some cases, more aggressive treatment, such as a procedure to remove excess fluid from the body or a liver transplant, may be necessary to improve leg swelling in cirrhosis. | Similar | 1 | 2 | 1 |
| 71. How to manage umbilical hernia from ascites in cirrhosis? | Umbilical hernias can often be managed conservatively with lifestyle modifications, such as avoiding straining during bowel movements and avoiding heavy lifting. It may also be helpful to wear a supportive abdominal binder or garment to help hold the hernia in place and reduce the risk of it becoming larger.  If the hernia is causing symptoms or is at risk of becoming incarcerated (when the hernia becomes stuck and unable to be reduced back into the abdomen), it may need to be repaired surgically. This typically involves making an incision in the abdominal wall and pushing the hernia back into place, then closing the opening with sutures or mesh.  In the case of ascites (accumulation of fluid in the abdomen) due to cirrhosis (liver disease), treatment will depend on the underlying cause of the ascites and the severity of the condition. Options may include diuretics (medications that help reduce fluid accumulation) and paracentesis (a procedure to remove fluid from the abdomen). In some cases, a shunt may be placed to help drain excess fluid from the abdomen. It is important to work closely with a healthcare provider to determine the most appropriate treatment plan. | Similar | 1 | 2 | 2 |
| 72. What are side effects of lactulose? Is there an alternative to lactulose for cirrhosis? | Lactulose is a medication that is used to treat constipation and to reduce the levels of ammonia in the blood. Some common side effects of lactulose include bloating, gas, abdominal cramps, diarrhea, and nausea. These side effects are usually mild and go away on their own after a few days of treatment.  There are a few alternatives to lactulose for the treatment of cirrhosis, a condition in which the liver becomes scarred and is unable to function properly. One option is to use a different type of laxative, such as polyethylene glycol (Miralax) or senna. Another option is to use medications that help to reduce the levels of ammonia in the blood, such as lactulose or rifaximin. It is important to speak with a healthcare provider about the best treatment option for your specific situation. | Lactulose is a medication that is used to treat constipation and to reduce the levels of ammonia in the blood in people with liver disease. Some possible side effects of lactulose include bloating, gas, diarrhea, stomach cramps, and nausea.  There are several alternative medications that may be used to treat constipation and reduce ammonia levels in people with liver disease, including:  Polyethylene glycol (PEG): This is a type of osmotic laxative that works by drawing water into the intestine, which can help to soften the stool and stimulate bowel movements.  Rifaximin: This is an antibiotic that is used to reduce the levels of harmful bacteria in the intestine. It may be used in people with liver disease to help prevent the development of hepatic encephalopathy, which is a condition that can occur when high levels of ammonia in the blood cause brain function to become impaired.  Lactitol: This is a type of sugar alcohol that is used as a laxative. It works by increasing the amount of water in the intestine, which can help to soften the stool and stimulate bowel movements.  Neomycin: This is an antibiotic that is used to reduce the levels of harmful bacteria in the intestine. It may be used in people with liver disease to help prevent the development of hepatic encephalopathy.  It's important to note that the appropriate treatment for constipation and liver disease will depend on the specific underlying cause of the problem and the overall health of the individual. It's always a good idea to speak with a healthcare provider to determine the best course of treatment. | 2,1 | 2,2 | 2,1 |
| 73. What is jaundice? | Jaundice is a condition that occurs when there is an excess of bilirubin, a yellowish substance, in the blood. This can cause the skin and whites of the eyes to appear yellow, and it can also lead to yellowing of the whites of the eyes. Jaundice can be caused by a variety of underlying conditions, such as liver disease, problems with the bile ducts, or certain types of anemia. It can also occur in newborn babies due to an immature liver. Treatment for jaundice depends on the underlying cause, but may include medications to remove excess bilirubin from the body or other supportive care. | Similar | 1 | 2 | 1 |
| 74. How to tell jaundice in a patient with tanned or dark skin? | Jaundice, also known as hyperbilirubinemia, is a condition that occurs when there is an excess of bilirubin in the blood. Bilirubin is a yellowish substance that is produced when the body breaks down red blood cells. In people with tanned or dark skin, it may be more difficult to recognize the signs of jaundice, as the yellowing of the skin and whites of the eyes may not be as noticeable. However, there are other signs and symptoms that may indicate the presence of jaundice in people with tanned or dark skin, including:  Pale or clay-colored stools: Jaundice can cause the stool to become pale or clay-colored, as the bilirubin is not being properly processed and excreted by the liver.  Dark urine: Jaundice can cause the urine to become darker in color, as the bilirubin is being eliminated through the urine instead of the stool.  Itching: Some people with jaundice may experience itching due to the buildup of bilirubin in the skin.  Fatigue: Jaundice can cause fatigue and weakness due to the body's inability to properly process and utilize red blood cells.  If you suspect that you or someone you know may have jaundice, it is important to see a healthcare provider for a proper diagnosis and treatment. The healthcare provider will perform a physical examination and may order laboratory tests to confirm the presence of jaundice and determine its underlying cause. | Similar | 3 | 2 | 3 |
| 75. Can a female patient with cirrhosis get pregnant? | It is generally not recommended for women with cirrhosis to get pregnant because cirrhosis can significantly impair the functioning of the liver and can affect the ability to carry a pregnancy to term. Pregnancy can also worsen the health of the mother and increase the risk of complications such as preterm labor, bleeding, and high blood pressure.  In general, it is important for women with cirrhosis to discuss their plans for pregnancy with their healthcare provider. The provider can evaluate the patient's individual situation and determine the best course of action. In some cases, the provider may recommend that the patient wait until the cirrhosis is better controlled before attempting pregnancy. In other cases, the provider may recommend that the patient not get pregnant at all due to the potential risks to both the mother and the baby.  It is important for women with cirrhosis to carefully consider the potential risks and benefits of pregnancy and to work closely with their healthcare provider to make the best decision for their individual situation. | Similar | 1 | 2 | 1 |
| 76. How to improve leg cramps in a patient with cirrhosis? | Leg cramps can be a common symptom in patients with cirrhosis, a type of liver disease that can lead to a build-up of toxins in the body and fluid accumulation in the legs (known as edema). There are several strategies that may help improve leg cramps in a patient with cirrhosis:  Exercise: Gentle stretching exercises can help improve circulation and reduce cramping.  Hydration: Drinking plenty of fluids can help prevent dehydration, which can contribute to cramping.  Diet: A low-sodium diet may help reduce fluid accumulation in the legs and reduce cramping.  Medications: Some medications, such as diuretics, may be used to help reduce fluid accumulation and improve leg cramps.  Compression stockings: Wearing compression stockings can help improve circulation and reduce cramping.  It is important to consult with a healthcare professional for proper diagnosis and treatment of leg cramps in a patient with cirrhosis. | Similar | 2 | 2 | N/A |
| 77. How to improve albumin levels in cirrhosis? | There are several potential approaches to improving albumin levels in cirrhosis:  Treat the underlying cause of cirrhosis: Cirrhosis is often caused by chronic liver diseases such as hepatitis B and C, alcohol abuse, and nonalcoholic fatty liver disease. Treating the underlying cause of cirrhosis may improve albumin levels.  Administer albumin replacement therapy: Albumin replacement therapy involves administering intravenous (IV) albumin to help increase albumin levels in the body. This approach is typically reserved for people with severe cirrhosis who have low albumin levels and are at risk of complications.  Nutritional support: Ensuring that a person with cirrhosis is getting enough nutrients, including protein, can help improve albumin levels. This may involve supplementing the diet with high-protein foods or supplements, or providing nutritional support through tube feeding or parenteral nutrition (IV nutrition).  Medications: There are a number of medications that may be used to improve albumin levels in people with cirrhosis. These include medications that help to improve liver function, such as pentoxifylline and s-adenosylmethionine (SAMe), and medications that help to reduce inflammation and improve protein synthesis, such as steroids and branched-chain amino acids.  Lifestyle changes: Making lifestyle changes, such as quitting smoking, reducing alcohol consumption, and maintaining a healthy weight, may also help to improve albumin levels in people with cirrhosis.  It's important to note that these approaches should be recommended and monitored by a healthcare provider. It's also important to remember that improving albumin levels may not be possible in all cases, and that treatment for cirrhosis should be tailored to the individual needs of each patient. | Similar | 1 | 2 | 2 |
| **Preventive Medicine** | | | | | |
| 78. Is schedule for other types of cancer screening different to individuals without cirrhosis | Cancer screening recommendations can vary depending on a person's individual risk factors and medical history. It is generally recommended that people at average risk for cancer undergo certain cancer screenings at certain intervals. However, people with certain medical conditions, such as cirrhosis, may have different recommendations for cancer screening.  Cirrhosis is a condition in which scar tissue replaces healthy liver tissue, and it can increase the risk of certain types of cancer, such as liver cancer. People with cirrhosis may be recommended to undergo more frequent or earlier cancer screenings for certain types of cancer. For example, people with cirrhosis may be recommended to undergo liver cancer screenings more frequently than people without cirrhosis.  It is important for individuals to discuss their cancer screening recommendations with their healthcare provider, taking into account their individual risk factors and medical history. The healthcare provider can help determine the appropriate cancer screening schedule for an individual. | Similar | 1 | 2 | 2 |
| 79. Can patients with decompensated cirrhosis get a COVID vaccine? | Yes, people with decompensated cirrhosis can receive the COVID-19 vaccine. Decompensated cirrhosis is a serious and advanced stage of liver disease, and individuals with this condition are at increased risk for severe illness and complications from COVID-19. It is important for people with decompensated cirrhosis to receive the vaccine to help protect them from the virus.  The Centers for Disease Control and Prevention (CDC) recommends that people with decompensated cirrhosis be included in the priority groups for vaccination, along with other people who are at increased risk for severe illness from COVID-19. The specific priority groups may vary depending on the specific vaccine and the availability of vaccine doses.  It is important to consult with a healthcare provider to determine the best course of action and to address any concerns or questions about the vaccine. | Similar | 1 | 1 | N/A |
| 80. Should liver cancer screening be done in patients with previously cured HCV infection? | It is generally recommended that patients with a history of hepatitis C infection undergo liver cancer screening, as they have an increased risk of developing liver cancer. This is because hepatitis C infection can cause chronic liver inflammation, which can lead to scarring of the liver (cirrhosis) and an increased risk of liver cancer.  The American Cancer Society recommends that people with a history of hepatitis C infection who have never had cirrhosis undergo liver cancer screening every 6 to 12 months. For those with a history of hepatitis C infection and cirrhosis, the recommendation is to undergo liver cancer screening every 6 months.  It is important to note that liver cancer screening is not a substitute for regular medical care and other cancer screening tests. It is also important to follow a healthy lifestyle, such as avoiding tobacco and excessive alcohol consumption, to help reduce the risk of developing liver cancer. | Similar | 3 | 2 | 3 |
| 81. How often should patients with cirrhosis with varices undergo endoscopy? | It is generally recommended that patients with cirrhosis and esophageal varices undergo endoscopy at least every 2 to 3 years to monitor the presence and size of the varices. However, the frequency of endoscopy may be increased if the varices are found to be large or if the patient has a history of bleeding from the varices. In addition, patients who have undergone variceal banding or sclerotherapy to treat the varices may need to undergo endoscopy more frequently to monitor the effectiveness of these treatments.  It is important to note that the frequency of endoscopy in patients with cirrhosis and esophageal varices may vary depending on the specific clinical circumstances and the recommendations of the patient's healthcare provider. It is always important for patients with cirrhosis and esophageal varices to follow the treatment recommendations of their healthcare provider to help prevent bleeding and other complications. | The frequency of endoscopy for patients with cirrhosis and varices depends on the size and number of varices and the patient's risk of bleeding. In general, patients with small varices and low risk of bleeding may be monitored with periodic imaging tests, such as ultrasound, to assess the status of their varices. However, patients with larger varices or higher risk of bleeding may require more frequent endoscopy to assess and treat the varices.  The American Association for the Study of Liver Diseases (AASLD) recommends that patients with cirrhosis and large varices (defined as varices with a diameter of at least 5 mm) undergo endoscopy every 1-2 years to assess the varices and perform preventive treatment, such as band ligation or injection sclerotherapy, to reduce the risk of bleeding. In addition, patients with cirrhosis and a history of variceal bleeding may require more frequent endoscopy to monitor and treat the varices.  It is important to note that the frequency of endoscopy may vary depending on the specific circumstances of each patient and the recommendation of their healthcare provider. It is important for patients with cirrhosis and varices to discuss their treatment plan with their healthcare provider and follow their recommendations for monitoring and management of their condition. | 3,3 | 2,2 | 3,3 |
| **Others** | | | | | |
| 82. What is the role of palliative care for patients with cirrhosis? | Palliative care is a type of medical care that focuses on relieving symptoms, improving quality of life, and providing emotional and spiritual support to people with serious illnesses, including cirrhosis. It can be provided at any stage of the disease and can be offered alongside curative treatment.  The goal of palliative care for patients with cirrhosis is to help manage the symptoms and complications of the disease, such as pain, fatigue, nausea, and difficulty breathing, and to provide support for the patient and their family. Palliative care can also help patients with cirrhosis understand their treatment options and make informed decisions about their care.  Palliative care may involve medications to control symptoms, such as pain medication or anti-nausea medication, as well as non-pharmacological approaches, such as relaxation techniques or counseling. Palliative care teams may include doctors, nurses, social workers, chaplains, and other professionals who work together to provide comprehensive care for the patient and their family.  Overall, the role of palliative care in the management of cirrhosis is to help improve the quality of life for patients and their families by addressing the physical, emotional, and spiritual needs of the patient. | Similar | 1 | 1 | N/A |
| 83. What are the symptoms that cirrhosis patients should look out and go to the emergency room for? | Cirrhosis is a serious condition that can lead to life-threatening complications. If you have cirrhosis, it is important to be aware of the signs and symptoms that may indicate a worsening of your condition or the onset of a complication. You should seek immediate medical attention if you experience any of the following symptoms:  Abdominal pain or swelling: This can be a sign of fluid accumulation in the abdomen (ascites) or a complication such as hepatitis or pancreatitis.  Confusion or altered mental status: This can be a sign of hepatic encephalopathy, a condition that occurs when the liver is not able to remove toxins from the blood.  Unusual bleeding or bruising: Cirrhosis can lead to a decrease in the production of clotting factors, which can cause bleeding and bruising.  Yellowing of the skin and whites of the eyes (jaundice): This can be a sign of liver failure or a complication such as hepatitis.  Shortness of breath: This can be a sign of fluid accumulation in the lungs (pulmonary edema) or a complication such as pneumonia.  Chest pain: This can be a sign of a heart attack or a pulmonary embolism.  Fainting or feeling lightheaded: This can be a sign of low blood pressure or a complication such as sepsis.  If you experience any of these symptoms, it is important to seek medical attention as soon as possible. Do not wait to see if the symptoms resolve on their own. | Similar | 1 | 2 | 2 |
| 84. What are the side effects of liver biopsy? How long does it take to recover from a liver biopsy, according to the type of biopsy? | A liver biopsy is a medical procedure in which a small sample of liver tissue is removed and examined under a microscope. It is usually done to diagnose a liver condition or to monitor the effectiveness of treatment.  There are several types of liver biopsy procedures, including percutaneous (through the skin), transvenous (through a vein), laparoscopic (through small incisions in the abdomen), and open (through a larger incision in the abdomen). The specific side effects and recovery time may vary depending on the type of biopsy and the individual patient.  Common side effects of a liver biopsy may include pain or discomfort at the biopsy site, bleeding or bruising, and dizziness or fainting. These side effects are usually temporary and can be managed with medication.  In general, recovery from a liver biopsy takes a few days to a week. You may be advised to rest for a few days and avoid strenuous activities until you feel better. You may also be given specific instructions on caring for the biopsy site and when to resume your normal activities.  It is important to follow your healthcare provider's instructions and report any unusual symptoms or concerns to them. They will be able to provide you with more information on the specific side effects and recovery time for your particular liver biopsy procedure. | Similar | 1 | 2 | 1 |
| 85. Will a cirrhotic liver regenerate? | In some cases, a cirrhotic liver may be able to regenerate, or repair itself to some degree. However, the extent to which this can occur depends on the underlying cause of the liver damage, as well as the stage of cirrhosis. In the early stages of cirrhosis, the liver may be able to repair itself and reverse some of the damage. However, once cirrhosis has progressed to a more advanced stage, the liver may no longer be able to regenerate effectively and the damage may be irreversible. It is important for individuals with cirrhosis to receive appropriate medical care and follow a treatment plan in order to manage the condition and prevent further liver damage. | A cirrhotic liver is one that has been significantly damaged and scarred, and it is not able to regenerate or repair itself in the same way that a healthy liver can. However, the liver is a resilient organ and it can still function to some extent even when it is scarred and damaged. In some cases, treatment can help to slow the progression of liver cirrhosis and may even improve liver function. However, a liver that has been severely damaged by cirrhosis will not be able to regenerate or repair itself completely. | 1,2 | 2,2 | 1 |
| 86. Does the liver heal itself? | Yes, the liver has the ability to regenerate itself. It is able to regenerate damaged or lost tissue and can even regrow from as little as 25% of its original size. The liver is the only organ in the human body that has this ability. This is why it is sometimes possible for people with liver damage to recover and regain full liver function, provided that the cause of the damage is addressed and treated. However, this ability to regenerate can be impaired if the liver is damaged repeatedly or if the damage is too severe. In these cases, liver damage can lead to scarring (fibrosis) and eventually to liver failure. | Yes, the liver is able to regenerate and repair itself. The liver is an important organ that performs many important functions in the body, including detoxification, synthesis of proteins, and production of bile. The liver is able to regenerate because it has a high capacity for cell repair and regrowth. If a part of the liver is damaged or removed, the remaining healthy liver cells can divide and grow to replace the damaged tissue. However, the liver can only regenerate to a certain extent. If the liver is severely damaged or diseased, it may not be able to repair itself completely, and liver transplantation may be necessary. | 1,2 | 2,2 | 1,1 |
| 87. Will the liver function of a patient with liver cirrhosis ever become normal? | The liver function of a patient with cirrhosis may not return to normal, but treatments can help to manage the condition and prevent further damage to the liver. The extent to which the liver function can be improved will depend on the severity of the damage and the underlying cause of the cirrhosis. In some cases, the damage to the liver may be reversible, but in other cases, it may be permanent.  It is important for patients with cirrhosis to follow their treatment plan and make lifestyle changes, such as avoiding alcohol and controlling their weight, to help manage their condition and improve their liver function. In some cases, a liver transplant may be necessary if the liver function is severely impaired. | The liver is a vital organ that performs many important functions in the body, including filtering toxins from the blood, producing bile to help digest fats, and storing nutrients and energy. Cirrhosis is a serious condition that occurs when scar tissue replaces healthy liver tissue, disrupting the organ's ability to function properly. In advanced stages of cirrhosis, the liver may be unable to perform its functions adequately.  In some cases, it may be possible for the liver to repair itself to some extent and restore some of its function. This can happen if the cause of the liver damage is treated or stopped (such as by stopping alcohol consumption in the case of alcoholic liver disease), and if the damage is not too severe. However, in many cases of cirrhosis, the liver damage is irreversible and the liver function will not return to normal.  If the liver function becomes severely impaired, a liver transplant may be necessary in order to sustain life. However, liver transplantation is a major surgery with significant risks and is not suitable for all patients with liver cirrhosis. | 1,1 | 2,2 | 1,1 |
| 88. Does liver cirrhosis increase the risk for falls and fractures? | Yes, liver cirrhosis can increase the risk for falls and fractures. One of the complications of liver cirrhosis is the development of osteoporosis, which is a condition that causes the bones to become weak and brittle. This can increase the risk for falls and fractures, especially in older individuals. Other factors that can contribute to the risk for falls and fractures in people with liver cirrhosis include muscle weakness, balance problems, and changes in blood pressure that can cause dizziness or fainting. It is important for individuals with liver cirrhosis to be aware of these risks and to take steps to prevent falls and fractures, such as getting regular exercise to strengthen the muscles and bones, avoiding activities that may be risky, and using assistive devices if necessary. | Yes, liver cirrhosis can increase the risk of falls and fractures. Cirrhosis is a chronic liver disease that can cause weakness, muscle wasting, and osteoporosis. Osteoporosis is a condition in which the bones become weak and brittle, making them more prone to fractures. People with cirrhosis may also experience dizziness, difficulty with balance, and impaired coordination, which can increase the risk of falls. It is important for people with cirrhosis to be cautious and to take steps to prevent falls, such as using assistive devices, avoiding alcohol, and exercising to improve balance and strength. | 1,1 | 2,2 | 1,1 |
| 89. Can a patient with liver cirrhosis drive a car if they have encephalopathy? | It is generally not safe for a patient with liver cirrhosis and encephalopathy to drive a car. Encephalopathy is a condition that affects brain function and can cause confusion, drowsiness, and other changes in mental function. These symptoms can interfere with a person's ability to safely operate a vehicle. It is important for individuals with encephalopathy to be under the care of a healthcare provider and to follow their recommendations, which may include not driving a car. If you have liver cirrhosis and encephalopathy, it is best to consult with your healthcare provider about your specific situation and whether it is safe for you to drive. | It is not recommended for a patient with liver cirrhosis and hepatic encephalopathy to drive a car due to the potential for impaired judgment, coordination, and reaction time. It is important for the safety of the patient and others on the road that they do not operate a vehicle while experiencing these symptoms. It is also important to note that some states have laws prohibiting individuals with certain medical conditions from driving, and it is the responsibility of the patient to abide by these laws. | 2,1 | 2,2 | 1,1 |
| 90. What can be done for liver cirrhosis patients if they have restless legs at night? | Restless leg syndrome (RLS) is a disorder that causes an uncontroll urge to move your legs, usually because of an uncomfortable sensation. It can be a symptom of liver cirrhosis, and it can also be caused by other factors such as iron deficiency, kidney failure, or nerve damage.  Treatment for RLS in liver cirrhosis patients may include medications to help reduce the symptoms of RLS, such as dopaminergic agents (e.g. pramipexole, ropinirole) or benzodiazepines (e.g. clonazepam). Other treatments that may help alleviate RLS symptoms include lifestyle changes, such as practicing good sleep hygiene and avoiding caffeine and alcohol. It is important for liver cirrhosis patients to discuss treatment options with their doctor, as the underlying cause of their RLS may need to be addressed in order to effectively manage the condition. | Restless leg syndrome (RLS) is a neurological disorder that causes an overwhelming urge to move the legs. It can be a symptom of liver cirrhosis, as well as other underlying medical conditions. If you have liver cirrhosis and are experiencing restless leg syndrome, it is important to speak with your healthcare provider for proper evaluation and treatment.  Treatment for RLS in people with liver cirrhosis may involve medications to address the underlying cause of the RLS, such as iron deficiency or uremia. In some cases, medications that are commonly used to treat RLS, such as dopamine agonists or opioid painkillers, may not be suitable for people with liver cirrhosis due to potential liver toxicity or interactions with other medications that the individual may be taking.  It may also be helpful to implement lifestyle changes that can help to manage RLS symptoms, such as:  Avoiding caffeine and alcohol  Engaging in regular physical activity  Getting enough sleep  Using relaxation techniques, such as meditation or deep breathing, to help you relax  Massaging your legs or applying heat to your legs before bedtime  Again, it is important to speak with your healthcare provider for proper evaluation and treatment of RLS in the context of liver cirrhosis. | 3,1 | 2,2 | 3,1 |
| 91. Can patients with liver cirrhosis take melatonin for sleep? | There is no information available to indicate that melatonin is harmful for people with liver cirrhosis, and it may be safe for use as a sleep aid in this population. However, it is important to note that melatonin is not a substitute for proper medical treatment and that any decisions regarding the use of melatonin or any other supplement should be made in consultation with a healthcare provider. Additionally, it is always a good idea to inform your healthcare provider about all medications and supplements you are taking to ensure that they are safe and appropriate for you. | It is generally not recommended for patients with liver cirrhosis to take melatonin for sleep. Melatonin is metabolized in the liver, and patients with liver cirrhosis may have difficulty metabolizing and eliminating melatonin from the body. This can lead to high levels of melatonin in the body and potentially harmful side effects.  Additionally, patients with liver cirrhosis may be more sensitive to the sedative effects of melatonin, which can further impair liver function. It is important for patients with liver cirrhosis to discuss their sleep concerns with their healthcare provider and to consider other treatment options that may be safer and more appropriate for their condition. | 1,3 | 2,3 | 1,3 |

### Supplemental Table 2. Grading of the responses from the ChatGPT Language Model for HCC-related questions

| Questions | Responses 1 | Responses 2 | Reviewer 1 grades 1. Comprehensive  2. Correct but inadequate  3. Mixed with correct and incorrect/outdated data 4. Completely incorrect | Reviewer 2 grades 1. Comprehensive  2. Correct but inadequate  3. Mixed with correct and incorrect/outdated data 4. Completely incorrect | Reviewer 3 grades for the responses with discrepancies |
| --- | --- | --- | --- | --- | --- |
| **Basic Knowledge** | | | | | |
| 1. What is the liver? what is the liver's function? Where is the liver located? | The liver is a large organ located in the upper right side of the abdomen. It plays a vital role in many important bodily functions, including:  Metabolism: The liver helps to break down nutrients from the diet and store them for energy or later use. It also plays a role in converting excess glucose (sugar) into glycogen, which can be stored in the liver and muscles for use when the body needs additional energy.  Detoxification: The liver helps to filter toxins, such as alcohol and certain medications, from the blood. It also plays a role in the metabolism of drugs, hormones, and other substances that can be harmful to the body if left unchecked.  Blood production: The liver produces certain proteins that are necessary for blood clotting, such as fibrinogen and prothrombin. It also stores and releases iron, which is necessary for the production of red blood cells.  Bile production: The liver produces bile, which is a yellowish-green fluid that helps to digest fats and absorb fat-soluble vitamins. Bile is stored in the gallbladder and released into the small intestine as needed.  Overall, the liver is a vital organ that plays a key role in maintaining the overall health and well-being of the body. | Similar | 2 | 2 | N/A |
| 2. What is hepatocellular carcinoma, also known as liver cancer? | Hepatocellular carcinoma, also known as liver cancer, is a type of cancer that starts in the liver. The liver is an important organ that performs many vital functions in the body, including filtering toxins from the blood, producing bile to help digest fats, and storing nutrients and energy.  Liver cancer is a relatively rare form of cancer, but it is often aggressive and has a poor prognosis. It can be caused by a variety of factors, including chronic infection with the hepatitis B or C virus, heavy alcohol consumption, and certain inherited conditions that increase the risk of liver disease.  Symptoms of liver cancer may include abdominal pain, weight loss, fatigue, and jaundice (a yellowing of the skin and whites of the eyes). The diagnosis of liver cancer is usually made through a combination of imaging tests (such as CT or MRI), blood tests, and biopsy (removing a small sample of tissue for examination under a microscope).  Treatment for liver cancer may include surgery, chemotherapy, radiation therapy, or a combination of these approaches. The choice of treatment depends on the stage and size of the cancer, as well as the overall health of the patient. | Similar | 3 | 2 | 2 |
| 3. Is hepatocellular carcinoma a fast-growing cancer? | Hepatocellular carcinoma, also known as liver cancer, can vary in terms of its growth rate. Some liver cancers can grow rapidly and spread to other parts of the body quickly, while others may grow more slowly. The rate at which a particular liver cancer grows can depend on a number of factors, including the stage of the cancer (how advanced it is), the location of the tumor, and the patient's overall health. It is important to discuss the specific characteristics of an individual's liver cancer with their healthcare team in order to determine the best course of treatment. | Similar | 3 | 2 | 3 |
| 4. Which population does hepatocellular carcinoma affect the most? | Hepatocellular carcinoma (HCC) is a type of liver cancer that is more common in certain populations. It is most commonly found in people who have underlying liver diseases such as chronic hepatitis B or C infection, alcohol-related liver disease, or cirrhosis. It is also more common in people who are obese or have diabetes.  HCC is more common in men than in women and is more common in people over the age of 50. It is also more common in certain geographic regions, such as Africa, Asia, and South America, where hepatitis B infection is more prevalent. HCC can also occur in people who have no known risk factors, although this is less common. | Hepatocellular carcinoma, also known as liver cancer, is more common in certain populations. It is more common in males than females and is more common in people over the age of 60. It is also more common in people with certain underlying health conditions, such as chronic hepatitis B or C infection, cirrhosis, or nonalcoholic fatty liver disease. It is also more common in people who have a family history of liver cancer or who have a history of heavy alcohol consumption. People who live in certain parts of the world, such as Africa and Asia, are also more likely to develop liver cancer. | 1,1 | 3,2 | 3,2 |
| 5. What are the symptoms of hepatocellular carcinoma? | Hepatocellular carcinoma (HCC) is a type of liver cancer that can cause a variety of symptoms, including:  Abdominal pain or discomfort: HCC can cause pain or discomfort in the upper right side of the abdomen, where the liver is located.  Weight loss: HCC can cause unintentional weight loss, as the body may be unable to properly metabolize food and nutrients due to the presence of cancer.  Loss of appetite: HCC can cause a loss of appetite, leading to further weight loss and malnutrition.  Nausea and vomiting: HCC can cause nausea and vomiting, which may be accompanied by abdominal pain or discomfort.  Jaundice: HCC can cause the skin and whites of the eyes to become yellow due to an accumulation of bilirubin in the bloodstream.  Swelling in the abdomen: HCC can cause swelling in the abdomen due to an accumulation of fluid, a condition known as ascites.  Fatigue: HCC can cause fatigue due to the body's inability to properly metabolize nutrients and produce energy.  Fever: HCC can cause a fever due to inflammation or infection.  It's important to note that many of these symptoms can be caused by other conditions, so it's important to see a doctor for proper diagnosis and treatment. If you are experiencing any of these symptoms, it's important to speak with a healthcare professional as soon as possible. | Similar | 3 | 2 | 3 |
| 6. How common is liver cancer? | Liver cancer, also known as hepatocellular carcinoma (HCC), is a relatively uncommon cancer, but it is the fifth most common cause of cancer-related death worldwide. The incidence of liver cancer varies widely by geographical region, with higher rates in developing countries, especially in Africa and East Asia. In the United States, the overall incidence of liver cancer is relatively low, with about 40,000 new cases diagnosed each year. However, the incidence of liver cancer has been increasing in the United States in recent decades, particularly among certain groups such as middle-aged and older men and individuals with chronic liver disease. Risk factors for liver cancer include hepatitis B or C infection, cirrhosis, nonalcoholic fatty liver disease, alcohol abuse, and certain inherited conditions such as hemochromatosis and alpha-1 antitrypsin deficiency. If liver cancer is diagnosed at an early stage, treatment may be more effective, so it is important for individuals to be aware of the signs and symptoms and to seek medical attention if they have any concerns. | Similar | 2 | 2 | N/A |
| 7. What are the causes or risk factors for hepatocellular carcinoma? | Hepatocellular carcinoma (HCC) is a type of liver cancer that occurs when the cells in the liver (hepatocytes) become cancerous and grow out of control. HCC is the most common type of liver cancer and is more common in men than women.  There are several risk factors for HCC, including:  Chronic infection with the hepatitis B or C virus: Chronic infection with these viruses can cause liver damage and increase the risk of HCC.  Cirrhosis: This is a condition in which the liver is scarred and damaged. It can be caused by long-term alcohol abuse, nonalcoholic fatty liver disease (NAFLD), or other liver diseases.  Alcohol abuse: Heavy alcohol consumption can cause liver damage and increase the risk of HCC.  Nonalcoholic fatty liver disease (NAFLD): This is a condition in which excess fat accumulates in the liver, leading to liver damage and an increased risk of HCC.  Diabetes: People with diabetes have an increased risk of developing HCC.  Obesity: Being overweight or obese can increase the risk of HCC.  Exposure to certain chemicals: Exposure to certain chemicals, such as aflatoxins (toxins produced by certain types of mold), can increase the risk of HCC.  Genetic factors: Some people may have a higher risk of developing HCC due to genetic factors.  It's important to note that having one or more of these risk factors does not necessarily mean that a person will develop HCC. However, it's important to be aware of these factors and to take steps to reduce the risk of HCC, such as getting vaccinated against hepatitis B, avoiding alcohol abuse and exposure to toxic chemicals, and maintaining a healthy weight. | Hepatocellular carcinoma (HCC) is a type of liver cancer that originates in the liver cells (hepatocytes). There are several known risk factors for HCC, including:  Chronic hepatitis B and C infections: These viral infections are the most common cause of HCC worldwide.  Cirrhosis: This is a condition in which scar tissue replaces healthy liver tissue, reducing its ability to function. Cirrhosis can be caused by a variety of factors, including alcohol abuse, nonalcoholic fatty liver disease (NAFLD), and autoimmune liver disease.  Alcohol abuse: Heavy alcohol consumption can damage the liver and increase the risk of HCC.  Nonalcoholic fatty liver disease (NAFLD): This is a condition in which excess fat is deposited in the liver, leading to inflammation and scarring. NAFLD is often associated with obesity, insulin resistance, and metabolic syndrome.  Hemochromatosis: This is a genetic disorder that causes the body to absorb and retain too much iron. Excess iron can accumulate in the liver and increase the risk of HCC.  Aflatoxins: These are toxins produced by certain types of fungi that can grow on foods such as grains, peanuts, and corn. Aflatoxins have been linked to an increased risk of HCC in certain parts of the world.  Oral contraceptives: Some studies have suggested that long-term use of oral contraceptives may increase the risk of HCC, particularly in women with other risk factors such as hepatitis B infection or cirrhosis.  Anabolic steroid use: Anabolic steroids are synthetic hormones that can be taken as drugs or injected to increase muscle mass. They have been linked to an increased risk of HCC.  Primary sclerosing cholangitis: This is a rare autoimmune disorder that affects the bile ducts in the liver. It can lead to scarring and inflammation of the liver, increasing the risk of HCC.  It's important to note that having one or more of these risk factors does not necessarily mean that a person will develop HCC. The risk of HCC can be reduced by avoiding behaviors and exposures that increase the risk, such as avoiding heavy alcohol consumption, protecting against viral infections, and maintaining a healthy weight. | 1,1 | 2,1 | 3,3 |
| 8. If a parent had/has liver cancer, are their children at greater risk of getting it? Is liver cancer hereditary? | Liver cancer can be hereditary in some cases, but it is not always passed down from parent to child. There are several factors that can increase a person's risk of developing liver cancer, including certain inherited genetic conditions, chronic viral infections, and certain environmental and lifestyle factors.  Inherited genetic conditions that can increase the risk of liver cancer include hereditary hemochromatosis, which is a condition that causes the body to absorb and retain too much iron, and familial adenomatous polyposis (FAP), which is a condition that causes the development of numerous polyps in the colon and rectum.  Chronic viral infections, such as hepatitis B and hepatitis C, can also increase the risk of liver cancer. These infections are caused by viruses that can damage the liver and lead to liver scarring (cirrhosis) over time. Cirrhosis is a major risk factor for the development of liver cancer.  There are also several environmental and lifestyle factors that can increase the risk of liver cancer, including heavy alcohol consumption, exposure to certain chemicals and toxins, and obesity.  If a parent has liver cancer, their children may be at an increased risk of developing the condition, depending on the underlying cause of the cancer. It is important to discuss any concerns about inherited risk with a healthcare provider. | Similar | 1 | 2 | 2 |
| 9. Is fatty liver disease associated with liver cancer? | Fatty liver disease, also known as nonalcoholic fatty liver disease (NAFLD), is a condition that occurs when there is a build-up of excess fat in the liver. It is a common condition that is often associated with obesity and other conditions such as diabetes and high cholesterol. In most cases, fatty liver disease does not cause serious health problems. However, in some cases, it can progress to a more serious condition called nonalcoholic steatohepatitis (NASH), which can lead to scarring of the liver (cirrhosis) and, in some cases, liver cancer.  Liver cancer is a rare type of cancer that occurs when cells in the liver grow out of control and form a tumor. It is a serious and potentially life-threatening condition that requires prompt medical attention. There are several types of liver cancer, including primary liver cancer, which starts in the liver, and secondary liver cancer, which spreads to the liver from another part of the body.  There is a small increased risk of liver cancer in people with advanced stages of nonalcoholic steatohepatitis (NASH), particularly in those with cirrhosis. However, the overall risk of developing liver cancer in people with NAFLD or NASH is still relatively low. It is important to note that there are many other risk factors for liver cancer, including heavy alcohol consumption, chronic hepatitis B or C infection, and certain inherited conditions.  It is important for people with fatty liver disease to manage their condition through lifestyle changes, such as eating a healthy diet and getting regular exercise, to prevent it from progressing to a more serious stage. It is also important for people with fatty liver disease to have regular check-ups with their healthcare provider to monitor the condition and catch any potential complications early. | Similar | 1 | 2 | 1 |
| 10. Does Hep B cause liver cancer? | Yes, chronic hepatitis B infection is a leading cause of liver cancer. When a person is infected with the hepatitis B virus (HBV), the virus can remain in the body for many years and cause inflammation and damage to the liver. This long-term inflammation can lead to the development of liver scarring (cirrhosis), which increases the risk of developing liver cancer. It is estimated that up to 25% of people with chronic hepatitis B will eventually develop liver cancer.  It's important to note that not everyone with hepatitis B will develop liver cancer. The risk of developing liver cancer is higher for people with chronic hepatitis B who have had the infection for many years, and for those who have other risk factors for liver cancer such as heavy alcohol use, obesity, or other liver diseases. Treatment with antiviral medications can help reduce the risk of liver cancer in people with chronic hepatitis B by controlling the virus and reducing inflammation. If you have hepatitis B, it's important to talk to your healthcare provider about your risk of developing liver cancer and the best ways to manage your health. | Similar | 1 | 2 | 1 |
| 11. Does Hep C cause liver cancer? | Yes, hepatitis C infection can increase the risk of developing liver cancer. Hepatitis C is a viral infection that affects the liver, and it can lead to chronic liver inflammation. Chronic inflammation can damage liver cells and lead to the development of liver scarring (fibrosis) and, in some cases, liver cancer.  The risk of developing liver cancer is higher in people who have had hepatitis C for a long time and in those who have other risk factors for liver cancer, such as heavy alcohol use or cirrhosis (advanced liver scarring). However, it is important to note that not all people with hepatitis C will develop liver cancer.  It is important for people with hepatitis C to receive proper treatment to reduce the risk of liver damage and liver cancer. There are now highly effective treatments available that can cure hepatitis C, which can help reduce the risk of liver cancer and other complications of the infection. If you have hepatitis C, it is important to talk to your healthcare provider about your treatment options. | Similar | 1 | 2 | 1 |
| 12. Does being overweight cause liver cancer? | Being overweight or obese can increase the risk of developing liver cancer. Obesity is a known risk factor for nonalcoholic fatty liver disease (NAFLD), which can progress to nonalcoholic steatohepatitis (NASH) and eventually liver cancer. However, it is important to note that there are other risk factors for liver cancer, including viral hepatitis infection, excessive alcohol consumption, and inherited liver disorders. It is also possible to develop liver cancer without having any of these risk factors. If you are concerned about your risk of liver cancer, it is a good idea to discuss your concerns with a healthcare provider. | Similar | 1 | 2 | 1 |
| 13. Will liver cancer come back after treatment? | It is possible for liver cancer to come back after treatment, although the likelihood of this occurring can vary depending on a number of factors. Some of the things that can influence the likelihood of liver cancer recurring include the stage of the cancer at the time of treatment, the type of treatment that was received, and the overall health of the individual. In general, the earlier liver cancer is diagnosed and treated, the better the chances of long-term survival and a lower risk of the cancer coming back. However, it is important to be aware that there are no guarantees when it comes to cancer, and some people may experience a recurrence despite receiving effective treatment. If you have received treatment for liver cancer and have concerns about the possibility of the cancer coming back, it is important to speak with your medical team for more information. | Liver cancer can come back after treatment, although the likelihood of this happening can vary depending on a number of factors. Some of the factors that can affect the likelihood of liver cancer recurring include the stage of the cancer at the time of treatment, the type of treatment that was received, and the overall health of the patient. In general, the earlier the cancer is caught and treated, the less likely it is to come back. However, it is important to note that there is no guarantee that liver cancer will not recur, and it is important for individuals who have had liver cancer to be monitored for any signs of a recurrence. | 1,2 | 2,2 | 1,2 |
| 14. Can a patient's relative donate their liver to a patient with liver cancer? | Yes, it is possible for a living donor to donate part of their liver to a patient with liver cancer. This is known as living donor liver transplantation. The procedure is complex and carries some risks for the donor, but it can be an option for patients who need a liver transplant but do not have a suitable deceased donor available. The decision to undergo living donor liver transplantation is typically made after careful evaluation by a transplant team, including doctors, surgeons, and social workers. The team will consider factors such as the health and suitability of the donor, the extent of the patient's liver cancer, and the availability of other treatment options. | Similar | 1 | 2 | 1 |
| 15. Does Hepatocellular Carcinoma (HCC) worsen after Hepatitis C (HCV) treatment? | It is generally believed that successful treatment of Hepatitis C (HCV) can prevent the development or progression of Hepatocellular Carcinoma (HCC). However, in some cases, HCC may still develop or worsen despite successful HCV treatment. This can be due to a variety of factors, such as a history of heavy alcohol consumption, exposure to other liver toxins, or the presence of other risk factors for HCC. It is important for individuals with a history of HCV to continue monitoring for the development or worsening of HCC and to seek medical attention if any signs or symptoms arise. | Hepatocellular carcinoma (HCC) is a type of liver cancer that can develop in people with chronic hepatitis C virus (HCV) infection. While HCV treatment can help to reduce the risk of developing HCC, it is not always successful in preventing the development of HCC in people with HCV. In fact, some people may still develop HCC even after successful HCV treatment.  There are several factors that can increase the risk of developing HCC in people with chronic HCV infection. These include:  Advanced age: The risk of developing HCC increases with age, especially in people over the age of 60.  Male gender: Men are more likely to develop HCC than women.  Heavy alcohol use: Chronic alcohol use can increase the risk of developing HCC.  Other liver diseases: People with other liver diseases, such as cirrhosis, are at increased risk of developing HCC.  Obesity: Being overweight or obese can increase the risk of developing HCC.  It is important for people with chronic HCV infection to be regularly monitored for the development of HCC, even if they have received successful HCV treatment. This can involve regular blood tests, ultrasound exams, and other imaging tests to check for signs of HCC. | 1,2 | 2,2 | 1,2 |
| 16. Does COVID accelerate the rate of hepatocellular carcinoma progression? | There is some evidence to suggest that COVID-19 may increase the risk of hepatocellular carcinoma (HCC), which is the most common type of liver cancer. One study found that individuals with COVID-19 and preexisting liver disease, including hepatitis B and C, were at higher risk of developing HCC.  However, it is important to note that the relationship between COVID-19 and the progression of HCC is not fully understood and more research is needed to determine the exact nature of this relationship. It is also worth noting that HCC is typically a slow-growing cancer and it can take years for it to progress to advanced stages.  It is important for individuals with liver disease, including those with HCC, to take steps to protect themselves from COVID-19, such as getting vaccinated and following recommended precautions to prevent the spread of the virus. It is also important for individuals with liver disease to continue to follow their treatment plan and to keep in close contact with their healthcare provider to manage their condition. | There is limited evidence to suggest that COVID-19 may have an effect on the progression of hepatocellular carcinoma (HCC), a type of liver cancer. Some studies have reported that COVID-19 infection may be associated with an increased risk of developing HCC, while others have not found such a connection.  One study published in the journal Hepatology found that COVID-19 infection was significantly associated with the development of HCC in a group of patients with liver cirrhosis. Another study published in the journal Gastroenterology found that COVID-19 infection was associated with an increased risk of HCC in patients with underlying liver disease.  However, it is important to note that these studies are observational and do not establish a causal relationship between COVID-19 and HCC. More research is needed to fully understand the potential relationship between the two.  It is also important to note that HCC is generally a slow-growing cancer, and the rate of progression can vary significantly from one person to another. Factors that may affect the rate of HCC progression include the size and location of the tumor, the presence of underlying liver disease, and the individual's overall health. | 1,1 | 2,1 | 1,1 |
| **Diagnosis** | | | | | |
| 17. How is hepatocellular carcinoma detected/diagnosed? | Hepatocellular carcinoma, also known as liver cancer, is typically detected through a combination of medical history, physical examination, and various imaging and laboratory tests.  Some common methods of detecting and diagnosing hepatocellular carcinoma include:  Blood tests: Blood tests can help detect liver cancer by measuring levels of certain proteins or enzymes that are often elevated in people with liver cancer. These include alpha-fetoprotein (AFP), lactate dehydrogenase (LDH), and gamma-glutamyltransferase (GGT).  Ultrasound: An ultrasound is a non-invasive test that uses sound waves to create images of the inside of the body. It can be used to detect abnormalities in the liver, such as a tumor.  CT scan: A CT scan is a type of X-ray that produces detailed images of the body. It can be used to detect abnormalities in the liver, such as a tumor.  MRI: An MRI is a test that uses magnetic fields and radio waves to create detailed images of the inside of the body. It can be used to detect abnormalities in the liver, such as a tumor.  Biopsy: A biopsy is a procedure in which a small sample of tissue is removed from the liver and examined under a microscope to determine if cancer cells are present. A biopsy is often necessary to confirm a diagnosis of liver cancer.  If liver cancer is suspected, a healthcare provider will typically order one or more of these tests to confirm the diagnosis and determine the stage of the cancer (how advanced it is). The treatment options for liver cancer depend on the stage of the cancer and the overall health of the person. | Similar | 2 | 1 | 3 |
| 18. What are the different stages of hepatocellular carcinoma? | Hepatocellular carcinoma (HCC) is a type of liver cancer that starts in the liver cells (hepatocytes). HCC is typically staged using the TNM staging system, which takes into account the size and spread of the tumor, as well as the presence of cancer in nearby lymph nodes and distant organs. The stages of HCC are as follows:  Stage I: The tumor is small and has not spread beyond the liver.  Stage II: The tumor is larger, but has not spread beyond the liver.  Stage III: The tumor is any size and has spread to nearby lymph nodes or organs.  Stage IV: The tumor is any size and has spread to distant organs or tissues.  It's important to note that the stage of a cancer, including HCC, can change over time as the cancer grows or spreads. Therefore, it's important for individuals with HCC to undergo regular follow-up exams and tests to monitor the cancer's progression. | Hepatocellular carcinoma (HCC) is a type of liver cancer that originates in the cells of the liver. The stages of HCC describe the extent of the cancer and how far it has spread. There are several different staging systems for HCC, but the most commonly used one is the Barcelona Clinic Liver Cancer (BCLC) staging system.  The BCLC staging system divides HCC into five stages:  Early stage (BCLC 0): The cancer is small and confined to the liver. It may or may not have spread to nearby lymph nodes.  Intermediate stage (BCLC A): The cancer is larger and may have spread to nearby lymph nodes or blood vessels, but it is still confined to the liver.  Advanced stage (BCLC B): The cancer has spread to both lobes of the liver or to other parts of the body, such as the lungs or bones.  Very advanced stage (BCLC C): The cancer has spread extensively and is not responding to treatment. The patient may also have significant liver damage or other serious health problems.  Terminal stage (BCLC D): The patient has advanced cancer and serious health problems that make treatment no longer appropriate. Palliative care is the main focus at this stage.  It's important to note that the stage of HCC can change over time as the cancer progresses or responds to treatment. | 3,3 | 3,2 | 3,3 |
| 19. what imaging test is best to diagnose HCC? | Hepatocellular carcinoma (HCC) is a type of liver cancer. There are several imaging tests that can be used to diagnose HCC, including the following:  Ultrasound: This is a non-invasive test that uses high-frequency sound waves to create images of the liver. It is often the first test performed to evaluate liver abnormalities.  Computed tomography (CT) scan: This test uses X-rays and a computer to create detailed images of the liver. It can show the size, shape, and location of a liver tumor.  Magnetic resonance imaging (MRI): This test uses a magnetic field and radio waves to create detailed images of the liver. It can show the size, shape, and location of a liver tumor, as well as the surrounding blood vessels and tissues.  Positron emission tomography (PET) scan: This test uses a small amount of radioactive material, called a tracer, to show how cells in the liver are functioning. It can help distinguish between cancerous and noncancerous tissue.  The best imaging test for diagnosing HCC will depend on the specific circumstances of the patient, including their overall health, the location and size of the suspected tumor, and any other medical conditions they may have. Your doctor will be able to recommend the most appropriate test for you. | Similar | 4 | 2 | 4 |
| 20. Can hepatocellular carcinoma be diagnosed from a PET, MRI or CT scan alone? | Hepatocellular carcinoma (HCC) can be diagnosed using a combination of imaging tests, including PET, MRI, and CT scans. However, none of these tests can definitively diagnose HCC on their own.  PET scans are used to look for areas of increased metabolic activity in the body, which can indicate the presence of cancer cells. However, PET scans are not very sensitive for detecting small tumors or tumors in certain locations, such as the liver.  MRI scans are used to create detailed images of the body using a strong magnetic field and radio waves. MRI scans can be used to detect HCC, but they may not be as sensitive as other imaging tests, such as CT scans.  CT scans are used to create detailed images of the body using x-rays. CT scans are highly sensitive for detecting HCC and can be used to evaluate the size, location, and extent of the tumor.  To diagnose HCC, a healthcare provider will typically order a combination of imaging tests, such as PET, MRI, and CT scans, along with other diagnostic tests, such as blood tests and biopsies. The results of these tests will be used to confirm the diagnosis and determine the best course of treatment. | Yes, hepatocellular carcinoma (HCC) can be diagnosed from a PET, MRI, or CT scan alone. These imaging tests can help identify the presence and location of abnormal growths or tumors in the liver.  PET (Positron Emission Tomography) scans use a small amount of radioactive tracer to create detailed images of the body's organs and tissues. This can help detect cancer cells or other abnormal activity in the liver.  MRI (Magnetic Resonance Imaging) uses a strong magnetic field and radio waves to produce detailed images of the body's internal structures. This can help detect abnormal growths or tumors in the liver.  CT (Computed Tomography) scans use x-rays to create detailed images of the body's internal structures. This can also help detect abnormal growths or tumors in the liver.  It is important to note that while these imaging tests can be helpful in diagnosing HCC, they are not always definitive. Other diagnostic tests, such as liver biopsy or blood tests, may also be needed to confirm the diagnosis. | 3,1 | 3,2 | 4,3 |
| 21. Is it possible to have hepatocellular carcinoma with a normal AFP test? | Yes, it is possible to have hepatocellular carcinoma (HCC) with a normal alpha-fetoprotein (AFP) test. HCC is a type of liver cancer that can develop in people with chronic liver disease, such as cirrhosis or hepatitis. AFP is a protein produced by some types of liver cells and is often elevated in the blood of people with HCC. However, not all people with HCC will have an elevated AFP level, and it is possible to have HCC with a normal AFP level.  There are several factors that can affect the accuracy of an AFP test in detecting HCC. For example, the stage of the cancer, the size of the tumor, and the presence of other liver abnormalities can all affect the level of AFP in the blood. In addition, certain medications and other medical conditions can also cause false-positive or false-negative results on an AFP test.  It is important to note that an AFP test is just one tool that can be used to help diagnose HCC. Other tests, such as imaging studies (such as ultrasound, CT scan, or MRI) and liver biopsy, may also be used to help diagnose HCC and determine the best treatment options. If you are concerned about your risk of HCC or have symptoms that may be related to liver cancer, it is important to speak with a healthcare provider for proper evaluation and management. | Similar | 1 | 2 | 2 |
| 22. what imaging test is best to diagnose HCC? | Hepatocellular carcinoma (HCC) is a type of liver cancer that can be difficult to diagnose because it often does not produce symptoms in the early stages. The most common imaging tests used to diagnose HCC are:  Ultrasound: This test uses high-frequency sound waves to create a detailed image of the liver. It is non-invasive, relatively inexpensive, and widely available.  CT scan: This test uses X-rays to create detailed images of the liver. It is more expensive and requires the use of contrast dye, but it can provide more detailed images than ultrasound.  MRI: This test uses a magnetic field and radio waves to create detailed images of the liver. It is more expensive and requires the use of contrast dye, but it can provide excellent images of the liver and surrounding structures.  PET scan: This test uses a small amount of radioactive tracer to create detailed images of the liver. It is more expensive and requires the use of contrast dye, but it can help determine if cancer is present and if it has spread to other parts of the body.  Ultimately, the best imaging test for diagnosing HCC will depend on the individual patient and their specific circumstances. It is important to discuss the risks and benefits of each test with your healthcare provider to determine the most appropriate course of action. | Similar | 4 | 2 | 4 |
| **Treatment** | | | | | |
| 23. What are the treatment options for hepatocellular carcinoma? | Hepatocellular carcinoma (HCC) is a type of liver cancer that is usually treated with a combination of therapies, including surgery, chemotherapy, targeted therapy, and radiation therapy. The specific treatment options for HCC depend on the stage and location of the cancer, as well as the overall health and preferences of the patient.  Surgery: If the cancer is localized (confined to the liver) and the patient is healthy enough to undergo surgery, the primary treatment option is usually surgical resection or liver transplantation. Surgical resection involves removing the part of the liver that contains the cancerous tumor, while liver transplantation involves replacing the entire diseased liver with a healthy one from a donor.  Chemotherapy: Chemotherapy involves using drugs to kill cancer cells. It can be given before or after surgery to shrink the tumor and kill any remaining cancer cells, or it can be given as a standalone treatment for advanced HCC that is not suitable for surgery.  Targeted therapy: Targeted therapies are drugs that block the growth and spread of cancer cells by targeting specific molecules involved in the growth and development of the cancer. They may be used in combination with chemotherapy or as a standalone treatment for advanced HCC.  Radiation therapy: Radiation therapy uses high-energy beams to kill cancer cells. It may be used to shrink the tumor before surgery, or as a standalone treatment for advanced HCC that is not suitable for surgery.  Other treatment options for HCC may include radiofrequency ablation (a procedure that uses heat to kill cancer cells), cryoablation (a procedure that uses extreme cold to kill cancer cells), and transarterial chemoembolization (a procedure that delivers chemotherapy directly to the liver through the blood vessels).  It is important to note that the success of these treatments depends on the stage and location of the cancer, as well as the overall health of the patient. It is also important to work closely with a healthcare team to determine the best treatment plan. | Similar | 2 | 2 | N/A |
| 24. What is transarterial chemoembolization for the treament of liver cancer? | Transarterial chemoembolization (TACE) is a treatment for liver cancer that involves injecting chemotherapy drugs directly into the liver through the artery that supplies blood to the liver. The chemotherapy drugs are delivered to the liver through a catheter, which is a thin tube inserted through a small incision in the groin.  The procedure is usually performed by an interventional radiologist or a hepatologist, who uses imaging techniques (such as X-ray or ultrasound) to guide the catheter to the correct location in the liver. Once the chemotherapy drugs are injected, the blood supply to the cancerous tissue is blocked off by injecting small particles or a substance called embolic agents, which helps to keep the chemotherapy in the liver and increase its effectiveness.  TACE is often used in combination with other treatments, such as surgery and radiation therapy, to help control the growth and spread of liver cancer. It may be an option for people with liver cancer that has not responded to other treatments or has spread to other parts of the body. However, TACE is not suitable for everyone, and it is important to discuss the potential benefits and risks of the procedure with a medical team before deciding whether it is the right treatment for you. | Similar | 1 | 2 | 1 |
| 25. What are the side effects of transarterial chemoembolization? | Transarterial chemoembolization (TACE) is a procedure used to treat cancer in which chemotherapy drugs are delivered directly to the tumor through the bloodstream. It is commonly used to treat liver cancer and other types of cancer that have spread to the liver.  Like any medical procedure, TACE carries the risk of side effects. Some common side effects of TACE include:  Nausea and vomiting: These side effects may occur after the procedure and can be managed with medications.  Pain: Some people may experience pain at the site where the catheter was inserted. This can usually be managed with over-the-counter pain medications or prescription painkillers.  Fatigue: TACE can cause fatigue, which may last for several days or weeks after the procedure.  Changes in liver function: TACE can affect the function of the liver, and it is important to have regular blood tests to monitor liver function.  Infection: There is a risk of infection at the site where the catheter is inserted. This can usually be treated with antibiotics.  Bleeding: There is a risk of bleeding at the site where the catheter is inserted.  Allergic reactions: Some people may have an allergic reaction to the chemotherapy drugs or the contrast dye used during the procedure.  It's important to discuss the potential risks and side effects of TACE with your doctor before the procedure. | Similar | 1 | 2 | 1 |
| 26. What is transarterial radioembolization for the treament of liver cancer? | Transarterial radioembolization (TARE) is a type of interventional radiology procedure that is used to treat liver cancer. It involves injecting small beads or microspheres containing a radioactive material into the blood vessels that supply blood to the liver tumor. The microspheres become lodged in the blood vessels around the tumor, delivering a high dose of radiation to the cancer cells while minimizing exposure to healthy tissue.  TARE is typically used in combination with other treatments, such as chemotherapy or external beam radiation therapy, to improve the effectiveness of treatment and reduce the size of the tumor. It is usually performed on patients with liver cancer that has spread to other parts of the body (metastatic liver cancer) or who are not candidates for surgery.  The procedure is performed under conscious sedation and usually takes about an hour. The patient is awake during the procedure but is given a medication to help them relax. The interventional radiologist uses imaging guidance, such as computed tomography (CT) or ultrasound, to guide the placement of the microspheres into the blood vessels around the liver tumor.  TARE is generally well tolerated and has a low risk of complications. However, as with any medical procedure, there are potential risks and side effects that should be discussed with a healthcare provider. | Similar | 3 | 1 | 3 |
| 27. What are the side effects of transarterial radioembolization? | Transarterial radioembolization (TARE) is a procedure used to treat cancer that involves injecting tiny beads or spheres containing a radioactive substance into the blood vessels that supply a tumor. As with any medical procedure, there are potential side effects associated with TARE. These may include:  Nausea and vomiting: Some people may experience nausea and vomiting after the procedure, which can usually be managed with medications.  Fatigue: It is common for people to feel tired or fatigued after TARE. This may be due to the effects of the procedure or the underlying cancer.  Pain: Some people may experience pain at the site where the beads or spheres were injected. This can usually be managed with pain medications.  Diarrhea: TARE may cause diarrhea in some people, which can usually be managed with medications.  Rash or itching: Some people may develop a rash or experience itching after TARE, which can usually be managed with medications.  Changes in liver function: TARE may cause changes in liver function, which can be monitored with blood tests.  It is important to discuss the potential side effects of TARE with your healthcare team before the procedure. They can help you understand what to expect and how to manage any side effects that may occur. | Similar | 1 | 2 | 1 |
| 28. What is radiofrequency ablation for the treatment of liver cancer? | Radiofrequency ablation (RFA) is a minimally invasive procedure used to treat liver cancer and other types of tumors. It involves the use of high-frequency radio waves to heat and destroy cancer cells in the liver.  During the procedure, a thin needle or probe is inserted through the skin and into the tumor. The probe is then used to deliver radiofrequency energy, which generates heat and destroys the cancer cells. RFA is typically performed under local anesthesia, and the procedure takes about 1-2 hours to complete.  RFA is often used as an alternative to surgery for the treatment of small, early-stage liver cancers that are not suitable for surgery. It may also be used to palliate (relieve symptoms) in patients with advanced liver cancer who are not candidates for other treatments.  RFA has a high success rate and is generally well-tolerated, with few serious side effects. However, as with any medical procedure, there are risks and potential complications associated with RFA, including bleeding, infection, and damage to surrounding tissues. Your doctor will discuss the potential risks and benefits of RFA with you before the procedure. | Similar | 3 | 2 | 2 |
| 29. Whats is stereotactic body radiation therapy treatment for liver cancer? | Stereotactic body radiation therapy (SBRT) is a type of radiation therapy that uses high doses of radiation to target a specific area of the body, such as the liver. It is often used to treat small, localized tumors that cannot be treated with surgery or traditional radiation therapy.  During SBRT, the patient is placed in a specialized frame that helps to accurately deliver the radiation to the targeted area. The frame is attached to the body and helps to ensure that the patient stays still during treatment. The radiation is delivered using a specialized machine that is able to deliver high doses of radiation to the targeted area with a high degree of accuracy.  SBRT is typically administered in a series of treatments, with each treatment lasting only a few minutes. The total number of treatments required depends on the size and location of the tumor, as well as the patient's overall health.  SBRT is generally well tolerated, with side effects being typically mild and temporary. Some common side effects of SBRT include fatigue, skin irritation, and nausea. More serious side effects, such as liver damage or problems with the digestive system, are rare.  Overall, SBRT is a promising treatment option for people with liver cancer, particularly those who are not candidates for surgery or traditional radiation therapy. It can be an effective way to shrink or eliminate the cancerous tumors, and may help to improve the patient's quality of life. | Similar | 1 | 2 | 2 |
| 30. What are the side effects of stereotactic body radiation therapy? | Stereotactic body radiation therapy (SBRT) is a highly targeted form of radiation therapy that delivers a high dose of radiation to a specific area of the body. SBRT is typically well-tolerated, and side effects are generally mild and temporary. However, as with any medical treatment, there are potential side effects associated with SBRT.  Common side effects of SBRT may include:  Fatigue: Many people experience fatigue during and after SBRT treatment. This can be due to the physical and emotional demands of treatment and the body's natural response to radiation exposure.  Skin changes: The skin in the treated area may become red, dry, or tender. It may also become darker or lighter in color. These changes are usually temporary and will resolve after treatment is complete.  Nausea and vomiting: Some people may experience nausea and vomiting during or after SBRT treatment. This can usually be managed with medications prescribed by the treating physician.  Diarrhea: Diarrhea is a common side effect of SBRT, especially in people with rectal or anal cancer. It can usually be managed with medications prescribed by the treating physician.  Pain: Some people may experience pain in the treated area during or after SBRT treatment. This can usually be managed with medications prescribed by the treating physician.  Difficulty swallowing: SBRT to the head and neck region may cause difficulty swallowing, which can usually be managed with medications and supportive care.  Urinary symptoms: SBRT to the prostate gland may cause urinary symptoms such as frequency, urgency, and difficulty urinating. These symptoms can usually be managed with medications and supportive care.  It's important to talk to your doctor about the potential side effects of SBRT and how they may be managed. They will be able to provide more information on the specific side effects that may be associated with your treatment. | Similar | 2 | 1 | 2 |
| 31. What is the role of liver transplantation for the treatment of liver cancer? | Liver transplantation can be an effective treatment option for certain types of liver cancer, particularly if the cancer is limited to a small area of the liver and has not spread to other organs.  In a liver transplantation procedure, the surgeon removes the diseased liver and replaces it with a healthy liver from a donor. The donor liver can be from a deceased donor or from a living donor.  There are several criteria that must be met in order for a person to be eligible for a liver transplant due to liver cancer. These include:  The cancer must be limited to a small area of the liver and has not spread to other organs  The cancer must not have recurred after previous treatment  The person must be in good general health and have no other serious medical conditions that would make a transplant risky  The person must be willing and able to follow a strict medication regimen after the transplant  If a person is eligible for a liver transplant due to liver cancer, the procedure can be highly successful in providing long-term survival and an improved quality of life. However, it is important to note that liver transplantation is a major surgery with risks and complications, and it may not be suitable for everyone. It is important to discuss the potential risks and benefits with a medical team experienced in liver transplantation. | Similar | 3 | 2 | 3 |
| 32. What are the side effects of liver transplantation | Liver transplantation is a major surgery that carries a number of risks and potential complications. Some of the most common side effects of liver transplantation include:  Infection: As with any surgery, there is a risk of infection following a liver transplant. This can occur at the surgical site or elsewhere in the body.  Rejection: The immune system may recognize the new liver as a foreign object and attempt to reject it. This can occur even if the donor and recipient are a good match.  Bleeding: There is a risk of bleeding during and after the surgery.  Blood clots: Blood clots can form in the veins or arteries following surgery, which can lead to serious complications such as pulmonary embolism (a blockage in the arteries of the lungs).  Failure of the transplanted liver: The transplanted liver may not function properly, or it may fail completely.  Scarring (fibrosis) of the transplanted liver: Scarring of the transplanted liver may occur over time, which can lead to problems with liver function.  Side effects of immunosuppressant medications: In order to prevent rejection of the transplanted liver, patients will need to take immunosuppressant medications. These medications can have a number of side effects, including an increased risk of infection, high blood pressure, kidney damage, and an increased risk of certain types of cancer.  It's important to discuss these risks and potential complications with your healthcare team before undergoing liver transplantation. | Similar | 2 | 2 | N/A |
| 33. What is the role of liver resection for the treatment of liver cancer? | Liver resection is a surgical procedure in which a portion of the liver is removed. It is often used as a treatment for liver cancer, also known as hepatocellular carcinoma (HCC).  The liver is a vital organ that performs a number of important functions in the body, including filtering toxins from the blood, producing bile to aid in digestion, and storing nutrients and energy. As a result, liver resection can be a major surgery with potential risks and complications. However, it can also be an effective way to remove cancerous tumors from the liver and improve the prognosis for patients with liver cancer.  Liver resection is typically only considered if the cancer is confined to a specific area of the liver and has not spread to other parts of the body. The extent of the resection will depend on the size and location of the tumor, as well as the overall health of the patient. In some cases, a partial liver resection may be sufficient to remove the cancerous tissue, while in other cases, a more extensive resection may be necessary.  After a liver resection, the remaining liver tissue will typically regenerate and return to its normal function. However, it is important for patients to follow a healthy lifestyle, including eating a healthy diet and avoiding harmful substances such as alcohol, to help maintain the health of the liver and reduce the risk of cancer recurrence. | Similar | 1 | 2 | 3 |
| 34. What are the side effects of liver resection | Liver resection is a surgical procedure in which a part of the liver is removed. Like any surgical procedure, it carries certain risks and potential complications.  Some common side effects of liver resection include:  Pain at the incision site  Blood loss and the need for a blood transfusion  Infection  Changes in liver function  Damage to nearby organs or structures  Wound healing problems  Adverse reactions to anesthesia  Less common but potentially serious complications of liver resection include:  Liver failure  Bile leak  Collection of fluid in the abdomen (ascites)  Development of a blood clot  Development of a hernia  It's important to discuss the risks and potential complications of liver resection with your surgeon before the procedure. Your surgeon will also discuss the precautions that will be taken to minimize the risks and what you can expect during your recovery. | Similar | 1 | 2 | 1 |
| 35. What is immunotherapy for liver cancer? | Immunotherapy is a type of treatment that uses the body's own immune system to help fight cancer. It can be used to treat liver cancer, also known as hepatocellular carcinoma (HCC).  There are several types of immunotherapy that may be used to treat liver cancer, including:  Checkpoint inhibitors: These medications help to boost the immune system's ability to recognize and attack cancer cells. Examples include PD-1 and CTLA-4 inhibitors.  Oncolytic viruses: These are viruses that can infect and kill cancer cells. They can be injected directly into the tumor or delivered through the bloodstream.  Vaccines: These are designed to stimulate the immune system to recognize and attack cancer cells.  Adoptive cell transfer: This involves taking immune cells from the patient, activating them in the laboratory to recognize and attack cancer cells, and then infusing them back into the patient.  Immunotherapy can be used alone or in combination with other treatments, such as chemotherapy or radiation therapy. It may be an option for people with liver cancer who have not responded to other treatments, or for those who are not candidates for surgery.  It's important to note that immunotherapy is still being researched and is not yet widely available for the treatment of liver cancer. It is not yet clear how effective it is at treating this type of cancer. | Similar | 1 | 3 | 3 |
| 36. What type of chemotherapy is used for HCC? | HCC stands for hepatocellular carcinoma, which is a type of cancer that affects the liver. There are several different types of chemotherapy that may be used to treat HCC, depending on the specific characteristics of the cancer and the overall health of the patient. Some of the chemotherapy drugs that may be used to treat HCC include:  Doxorubicin: This is a chemotherapy drug that is commonly used to treat a variety of different types of cancer, including HCC. It works by damaging the DNA of cancer cells, which can help to slow or stop their growth and reproduction.  Gemcitabine: This is a chemotherapy drug that is often used to treat pancreatic cancer, but it may also be used to treat HCC. It works by inhibiting the production of DNA and RNA, which are important for cancer cell growth and reproduction.  Fluorouracil: This is a chemotherapy drug that is commonly used to treat a variety of different types of cancer, including HCC. It works by inhibiting the production of DNA and RNA, which can help to slow or stop the growth and reproduction of cancer cells.  Oxaliplatin: This is a chemotherapy drug that is often used to treat colorectal cancer, but it may also be used to treat HCC. It works by damaging the DNA of cancer cells, which can help to slow or stop their growth and reproduction.  It's important to note that chemotherapy is just one of several different treatment options that may be used to treat HCC. Other treatment options may include surgery, radiation therapy, and targeted therapies, which are medications that specifically target certain proteins or pathways involved in cancer cell growth and survival. Your healthcare team will work with you to develop a treatment plan that is tailored to your specific needs and circumstances. | Similar | 4 | 3 | 4 |
| 37. What are the side effects of lenvima? | Lenvima (lenvatinib) is a prescription medication used to treat certain types of cancer. Like all medications, Lenvima can cause side effects. The most common side effects of Lenvima include:  Fatigue  High blood pressure  Diarrhea  Decreased appetite  Nausea and vomiting  Constipation  Stomach pain or discomfort  Weight loss  Rash or dry skin  Swelling in the arms, legs, or ankles  Less common but more serious side effects of Lenvima include:  Heart problems, such as abnormal heart rhythm or heart failure  Liver problems, including liver failure  Changes in blood cell counts, which can increase the risk of infection or bleeding  Hypothyroidism (low thyroid hormone levels)  Allergic reactions  If you experience any of the above side effects while taking Lenvima, it's important to tell your doctor as soon as possible. They can suggest ways to manage the side effects and may adjust your treatment plan if necessary. | Similar | 1 | 2 | 1 |
| 38. What are the side effects of atezolizumab? | Atezolizumab is a monoclonal antibody that is used to treat certain types of cancer, including bladder cancer, non-small cell lung cancer, and breast cancer. Like all medications, atezolizumab can cause side effects. The most common side effects of atezolizumab include:  Fatigue: A feeling of tiredness or lack of energy.  Nausea: A feeling of discomfort or dizziness that may lead to vomiting.  Diarrhea: Loose or watery stools.  Rash: A red, raised, itchy area on the skin.  Itching: A sensation that causes the desire to scratch the skin.  Decreased appetite: A decreased desire to eat.  Constipation: Infrequent or hard stools.  Abdominal pain: Pain or discomfort in the abdominal (stomach) area.  Cough: A reflex that helps to clear foreign substances from the airways.  Shortness of breath: Difficulty breathing or a feeling of not being able to get enough air.  Less common side effects of atezolizumab may include fever, headache, difficulty sleeping, and muscle or joint pain. If you experience any of these side effects while taking atezolizumab, you should contact your healthcare provider. It is important to let your healthcare provider know about any side effects you experience so they can determine the appropriate course of action. | Atezolizumab is a medication that is used to treat certain types of cancer. It belongs to a class of drugs called monoclonal antibodies, which work by targeting specific proteins in the body to help the immune system fight cancer cells.  Like all medications, atezolizumab can cause side effects. The most common side effects of atezolizumab include:  Fatigue  Nausea  Fever  Diarrhea  Decreased appetite  Rash  Less common, but more serious side effects of atezolizumab may include:  Infusion reactions (reactions that occur during the infusion of the medication)  Serious infections  Lung problems  Colitis (inflammation of the colon)  Hepatitis (inflammation of the liver)  It's important to note that not everyone who takes atezolizumab will experience side effects. If you are taking atezolizumab and experience any side effects, it is important to talk to your healthcare provider. They can help you manage any side effects you may be experiencing. | 1,2 | 2,2 | 2,2 |
| 39. What are the side effects of bevacizumab? | Bevacizumab is a medication that is used to treat certain types of cancer. It works by blocking the action of a protein called vascular endothelial growth factor (VEGF), which helps to promote the growth of new blood vessels in the body. This can help to slow or stop the growth of cancer cells, which need a good blood supply in order to grow and spread.  Like all medications, bevacizumab can cause side effects. The most common side effects of bevacizumab include:  High blood pressure  Tiredness or fatigue  Headache  Nausea and vomiting  Diarrhea  Loss of appetite  Constipation  Mouth sores  Rash  Fever  Infection  Less common, but more serious side effects of bevacizumab can include:  Bleeding or bruising more easily than normal  A hole in the wall of the intestine (intestinal perforation)  A blockage in the intestine (intestinal obstruction)  Wound healing problems  Blood clots in the veins or arteries  A hole in the wall of the stomach or esophagus (gastrointestinal perforation)  Heart attack or stroke  It's important to tell your healthcare provider if you experience any of these side effects while taking bevacizumab. Your healthcare provider can help you manage the side effects and may be able to adjust your treatment plan to help reduce the severity of the side effects. | Similar | 1 | 2 | 2 |
| 40. What are the side effects of sorafenib? | Sorafenib is a medication used to treat certain types of cancer. Some common side effects of sorafenib include:  Skin rash or dryness  Diarrhea  Fatigue  Nausea and vomiting  Loss of appetite  High blood pressure  Mouth sores  Hand-foot syndrome (a condition in which the skin on the palms of the hands and soles of the feet becomes painful and tender)  Less common but more serious side effects of sorafenib can include:  Bleeding or bruising easily  Fever or infections  Shortness of breath  Chest pain  Swelling in the legs or feet  Decreased urine output  It's important to discuss the potential side effects of any medication with your healthcare provider before starting treatment. They can help you understand the risks and benefits of the medication and manage any side effects that may occur. | Sorafenib is a medication that is used to treat certain types of cancer. It is a type of targeted therapy, which means it is designed to specifically target cancer cells and minimize the effect on normal cells.  Like all medications, sorafenib can cause side effects. The most common side effects of sorafenib include:  Skin rash or dry skin  Diarrhea  Fatigue  Loss of appetite  Nausea and vomiting  High blood pressure  Hand-foot syndrome (redness, swelling, and pain in the palms of the hands and soles of the feet)  Less common side effects of sorafenib include:  Hair loss  Anemia (low red blood cell count)  Decreased white blood cell count  Decreased platelet count  Swelling of the legs and ankles  Changes in liver function test results  It is important to discuss the potential side effects of sorafenib with your doctor before starting treatment. Your doctor will monitor you closely for any side effects and will work with you to manage them. | 1,2 | 2,2 | 1,1 |
| 41. What are the side effects of regorafenib? | Regorafenib is a prescription medication that is used to treat certain types of cancer. It is a chemotherapy drug that belongs to a class of medications called kinase inhibitors. Like all medications, regorafenib can cause side effects.  The most common side effects of regorafenib include:  Fatigue  Hand-foot skin reaction (redness, swelling, and pain on the palms of the hands and soles of the feet)  Diarrhea  Decreased appetite  Nausea  Vomiting  Mouth sores  High blood pressure  Liver problems (elevated liver enzymes)  Rash  Weight loss  Interstitial lung disease (inflammation of the lungs)  Less common side effects of regorafenib include:  Hemorrhage (bleeding)  Infection  Thrombocytopenia (low platelet count)  Anemia (low red blood cell count)  Hypothyroidism (low thyroid hormone levels)  Peripheral neuropathy (nerve damage in the arms and legs)  Pulmonary embolism (blood clot in the lungs)  These are not all of the possible side effects of regorafenib. If you experience any side effects while taking regorafenib, it is important to contact your healthcare provider. They can advise you on how to manage the side effects and determine if any further evaluation or treatment is needed. | Similar | 1 | 2 | 1 |
| 42. What are the side effect of cabozantinib? | Cabozantinib is a prescription medication used to treat certain types of cancer. As with all medications, it can cause side effects. The most common side effects of cabozantinib include diarrhea, nausea, fatigue, decreased appetite, weight loss, and mouth sores. Other possible side effects include:  High blood pressure  Swelling in the hands, feet, or ankles  Changes in skin color or texture  Rash or itching  Dry skin  Changes in liver function tests  Changes in blood counts  It's important to let your healthcare provider know if you experience any side effects while taking cabozantinib. In some cases, side effects can be managed or relieved with medications or other treatments. It's also important to follow your healthcare provider's instructions for managing any side effects. | Cabozantinib is a medication that is used to treat certain types of cancer. It works by inhibiting the activity of certain proteins that are involved in the growth and spread of cancer cells.  Like all medications, cabozantinib can cause side effects. Some common side effects of cabozantinib include:  Diarrhea: This is a common side effect of cabozantinib and can often be managed with medications prescribed by your healthcare provider.  Fatigue: Many people who take cabozantinib experience fatigue, which can be managed with rest and by pacing yourself during activities.  Mouth sores: Cabozantinib can cause mouth sores or ulcers, which can be managed with proper oral hygiene and by avoiding spicy or acidic foods.  Nausea: Nausea is a common side effect of cabozantinib and can often be managed with medications prescribed by your healthcare provider.  Skin changes: Cabozantinib can cause changes in the skin, including dryness, rash, or redness. These changes can often be managed with moisturizers or other skin care products.  Other possible side effects of cabozantinib include high blood pressure, anemia, and decreased appetite. If you experience any of these side effects, it is important to contact your healthcare provider for further evaluation and management.  It is important to remember that everyone is different and may experience different side effects. Be sure to discuss any concerns you have about the side effects of cabozantinib with your healthcare provider. | 2,2 | 2,2 | N/A |
| 43. What side effects are there for the drug combination of Atezolizumab & Bevacizumab? | Atezolizumab and bevacizumab are both medications that are used to treat cancer. They are often used in combination with other cancer treatments, such as chemotherapy.  Common side effects of atezolizumab and bevacizumab may include:  Fatigue  Nausea  Diarrhea  Constipation  Decreased appetite  Rash or itching  Headache  Fever  Less common, but more serious side effects may include:  Infusion-related reactions, including allergic reactions, which may occur during or shortly after receiving the medication  Decreased white blood cell counts, which can increase the risk of infection  Blood clotting disorders  Heart problems, such as heart attack or heart failure  High blood pressure  Kidney problems  Liver problems  Perforation or fistula (abnormal connection) in the intestine  It's important to note that everyone reacts differently to medications, and not everyone will experience these side effects. It's also important to talk to your doctor about the potential risks and benefits of treatment with atezolizumab and bevacizumab. Your doctor can help you understand what to expect during treatment and how to manage any side effects that may occur. | Similar | 2 | 2 | N/A |
| 44. What side effects are there for the drug combination of Durvalumab and Tremelimumab? | Durvalumab and tremelimumab are monoclonal antibodies that are used in combination to treat certain types of cancer. The combination of these drugs is known as the Durvalumab and Tremelimumab regimen.  Common side effects of the Durvalumab and Tremelimumab regimen may include:  Fatigue  Nausea  Diarrhea  Rash or skin irritation  Decreased appetite  Fever  Less common side effects may include:  Inflammation of the lungs (pneumonitis)  Inflammation of the colon (colitis)  Changes in liver function test results  Decreased white blood cell count (neutropenia)  Decreased platelet count (thrombocytopenia)  Inflammation of the thyroid gland (thyroiditis)  Inflammation of the pancreas (pancreatitis)  Inflammation of the eye (uveitis)  It's important to note that these are just some of the possible side effects of the Durvalumab and Tremelimumab regimen, and that not all individuals who receive this treatment will experience these side effects. It's also important to discuss the potential risks and benefits of treatment with your healthcare provider. | Similar | 1 | 2 | 1 |
| 45. What is the life expectancy of a person with hepatocellular carcinoma based on the cancer stage? (Prognosis) | Hepatocellular carcinoma (HCC) is a type of liver cancer that is usually associated with underlying liver damage, such as cirrhosis. The prognosis for HCC depends on several factors, including the stage of the cancer, the underlying liver function, and the presence of other medical conditions.  The stage of HCC refers to the size and extent of the cancer. In general, the earlier the stage of HCC, the better the prognosis. However, HCC is often not diagnosed until it is at an advanced stage, which can affect the prognosis.  The five-year survival rate for HCC is generally low, especially for those with advanced cancer. According to the American Cancer Society, the five-year survival rate for HCC is about 15% for people with stage I cancer, 7% for those with stage II cancer, 3% for those with stage III cancer, and less than 1% for those with stage IV cancer.  However, it's important to note that these are just general estimates and that individual prognoses can vary widely. It's always best to discuss the specifics of your prognosis with your healthcare team, who can take into account your individual circumstances and treatment options. | Similar | 4 | 2 | 4 |
| 46. Can hepatocellular carcinoma be cured? | Hepatocellular carcinoma (HCC), also known as liver cancer, can be challenging to treat, and the outlook for people with HCC depends on various factors, including the stage of the cancer, the underlying cause of the cancer, and the overall health of the person. In general, the earlier HCC is diagnosed and treated, the better the chances of a cure.  There are several treatment options for HCC, including surgery, chemotherapy, targeted therapy, and radiation therapy. The most effective treatment option will depend on the specifics of the case, such as the size and location of the tumor, the general health of the person, and other medical conditions they may have.  For people with early-stage HCC, surgery to remove the cancerous tissue may be an option. This can be curative if all of the cancerous tissue can be completely removed. However, not all people with HCC are candidates for surgery due to the location of the tumor or other health issues that make the procedure too risky.  For people with more advanced HCC, other treatments such as chemotherapy, targeted therapy, or radiation therapy may be used to try to shrink the tumor or slow its growth. These treatments may be used alone or in combination with each other. While these treatments can help to control the cancer, they are generally not curative for advanced HCC.  It is important to speak with a healthcare provider about the specific treatment options and prognosis for HCC. They can provide more information about the treatment options available and the likelihood of a cure based on the specific circumstances of the case. | Similar | 2 | 3 | 2 |
| 47. Are Stereotactic Body Radiation Therapy (SBRT) and Immunotherapy combined together in hepatocellular carcinoma treatment? | It is possible to combine stereotactic body radiation therapy (SBRT) with immunotherapy as a treatment option for hepatocellular carcinoma (HCC), a type of liver cancer. SBRT is a type of external beam radiation therapy that delivers high doses of radiation to a precise location in the body with the goal of minimizing exposure to surrounding healthy tissue. Immunotherapy, on the other hand, is a type of treatment that uses the body's own immune system to help fight cancer.  There is evidence to suggest that combining SBRT with immunotherapy may be more effective in treating HCC than either treatment alone. For example, one study found that the combination of SBRT and immune checkpoint inhibitor therapy (a type of immunotherapy) was associated with improved survival outcomes in patients with HCC compared to SBRT alone. However, more research is needed to fully understand the benefits and potential risks of combining these treatments for HCC.  It's important to note that the optimal treatment for HCC will depend on the individual patient's specific circumstances, including the stage and location of the cancer, as well as the overall health and preferences of the patient. It is important for patients to discuss all treatment options with their healthcare team to determine the most appropriate treatment plan for their specific situation. | Similar | 1 | 2 | 1 |
| 48. Why do doctors test for Tyrroid-Stimulating Hormone (TSH) levels for immunotherapy treatment when patients have HCC? | Thyroid-stimulating hormone (TSH) is produced by the pituitary gland in the brain and plays a crucial role in the regulation of thyroid function. The thyroid gland produces hormones that are important for many functions in the body, including metabolism and energy production. TSH levels can be used as an indicator of thyroid function, and abnormal TSH levels can be a sign of a thyroid disorder.  In patients with hepatocellular carcinoma (HCC), a type of liver cancer, doctors may test for TSH levels as part of their evaluation of the patient's overall health and to determine whether the patient is a good candidate for immunotherapy treatment. Immunotherapy is a type of cancer treatment that uses the body's own immune system to fight cancer cells. It is a complex and highly personalized treatment that requires careful evaluation of the patient's health and cancer status.  There are several reasons why doctors may test for TSH levels in patients with HCC who are considering immunotherapy treatment. One reason is that the immune system plays a crucial role in the body's response to cancer, and abnormal thyroid function can affect the immune system. TSH levels can provide important information about the patient's thyroid function and help the doctor determine the best course of treatment. Additionally, some immunotherapy treatments may affect thyroid function, and monitoring TSH levels can help the doctor manage any potential side effects.  It is important to note that TSH levels are just one factor that doctors consider when evaluating a patient for immunotherapy treatment. They will also consider other factors, such as the stage and grade of the cancer, the patient's overall health, and the potential side effects of treatment. | Similar | 1 | 2 | 2 |
| 49. Why does bilirubin go up after Yttrium-90 (Y90) treatment? | Yttrium-90 (Y90) is a radioactive isotope that is used in the treatment of certain types of cancer, such as liver cancer. When Y90 is used in the treatment of liver cancer, it is typically administered through a process called radioembolization. In this procedure, Y90 is delivered directly to the liver via a catheter that is inserted through the artery.  One potential side effect of Y90 treatment is an increase in bilirubin levels in the blood. Bilirubin is a pigment that is produced by the breakdown of red blood cells. It is normally removed from the body by the liver, but if the liver is damaged or not functioning properly, bilirubin can accumulate in the blood, leading to an increase in bilirubin levels.  There are several possible reasons why Y90 treatment may cause an increase in bilirubin levels. One possibility is that the Y90 particles may damage the liver cells, leading to inflammation and impaired function. This could cause the liver to become less efficient at removing bilirubin from the blood, leading to an accumulation of bilirubin in the bloodstream.  Another possible reason is that Y90 treatment may cause blockages in the bile ducts, which are the tubes that carry bile from the liver to the intestine. If the bile ducts become blocked, bile cannot flow freely from the liver to the intestine, and bilirubin can accumulate in the blood.  It is important to note that an increase in bilirubin levels after Y90 treatment is not always a cause for concern. In many cases, the increase in bilirubin levels is temporary and will resolve on its own over time. However, if the increase in bilirubin levels is severe or persistent, it may be necessary to seek medical attention. Your healthcare provider will be able to assess your bilirubin levels and determine the appropriate course of treatment. | Similar | 1 | 1 | N/A |
| 50. Does bilirubin level matter in hepatocellular carcinoma treatment? | Bilirubin is a yellowish substance that is produced when red blood cells break down. It is normally eliminated from the body through the liver and excreted in the bile. Elevated levels of bilirubin in the blood, a condition known as hyperbilirubinemia, can be a sign of liver damage or dysfunction.  In the context of treatment for hepatocellular carcinoma (HCC), or liver cancer, bilirubin levels can be an important factor to consider. HCC is a type of cancer that arises from the liver cells, and it can be associated with underlying liver damage or disease. Elevated bilirubin levels can be a sign of liver damage or dysfunction, and they can also be a marker of the severity of the underlying liver disease.  Elevated bilirubin levels can affect the treatment options and prognosis for patients with HCC. For example, patients with high bilirubin levels may not be candidates for certain types of treatment, such as liver transplantation, due to the risk of complications. In addition, high bilirubin levels can also be a prognostic factor for survival in patients with HCC, with higher levels being associated with a worse prognosis.  It is important for patients with HCC to work closely with their healthcare team to monitor their bilirubin levels and manage any underlying liver disease. This may involve a combination of treatments, such as chemotherapy, targeted therapies, and surgery, depending on the individual circumstances of the patient. | Similar | 3 | 2 | 3 |
| 51. What is CAR-T cell treatment? Can it be used for hepatocellular carcinoma? (Clinical trial) | CAR-T cell therapy is a type of cancer treatment that involves the use of genetically modified T cells, a type of immune cell, to recognize and attack cancer cells. T cells are a key component of the immune system and play a critical role in helping the body fight infections and other diseases.  In CAR-T cell therapy, T cells are removed from the patient's blood and modified in the laboratory to produce a special protein called a chimeric antigen receptor (CAR). This CAR allows the modified T cells to recognize and bind to a specific protein found on the surface of cancer cells. The modified T cells are then infused back into the patient's bloodstream, where they can specifically target and attack cancer cells bearing the protein to which the CAR is designed to bind.  CAR-T cell therapy has shown promise as a treatment for a number of different types of cancer, including leukemia, lymphoma, and multiple myeloma. However, it is not currently approved for the treatment of hepatocellular carcinoma, which is a type of liver cancer. There is ongoing research into the potential use of CAR-T cell therapy for the treatment of hepatocellular carcinoma, but more studies are needed to determine its safety and effectiveness in this setting. | Similar | 1 | 1 | N/A |
| 52. What if the liver cancer treatment fails? | If liver cancer treatment fails, it may be necessary to try a different treatment approach. This could include a different type of chemotherapy or targeted therapy, surgery, or radiation therapy. In some cases, a combination of treatments may be recommended. It is important to work closely with a medical team to determine the most appropriate treatment plan. It is also important to keep in mind that not all cases of liver cancer can be cured, and the goal of treatment may be to control the cancer and manage symptoms. | If liver cancer treatment fails, it may be necessary to try a different treatment approach or to consider participating in a clinical trial to explore new treatments. It is important to work closely with a healthcare team to determine the best course of action. Options may include a liver transplant, palliative care to manage symptoms, or enrolling in a clinical trial to explore new treatments. It is also important to maintain a healthy lifestyle and manage any underlying medical conditions, as this can help improve quality of life and potentially slow the progression of the disease. | 1,2 | 2,2 | 1,2 |
| **Lifestyle** | | | | | |
| 53. Are herbal medicines recommended to treat liver cancer? | Herbal medicines have not been proven to be effective in the treatment of liver cancer. In general, it is important to be cautious when considering using herbal remedies, as they are not regulated in the same way as prescription medications and may not be safe or effective.  Liver cancer is a serious and potentially life-threatening condition that requires prompt and appropriate medical treatment. If you have liver cancer or are experiencing symptoms that may be related to liver cancer, it is important to speak with a healthcare professional as soon as possible. They will be able to recommend the most appropriate treatment options based on your specific circumstances. | Similar | 1 | 2 | 1 |
| 54. Are there any non-medical therapies to treat liver cancer? | There are several non-medical therapies that may be used to treat liver cancer:  Nutrition therapy: A healthy diet can help improve overall health and support the body's natural healing processes. This may include increasing intake of fruits, vegetables, and whole grains while limiting intake of unhealthy fats and processed foods.  Exercise therapy: Regular exercise has been shown to have numerous health benefits, including improving the immune system and reducing the risk of cancer.  Stress reduction techniques: Stress can have a negative impact on overall health, so finding ways to manage stress can be beneficial. This may include relaxation techniques such as meditation or yoga, or seeking support from friends or family.  Complementary and alternative therapies: There are a number of therapies that are not traditionally considered part of mainstream medical treatment, such as acupuncture, herbal remedies, and chiropractic care. These therapies may be used alongside medical treatment for liver cancer.  It is important to note that non-medical therapies should not be used as a replacement for medical treatment for liver cancer, and should be discussed with a healthcare provider before starting. | Similar | 2 | 2 | N/A |
| 55. What diet should hepatocellular carcinoma patients stick to? | Hepatocellular carcinoma (HCC) is a type of liver cancer that can be treated with a combination of therapies, including surgery, chemotherapy, and radiation. The specific treatment plan for an HCC patient will depend on the stage and severity of the cancer, as well as the overall health of the patient.  Diet can be an important part of the treatment plan for HCC patients. It is important for HCC patients to maintain a well-balanced and nutritious diet, as they may have special nutritional needs due to their cancer and its treatment. Here are some general dietary recommendations for HCC patients:  Eat a variety of nutrient-rich foods: Choose a variety of foods from all food groups, including whole grains, fruits, vegetables, proteins, and healthy fats. These foods can help provide the nutrients you need to stay healthy and fight cancer.  Limit your intake of saturated and trans fats: These types of fats can increase your risk of heart disease and other health problems. Choose lean proteins and plant-based fats instead.  Avoid alcohol: Alcohol can harm the liver and may increase your risk of liver cancer. If you drink alcohol, it is important to limit your intake to reduce your risk of liver cancer and other health problems.  Drink plenty of fluids: It is important to stay hydrated, especially if you are experiencing side effects from your cancer treatment. Aim for at least 8-8 ounces of water per day.  Talk to a registered dietitian: A registered dietitian can help you create a personalized dietary plan that meets your specific needs and takes into account any dietary restrictions or limitations you may have. They can also provide guidance on supplements and other nutrients that may be beneficial for HCC patients.  It is important to remember that every individual is different, and what works for one person may not work for another. It is always a good idea to speak with your healthcare team, including a registered dietitian, before making any changes to your diet. | Similar | 2 | 3 | 2 |
| 56. How can liver cancer be prevented? | There are several ways to reduce the risk of developing liver cancer:  Avoid risky behaviors: Avoiding behaviors that can damage the liver, such as drinking alcohol excessively, using illegal drugs, or engaging in risky sexual practices, can help reduce the risk of liver cancer.  Get vaccinated: Some viruses, such as hepatitis B and hepatitis C, can increase the risk of liver cancer. Getting vaccinated against these viruses can help reduce the risk.  Maintain a healthy weight: Being overweight or obese can increase the risk of liver cancer. Maintaining a healthy weight through diet and exercise can help reduce the risk.  Avoid certain medications: Some medications, such as anabolic steroids, can increase the risk of liver cancer. It is important to discuss the potential risks and benefits of any medications with a healthcare provider before taking them.  Avoid exposure to toxins: Some chemicals and toxins, such as certain pesticides and industrial solvents, can increase the risk of liver cancer. It is important to avoid exposure to these substances whenever possible.  Get screened: If you are at high risk for liver cancer, your healthcare provider may recommend regular screenings to detect the disease early, when it is more treatable.  It is important to note that these measures may not completely eliminate the risk of developing liver cancer, but they can help reduce the risk. It is also important to consult with a healthcare provider for personalized recommendations based on individual risk factors. | Similar | 1 | 2 | 1 |
| 57. Can liver cancer or the treatment affect a person's fertility? | Yes, liver cancer and its treatment can potentially affect a person's fertility. Liver cancer is a type of cancer that starts in the liver, an organ located in the upper right side of the abdomen. The liver is involved in a number of important functions in the body, including the production of hormones and the regulation of blood glucose levels. It also plays a role in the metabolism of drugs and other substances, including those that are involved in fertility.  The treatment of liver cancer, including surgery, chemotherapy, and radiation therapy, can also have an impact on fertility. For example, chemotherapy and radiation therapy can damage the cells that produce eggs or sperm, potentially leading to fertility problems. Surgery to remove part or all of the liver may also affect fertility if it damages the organs or tissues that are involved in fertility.  It is important to discuss fertility concerns with a healthcare provider before starting treatment for liver cancer. In some cases, it may be possible to preserve fertility before treatment begins. There are also options for fertility preservation after treatment, such as freezing eggs or sperm for future use. | Similar | 4 | 2 | 4 |
| 58. Can liver cancer or the treatment affect a person's sex life? | Yes, liver cancer and its treatment can potentially affect a person's sex life.  Cancer and its treatment can often cause physical and emotional changes that can affect a person's sexual functioning and desire. These changes can be related to the cancer itself or to the side effects of treatment, such as chemotherapy, radiation therapy, or surgery.  For example, liver cancer and its treatment can cause fatigue, which can decrease a person's energy levels and affect their desire for sex. Cancer and its treatment can also cause changes in body image, which can affect a person's self-esteem and confidence in their sexual relationships. In addition, certain medications used to treat cancer, such as chemotherapy drugs, can cause side effects such as nausea, vomiting, and diarrhea, which can also affect a person's desire for sex.  It is important for people with liver cancer to discuss any concerns or questions they have about their sex life with their healthcare team. They can offer support and suggest strategies to help manage any sexual side effects that may arise. | Similar | 1 | 2 | 2 |
| 59. Is it possible to shrink liver tumors from food alone without other treatments? | It is not generally possible to shrink liver tumors with food alone without other treatments such as surgery, chemotherapy, or radiation therapy. However, making certain dietary changes may be an important part of your overall treatment plan and can potentially help improve your overall health and well-being.  Some dietary changes that may be beneficial for people with liver tumors include:  Eating a balanced and varied diet that is rich in fruits, vegetables, whole grains, and lean protein sources  Limiting your intake of processed and high-fat foods  Avoiding alcohol, or at least limiting your consumption to a moderate amount  Drinking plenty of fluids, especially water, to help flush toxins out of your body  It is important to note that dietary changes should be discussed with your healthcare team, as they will be able to provide guidance and recommendations based on your specific situation and treatment plan. Additionally, it is important to follow your treatment plan as prescribed by your healthcare team, as this is the most effective way to manage your condition. | It is possible that making certain dietary changes could potentially help to reduce the size of liver tumors, but it is important to note that these changes should be made in conjunction with medical treatment and under the guidance of a healthcare professional. It is not recommended to rely solely on dietary changes as a means of treating liver tumors.  Eating a healthy, balanced diet that is low in fat and high in fruits, vegetables, and whole grains may be beneficial for overall liver health. Maintaining a healthy weight and avoiding tobacco and excessive alcohol consumption may also help to reduce the risk of developing liver tumors.  It is important to talk to a healthcare professional about the best treatment plan for liver tumors, as the appropriate course of action will depend on the specific type and stage of the tumor. Treatment options may include surgery, chemotherapy, radiation therapy, or a combination of these approaches. | 1,2 | 3,3 | 1,4 |
| 60. Do patients with liver cancer need to stop drinking alcohol to prevent liver cancer from worsening? | Yes, patients with liver cancer should stop drinking alcohol to prevent the cancer from worsening. Alcohol can cause further damage to the liver and can interfere with treatment for liver cancer. Additionally, heavy alcohol consumption is a risk factor for the development of liver cancer, so avoiding alcohol can help to reduce the risk of the cancer returning or progressing. It is important for patients with liver cancer to follow their treatment plan and to discuss any lifestyle changes, including alcohol consumption, with their healthcare team. | Similar | 1 | 1 | N/A |
| **Others** | | | | | |
| 61. What are the symptoms that liver cancer patients should look out and go to the emergency room for? | Liver cancer can cause a variety of symptoms, and it's important to consult a medical professional if you experience any unusual or persistent symptoms. Some symptoms that may indicate the need for emergency medical attention include:  Abdominal pain: Pain in the upper right side of the abdomen may be a sign of liver cancer.  Swelling in the abdomen: Liver cancer can cause fluid to build up in the abdomen, which can cause swelling.  Weight loss: Unexpected weight loss can be a symptom of liver cancer.  Jaundice: This is a condition in which the skin and whites of the eyes become yellow. It is often a sign of liver problems.  Nausea and vomiting: These symptoms may be caused by liver cancer or other conditions, but it's important to get them checked out by a medical professional.  Fever: A fever may indicate an infection or other serious condition.  Blood clots: Blood clots can be a serious complication of liver cancer.  It's important to seek medical attention if you experience any of these symptoms, as they may indicate a serious medical condition that requires prompt treatment. | Similar | 2 | 2 | N/A |
| 62. Is cannabidiol (CBD) a good option for pain relief for hepatocellular carcinoma? | There is limited research on the use of cannabidiol (CBD) for pain relief in patients with hepatocellular carcinoma (HCC), which is a type of liver cancer. Some studies have suggested that CBD may have potential as a pain management option, but more research is needed to fully understand its effectiveness and safety for this use.  It is important to note that CBD is not a cure for HCC and should not be used as a replacement for standard medical treatment. If you are experiencing pain due to HCC, it is important to consult with your healthcare provider for appropriate treatment options. They can advise you on the best course of action for managing your pain and overall care. | Similar | 1 | 2 | 2 |
| 63. Can marijuana be used to treat the loss of appetite during hepatocellular carcinoma treatment? | There is some evidence that marijuana, or products containing compounds found in marijuana, may be useful in treating the loss of appetite and other symptoms that can occur during cancer treatment. However, it is important to note that marijuana is not a cure for cancer, and it is not a replacement for standard cancer treatments such as chemotherapy, radiation therapy, or surgery.  One of the active compounds in marijuana, delta-9-tetrahydrocannabinol (THC), has been shown to stimulate appetite and reduce nausea and vomiting in some people. THC is available in a number of forms, including pills, oils, and inhalers, and it can be prescribed by a doctor for use in cancer treatment. However, marijuana is still a controlled substance in many states and countries, and it is not legal for use in all locations.  If you are interested in using marijuana or THC to treat the loss of appetite or other symptoms related to hepatocellular carcinoma treatment, it is important to discuss this with your healthcare provider. They can help you understand the potential risks and benefits of using marijuana in your treatment plan and help you determine if it is a safe and appropriate option for you. | Similar | 1 | 1 | N/A |
| 64. What are the contraindications for undergoing liver transplantation for hepatocellular carcinoma? | There are several contraindications for undergoing liver transplantation for hepatocellular carcinoma (HCC), which is a type of liver cancer. These contraindications may include:  Advanced stage of HCC: The cancer must be at an early stage and not have spread to other parts of the body in order to be eligible for liver transplantation.  Other underlying liver conditions: If the patient has other underlying liver conditions, such as cirrhosis or liver failure, they may not be a candidate for liver transplantation.  Other medical conditions: If the patient has other medical conditions, such as diabetes or heart disease, they may not be a candidate for liver transplantation.  Age: Patients who are older or in poor general health may not be suitable candidates for liver transplantation due to the risk of complications and poor outcomes.  Active infection: Patients with active infections may not be eligible for liver transplantation until the infection has been treated and resolved.  Substance abuse: Patients with a history of substance abuse may not be suitable candidates for liver transplantation due to the risk of relapse and the negative impact on transplant outcomes.  It's important to note that these are just a few of the contraindications for liver transplantation for HCC. A thorough evaluation by a liver transplant team is necessary to determine if liver transplantation is an appropriate treatment option for a particular patient. | Similar | 3 | 2 | 3 |
| 65. How can I help a family member cope with a diagnosis of hepatocellular carcinoma? | Hepatocellular carcinoma (HCC) is a type of liver cancer that can be difficult for a person to cope with. Here are some ways you can support a family member who has been diagnosed with HCC:  Offer emotional support: It's important to be there for your family member and to listen to their feelings and concerns. Let them know that you are there for them and that they are not alone.  Help with practical tasks: Offer to help with tasks such as grocery shopping, transportation to appointments, or household chores. This can help reduce stress and allow your family member to focus on their treatment and recovery.  Encourage healthy habits: Encourage your family member to eat a healthy diet, exercise, and get plenty of rest. These habits can help them feel better physically and emotionally.  Get involved in treatment decisions: Encourage your family member to participate in treatment decisions and to ask their healthcare team questions. It can be helpful to bring a trusted family member or friend to appointments to provide support and to help remember important information.  Find support groups: Look for support groups or online communities for people with HCC. These can be a great source of emotional support and can provide valuable information and resources.  It's important to remember that everyone copes with a diagnosis of HCC differently, so it's important to be patient and understanding. Above all, it's important to be there for your family member and to provide emotional support. | Similar | 1 | 1 | N/A |
| 66. How can I help a family member cope with hepatocellular carcinoma treatments? | HCC stands for hepatocellular carcinoma, which is a type of liver cancer. If you have a family member who is undergoing treatment for HCC, there are several things you can do to help them cope:  Offer emotional support: Being diagnosed with cancer and undergoing treatment can be overwhelming and stressful for your family member. It's important to be there for them and provide emotional support during this time. Let them know that you are there for them and that you care about them.  Help with practical tasks: Your family member may have difficulty completing everyday tasks due to their treatment. Offer to help with tasks such as grocery shopping, cooking, or cleaning.  Provide information and resources: Educate yourself about HCC and the treatment options available. This will help you understand what your family member is going through and provide you with the knowledge to support them. You can also help your family member find resources, such as support groups or financial assistance, that may be helpful during their treatment.  Encourage self-care: Encourage your family member to prioritize self-care during treatment. This can include activities such as exercising, eating a healthy diet, and getting enough rest.  Respect their decisions: It's important to respect your family member's decisions about their treatment and care. Respect their autonomy and allow them to make their own decisions, even if you disagree with them.  It's also important to remember to take care of yourself during this time. It can be emotionally and physically draining to support a loved one through a difficult experience like cancer treatment. Make sure to take breaks and seek support if you need it. | Similar | 1 | 1 | N/A |
| 67. How often is ultrasound done to screen for hepatocellular carcinoma? | Ultrasound is commonly used to screen for hepatocellular carcinoma (HCC), a type of liver cancer, in high-risk populations. The frequency of screening depends on the specific recommendations of a healthcare provider and the individual's risk factors for HCC.  There are several risk factors that may increase a person's likelihood of developing HCC, including a history of chronic liver disease, such as hepatitis B or C infection or cirrhosis, heavy alcohol consumption, and obesity. People with these risk factors may be recommended to undergo ultrasound screenings more frequently to monitor for the development of HCC.  In general, the frequency of ultrasound screenings for HCC is determined by the individual's risk level and the results of previous screenings. For example, if a person has a high risk of developing HCC and has previously had an ultrasound that showed no signs of cancer, they may be recommended to have follow-up ultrasounds every six months to a year. On the other hand, if a person has a lower risk of HCC and has had normal ultrasound results in the past, they may be recommended to have ultrasounds less frequently, such as every two to three years.  It's important to note that ultrasound is only one of several tools that may be used to screen for HCC. Other tests, such as alpha-fetoprotein (AFP) blood tests and liver imaging tests, such as CT scans or MRIs, may also be used to monitor for the development of HCC. | Similar | 3 | 3 | N/A |
| 68. Is high fever with chemotherapy for hepatocellular carcinoma normal? | It is not uncommon for people undergoing chemotherapy to experience side effects, including fever. The severity and specific symptoms can vary from person to person and depend on a variety of factors, including the type and dose of chemotherapy being administered and the individual's overall health.  In general, fever can be a sign of infection, and it is important for people undergoing chemotherapy to monitor their temperature and report any fever or other symptoms to their healthcare team. It is possible that the fever may be related to the chemotherapy or it may be due to another underlying cause. It is important for the healthcare team to determine the cause of the fever so that it can be treated appropriately.  If you are undergoing chemotherapy for hepatocellular carcinoma and are experiencing fever or any other symptoms, it is important to speak with your healthcare team for further guidance and support. They can help determine the cause of the symptoms and provide you with the appropriate treatment. | Similar | 1 | 2 | 2 |
| 69. How to stop/slow hair loss from hepatocellular carcinoma chemotherapy? | Hair loss (alopecia) is a common side effect of chemotherapy for hepatocellular carcinoma (HCC). It is caused by the chemotherapy drugs attacking rapidly dividing cells in the body, including those in the hair follicles. Unfortunately, there is no way to completely prevent hair loss during chemotherapy. However, there are a few things that you can try to help slow or minimize hair loss:  Use a gentle shampoo and avoid styling products: Choose a mild, sulfate-free shampoo and avoid using styling products, such as hair sprays, gels, and mousses, which can be drying and irritating to the scalp.  Protect your scalp: If your scalp is sensitive, you can try wearing a hat or scarf to cover your head and protect it from the sun and cold weather. This can help to prevent further irritation and dryness.  Eat a healthy diet: A healthy diet rich in protein, iron, and other nutrients can help to support the health of your hair. Try to include foods such as eggs, fish, chicken, beans, and leafy green vegetables in your diet.  Use a hair loss prevention product: There are a variety of products available that claim to help prevent hair loss during chemotherapy. These may include topical treatments, such as minoxidil (Rogaine), or supplements, such as biotin or saw palmetto. However, it is important to speak with your doctor before starting any new treatment, as some of these products may interact with your chemotherapy medications or have other potential side effects.  Consider a wig or hairpiece: If your hair loss is severe, you may want to consider wearing a wig or hairpiece to help cover your head and boost your confidence. There are many different styles and materials available, so you can choose one that best suits your needs and preferences.  It is also important to remember that hair loss during chemotherapy is usually temporary, and your hair will typically begin to regrow a few months after treatment is completed. If you have any concerns about your hair loss, be sure to discuss them with your doctor or a healthcare professional. | Similar | 1 | 2 | 2 |
| 70. Is it safe to get COVID vaccines when undergoing hepatocellular carcinoma treatment? | It is generally safe for people undergoing treatment for hepatocellular carcinoma (HCC) to receive COVID-19 vaccines. HCC is a type of liver cancer, and people with liver cancer can be at higher risk of serious illness from COVID-19. Therefore, it is especially important for people with HCC to be vaccinated against COVID-19 to reduce their risk of severe illness from the virus.  If you are currently undergoing treatment for HCC, it is important to discuss the COVID-19 vaccine with your healthcare provider. They can advise you on the best course of action based on your specific situation and the type of treatment you are receiving. In general, it is safe to receive most vaccines while undergoing cancer treatment, but your healthcare provider will be able to provide more specific guidance. | Similar | 1 | 2 | 1 |
| 71. Should patients wait to cure Hepatitis C (HCV) before or after treatment for hepatocellular carcinoma? | It is generally recommended to treat hepatitis C (HCV) before treating hepatocellular carcinoma (HCC). This is because HCV is a leading cause of HCC, and treating the HCV can help prevent the development or progression of HCC. Additionally, HCV treatment can improve the effectiveness of HCC treatment. It is important for patients with HCV and HCC to work with their healthcare team to determine the most appropriate treatment plan. | It is generally recommended to treat hepatitis C (HCV) before treating hepatocellular carcinoma (HCC). This is because HCV is a major risk factor for the development of HCC, and treating the HCV can help to prevent the progression of liver damage and the recurrence of HCC. In addition, treating HCV can improve the overall prognosis for patients with HCC. It is important for patients with HCV to discuss their treatment options with a healthcare provider and make a plan that is appropriate for their specific situation. | 2 | 2 | N/A |
| 72. Are there any herbal meds that can be used to help prevent liver cancer from worsening? | There is no evidence to suggest that any herbal remedies can prevent liver cancer from worsening. Liver cancer is a serious and potentially life-threatening condition that requires medical treatment. If you are concerned about liver cancer, it is important to speak with a healthcare professional. They can discuss your treatment options and help you develop a plan for managing your condition. | There is no evidence to suggest that any particular herbs or dietary supplements can prevent liver cancer from worsening. It is important to note that liver cancer is a serious and potentially life-threatening condition that requires medical treatment. If you have liver cancer or are at risk for developing it, it is important to speak with your doctor about your treatment options. In some cases, surgery, chemotherapy, and/or radiation therapy may be recommended to treat liver cancer. It is also important to follow a healthy lifestyle, including maintaining a healthy weight, not smoking, and limiting your alcohol intake, as these factors may help to reduce your risk of developing liver cancer. | 1,1 | 2,1 | 1,1 |
| 73. Does milk thistle help with liver cancer? | Milk thistle is a plant that has been traditionally used to support liver health and is thought to have liver-protective properties. Some studies have suggested that milk thistle may have potential as a treatment for liver cancer, but more research is needed to confirm this. It is important to note that milk thistle should not be used as a replacement for standard medical treatment for liver cancer. If you have liver cancer or are concerned about your liver health, you should talk to a healthcare provider. | Milk thistle is a plant that has been traditionally used for liver health. Some studies have suggested that milk thistle may have liver-protective effects and may be beneficial for people with liver conditions such as cirrhosis and hepatitis. However, there is not enough scientific evidence to support the use of milk thistle for the treatment of liver cancer.  It is important to note that liver cancer is a serious condition that requires medical treatment. If you are concerned about liver cancer, it is important to speak with a healthcare professional for a proper diagnosis and treatment plan. | 1,1 | 2,1 | 2,1 |

### Supplemental Table 3. Grading of the responses from the ChatGPT Language Model for the questions related to cirrhosis management quality measure

| Questions | Responses 1 | Responses 2 | Reviewer Grade  1. Comprehensive  2. Correct but inadequate  3. Mixed with correct and incorrect/outdated data 4. Completely incorrect |
| --- | --- | --- | --- |
| 1. When should patients with ascites who are admitted to the hospital for evaluation and management of symptoms related to ascites or encephalopathy receive a diagnostic paracentesis? | A diagnostic paracentesis is typically performed in patients with ascites who are experiencing symptoms related to the accumulation of fluid in the abdomen, such as abdominal pain, difficulty breathing, or feeling full or bloated. The procedure may also be performed in patients who have developed hepatic encephalopathy, a condition in which the accumulation of toxins in the blood can cause changes in mental status, including confusion, altered levels of consciousness, and coma. A diagnostic paracentesis may be performed as part of the initial evaluation of these patients when they are admitted to the hospital to help determine the cause of the ascites or encephalopathy and guide further treatment. It is important to note that the timing and indications for a diagnostic paracentesis may vary depending on the specific patient and the clinical circumstances. | Similar | 1/2 |
| 2. Patients who are admitted with or develop GI bleeding should receive antibiotics within how many hours? What is the minimum duration for the course of antibiotics? | The timing and duration of antibiotic treatment for gastrointestinal (GI) bleeding depend on the underlying cause of the bleeding and the patient's individual circumstances. In general, antibiotics should be administered as soon as possible after the diagnosis of GI bleeding is made. There is no specific time frame for initiating antibiotic treatment, as the decision to use antibiotics should be based on the patient's clinical presentation and the suspected or confirmed cause of the bleeding.  As for the duration of antibiotic treatment, it is typically at least 3-5 days, but may be longer depending on the severity of the infection and the patient's response to treatment. It is important to continue antibiotics for the full course of treatment as prescribed by a healthcare provider, even if the patient's symptoms improve, to ensure that the infection is fully cleared. | It is generally recommended that patients who are admitted with or develop gastrointestinal (GI) bleeding should receive antibiotics within 24 hours. The minimum duration for the course of antibiotics will depend on the specific type of infection present and the patient's response to treatment. In general, antibiotics should be given for at least 3-5 days, and sometimes longer if the infection is severe or if the patient is not responding well to treatment. It is important to follow the recommendations of the healthcare provider in charge of the patient's care, as the specific treatment plan may vary depending on the individual case | 3 |
| 3. What is the minimum amount of ascites removed from paracentesis in patients with cirrhosis for intravenous albumin to be indicated? | Intravenous albumin is usually indicated in patients with cirrhosis and ascites who have had at least 5 liters of ascites removed through paracentesis. This is based on the finding that patients with large-volume ascites (> 5 liters) who have had at least this amount of fluid removed are at increased risk of developing hypovolemia, or low blood volume. In these patients, intravenous albumin can be used to help restore and maintain normal blood volume. However, the decision to use intravenous albumin should be made on a case-by-case basis, taking into account the patient's overall clinical condition and any other relevant factors. | It is generally recommended that patients with cirrhosis and ascites who have had more than 5 liters of ascites removed during paracentesis within the previous 3 months, or more than 6 liters removed within the previous 6 months, receive intravenous albumin to help prevent further fluid accumulation and reduce the risk of complications such as hepatorenal syndrome. However, the decision to administer intravenous albumin should be made on a case-by-case basis, taking into account the patient's overall clinical condition and the presence of other risk factors for fluid accumulation. It is important to note that intravenous albumin should not be used as a substitute for proper management of the underlying liver disease, and that other measures such as salt and water restriction, diuretic therapy, and antibiotic prophylaxis may also be necessary to prevent recurrent ascites. | 1/2 or 3 |
| 4. In hospitalized patients with cirrhosis and ascites, what is the threshold ascitic fluid polymorphonuclear count to start empiric antibiotics and albumin? What is the dose of albumin given in this scenario? | In hospitalized patients with cirrhosis and ascites, empiric antibiotic treatment is generally recommended for those with an ascitic fluid polymorphonuclear (PMN) count of at least 250 cells/mm^3. This is based on the finding that patients with an ascitic fluid PMN count of 250 cells/mm^3 or higher are at increased risk of developing spontaneous bacterial peritonitis (SBP), a serious complication of cirrhosis that requires prompt treatment with antibiotics.  If intravenous albumin is used in conjunction with antibiotics for the treatment of SBP, the usual dose is 1.5 g/kg body weight per day, given in divided doses. For example, a 70 kg patient would receive 105 g of albumin per day. It is important to note that the use of intravenous albumin in the treatment of SBP is controversial, and some studies have suggested that it may not provide any additional benefit beyond that of antibiotic treatment alone. The decision to use intravenous albumin should be made on a case-by-case basis, taking into account the patient's overall clinical condition and any other relevant factors. | Similar | 1/2 |
| 5. Should patients with ascites and/or hepatic hydrothorax be given both sodium restriction and diuretics? | Patients with ascites and/or hepatic hydrothorax may benefit from both sodium restriction and diuretics. Sodium restriction can help to reduce the amount of fluid accumulation in the body, while diuretics can help to increase the amount of urine produced and promote the elimination of excess fluid. However, the specific treatment plan will depend on the underlying cause of the fluid accumulation, as well as the patient's overall health status and other medical conditions. It is important for patients with ascites and/or hepatic hydrothorax to work closely with their healthcare team to determine the most appropriate treatment approach. | Similar | 1/2 |
| 6. Should patients who undergo paracentesis receive fresh frozen plasma or platelets? | Paracentesis is a procedure in which a needle is used to remove fluid from the abdomen. The decision to administer fresh frozen plasma or platelets to a patient who has undergone paracentesis will depend on the specific circumstances of the case and the patient's overall health and medical history.  In general, fresh frozen plasma (FFP) may be administered to patients who have lost a significant amount of blood or have a deficiency of clotting factors. Platelets may be given to patients who have a low platelet count or who are at risk of bleeding.  It is important to note that FFP and platelets are blood products that are derived from donated blood. As such, they carry a risk of allergic reactions and other adverse effects. Therefore, these products should be administered only when the potential benefits outweigh the potential risks.  If you have undergone paracentesis and have concerns about whether you should receive FFP or platelets, it is best to consult with your healthcare provider. They will be able to assess your specific needs and determine the most appropriate course of treatment. | Similar | 3 |
| 7. In patients with cirrhosis but no documentation of previous GI bleeding, what is the cut-off platelet count and liver stiffness measurement for indication of upper endoscopy to screen for varices? | The cut-off platelet count for indication of upper endoscopy to screen for varices in patients with cirrhosis but no documentation of previous GI bleeding is typically a platelet count of less than 50,000/microliter. The cut-off liver stiffness measurement for indication of upper endoscopy in this population is typically a liver stiffness measurement of greater than 15 kPa. It is important to note that these cut-off values may vary based on individual patient factors and clinical judgment. It is always best to consult with a healthcare provider for specific recommendations. | There are different guidelines for the management of patients with cirrhosis and the indication for upper endoscopy to screen for varices. The cut-off platelet count and liver stiffness measurement for the indication of upper endoscopy can vary depending on the specific guidelines and the clinical context.  In general, patients with cirrhosis and a platelet count of less than 50,000/mm3 are at high risk of bleeding from varices and may benefit from upper endoscopy to screen for and treat varices. In addition, patients with cirrhosis and a liver stiffness measurement (as measured by transient elastography) of more than 20-25 kPa are also at high risk of bleeding from varices and may benefit from upper endoscopy.  It is important to note that these are general guidelines and the decision to perform upper endoscopy in a specific patient with cirrhosis should be based on a thorough assessment of the patient's individual risk factors and clinical presentation. It is also important to consult with a gastroenterologist or hepatologist for further guidance on the management of patients with cirrhosis. | 3 |
| 8. Should patients with decompensated cirrhosis and no documented history of previous GI bleeding receive upper endoscopy to screen for varices? If yes, the upper endoscopy be performed within how many months of diagnosis? | Yes, patients with decompensated cirrhosis and no documented history of previous GI bleeding should receive upper endoscopy to screen for varices. This is because patients with decompensated cirrhosis are at high risk for developing varices, which are dilated blood vessels in the lining of the esophagus or stomach that can rupture and cause serious bleeding. Upper endoscopy allows for the visualization and assessment of these blood vessels, and can help to identify those at high risk for bleeding.  The upper endoscopy should be performed within 3-6 months of the diagnosis of decompensated cirrhosis. This is because the risk of variceal bleeding increases with the severity of liver disease, and patients with decompensated cirrhosis are at particularly high risk. Early identification and management of varices can help to prevent serious bleeding events and improve outcomes for these patients. | It is generally recommended that patients with decompensated cirrhosis undergo upper endoscopy to screen for esophageal varices. This is because patients with cirrhosis are at an increased risk of developing esophageal varices, which are enlarged veins in the esophagus that can rupture and cause life-threatening bleeding.  The timing of the upper endoscopy may vary depending on the clinical context and the individual patient's circumstances. In general, it is recommended that patients with decompensated cirrhosis undergo upper endoscopy within 6-12 months of diagnosis. However, some guidelines recommend that the procedure be performed sooner in patients with more advanced cirrhosis or in those who have other risk factors for variceal bleeding, such as a high Child-Pugh score or a history of variceal bleeding.  It is important to note that the decision to perform an upper endoscopy in a particular patient should be made on a case-by-case basis, taking into account the patient's overall clinical condition and the potential risks and benefits of the procedure. | 3 |
| 9. What treatment or medication should be given to patients with cirrhosis, no documented history of previous GI bleeding, and medium/large varices on endoscopy? Within how many months the treatment should be given? | If a patient with cirrhosis has medium or large varices on endoscopy, it is generally recommended to start prophylactic treatment to prevent the varices from bleeding. This is typically done with a combination of medications called beta blockers and band ligation. Beta blockers work by reducing the pressure in the veins in the esophagus, which helps to prevent the varices from bleeding. Band ligation involves placing a rubber band around the base of the varix, which cuts off the blood supply to the vein and causes it to shrink. This treatment should be initiated as soon as possible, ideally within a few weeks of the endoscopy. If the patient has a history of bleeding from varices, or if the varices are very large or have already bled, more aggressive treatment may be necessary. This may include medications to reduce portal pressure, such as non-selective beta blockers or nitrates, or procedures to destroy the varices, such as sclerotherapy or transjugular intrahepatic portosystemic shunt (TIPS). | Similar | 1/2 |
| 10. Should patients with cirrhosis who present with upper GI bleeding receive upper endoscopy? If yes, within how many hours of presentation should the upper endoscopy be done? | Yes, patients with cirrhosis who present with upper gastrointestinal (GI) bleeding should generally receive upper endoscopy. Upper endoscopy is a procedure that allows a doctor to examine the inside of the upper digestive tract, including the esophagus, stomach, and duodenum. It can be used to identify the source of bleeding and determine the best treatment approach.  In general, it is recommended that upper endoscopy be performed as soon as possible in patients with upper GI bleeding, as prompt diagnosis and treatment can be important for reducing the risk of complications. However, the specific timing will depend on the patient's clinical condition and the availability of endoscopy services. It is generally recommended that upper endoscopy be performed within 24 hours of presentation in patients with upper GI bleeding. However, in cases where the patient is stable and there is a delay in obtaining endoscopy, supportive care can be provided while waiting for the procedure. | Similar | 3 |
| 11. When should patients with cirrhosis who are found to have bleeding esophageal varices during endoscopy receive EVL or sclerotherapy? | Patients with cirrhosis who are found to have bleeding esophageal varices during endoscopy should receive either EVL (endoscopic variceal ligation) or sclerotherapy as soon as possible. This is because bleeding esophageal varices can be life-threatening and require immediate treatment to stop the bleeding and prevent further complications. In some cases, patients may be given a prophylactic treatment of EVL or sclerotherapy to prevent bleeding from occurring in the future. The decision to use EVL or sclerotherapy will depend on the severity of the bleeding, the patient's overall health, and any other underlying medical conditions. | Similar | 1/2 |
| 12. What combination of treatment should be given to patients with cirrhosis who survive an episode of acute variceal hemorrhage? | The combination of treatment for patients with cirrhosis who survive an episode of acute variceal hemorrhage should include:  Endoscopic therapy to stop the bleeding and prevent further bleeding, such as band ligation or sclerotherapy.  Prophylactic measures to prevent future episodes of variceal hemorrhage, such as beta blockers or nitrates.  Management of underlying cirrhosis, such as lifestyle changes (such as avoiding alcohol) and medications to reduce portal hypertension and improve liver function.  Treatment of any underlying causes of cirrhosis, such as viral hepatitis or autoimmune liver diseases.  Close monitoring and follow-up to ensure that the patient's condition is stable and to identify any potential complications or new episodes of bleeding. | Similar | 1/2 |
| 13. Should Patients with previous overt hepatic encephalopathy be counseled regarding the risks associated with driving? Why? | Yes, patients with previous overt hepatic encephalopathy should be counseled regarding the risks associated with driving. Hepatic encephalopathy is a condition that causes changes in mental function and behavior due to liver failure or other liver-related issues. These changes can affect a person's ability to safely operate a vehicle.  Driving requires cognitive and motor skills that may be impaired in individuals with hepatic encephalopathy. This can increase the risk of accidents and injuries for both the patient and other road users. In addition, hepatic encephalopathy can cause fatigue and drowsiness, which can also affect a person's ability to safely operate a vehicle.  Counseling patients about the risks associated with driving can help them make informed decisions about their ability to drive safely. It is important for patients to understand the potential consequences of driving while impaired and to seek alternative transportation options if necessary. | Similar | 1/2 |
| 14. Should the physician perform a search for evidence of precipitating factors documented in the chart for patients with hepatic encephalopathy? | Yes, it is important for the physician to perform a search for evidence of precipitating factors in patients with hepatic encephalopathy. Hepatic encephalopathy is a condition that occurs when there is a disturbance in the normal functioning of the brain due to liver disease. Precipitating factors are events or conditions that can trigger or worsen hepatic encephalopathy. Examples of precipitating factors include gastrointestinal bleeding, infections, dehydration, electrolyte imbalances, and certain medications. Identifying and addressing these factors can help to improve the patient's symptoms and prevent further deterioration of their condition. | Similar | 1/2 |
| 15. What medication should be given to patients who are hospitalized and have an acute episode of overt hepatic encephalopathy? | Acute episodes of overt hepatic encephalopathy are a medical emergency that require immediate treatment. The main goals of treatment are to remove the precipitating factors, such as constipation or infection, and to improve the function of the liver.  Treatment may include medications such as lactulose, a laxative that helps to reduce the levels of toxic substances in the blood by promoting the removal of waste products through the intestines, and antibiotics to treat any infections. Other medications that may be used to treat hepatic encephalopathy include neomycin, rifaximin, and metronidazole.  It is important to note that the specific treatment plan will depend on the underlying cause of the hepatic encephalopathy and the overall health of the patient. It is important to work closely with a healthcare team to determine the most appropriate treatment approach. | Similar | 1/2 |
| 16. What prophylactic medication should be given to patients who are discharged after an acute episode of hepatic encephalopathy? | Prophylactic medication is often prescribed to patients who have had an acute episode of hepatic encephalopathy in order to prevent future episodes from occurring. The most commonly used prophylactic medications for hepatic encephalopathy are lactulose and rifaximin. Lactulose is a laxative that helps to decrease the levels of toxins in the blood by increasing the frequency of bowel movements and reducing the absorption of toxins in the intestine. Rifaximin is an antibiotic that targets bacteria in the intestine and helps to reduce the production of ammonia, a common cause of hepatic encephalopathy. It is important to follow the recommendations of a healthcare provider regarding the use of prophylactic medication and to take the medication as directed in order to reduce the risk of future episodes of hepatic encephalopathy. | Similar | 1/2 |
| 17. For patients with cirrhosis who do not have absolute contraindications to liver transplantation, what is the cut-off for MELD-score to indicate an evaluation for liver transplantation | The Model for End-Stage Liver Disease (MELD) score is a numerical scale that is used to assess the severity of liver disease and predict the risk of mortality in patients with cirrhosis. The MELD score is calculated using three laboratory values: serum creatinine, bilirubin, and INR (international normalized ratio). The score ranges from 6 to 40, with a higher score indicating more severe liver disease.  For patients with cirrhosis who do not have absolute contraindications to liver transplantation, the cut-off for MELD score to indicate an evaluation for liver transplantation varies depending on the transplant center and the availability of donor organs. In general, patients with a MELD score of 20 or higher are considered to be candidates for liver transplantation. However, some transplant centers may consider patients with a MELD score lower than 20 if they have other factors that suggest a high risk of mortality without a transplant. It is important to note that the MELD score is just one factor that is considered in the evaluation for liver transplantation, and other factors such as the patient's overall health, age, and cause of liver disease are also taken into account. | Similar | 3 |
| 18. Should Patients with cirrhosis, who do not have absolute contraindications to liver transplantation, have hepatocellular carcinoma (HCC) meeting the transplant criteria, be considered for liver transplantation regardless of Model for End-Stage Liver Disease (MELD) score? | Yes, patients with cirrhosis and HCC who do not have absolute contraindications to liver transplantation should be considered for liver transplantation regardless of MELD score if they meet the transplant criteria for HCC. MELD scores are used to prioritize liver transplantation for patients with end-stage liver disease, but they do not take into account specific conditions such as HCC. Therefore, patients with HCC who meet the transplant criteria should not be denied a transplant solely based on their MELD score. | Similar | 1/2 |
| 19. What is the time interval for hepatocellular carcinoma (HCC) screening using abdominal imaging with or without serum α-fetoprotein in patients with cirrhosis? | The recommended time interval for hepatocellular carcinoma (HCC) screening using abdominal imaging with or without serum alpha-fetoprotein (AFP) in patients with cirrhosis depends on the specific guidelines being followed and the individual patient's risk factors for HCC.  The American Association for the Study of Liver Diseases (AASLD) recommends that patients with cirrhosis undergo HCC surveillance with abdominal ultrasound every 6 months, along with serum AFP testing every 6 months. However, if a patient has a high risk of developing HCC (e.g., due to a history of heavy alcohol consumption or hepatitis B infection), the AASLD recommends more frequent imaging (e.g., every 3-4 months) and serum AFP testing.  The European Association for the Study of the Liver (EASL) recommends similar HCC surveillance intervals for patients with cirrhosis. However, the EASL also suggests that patients with a low risk of developing HCC (e.g., due to non-alcoholic fatty liver disease or hepatitis C infection) may be able to undergo HCC surveillance every 12-24 months.  It is important to note that these are general guidelines and the appropriate HCC surveillance interval for a particular patient may vary based on their individual circumstances. It is always best to discuss the appropriate HCC screening intervals with a healthcare provider. | Similar | 1/2 |
| 20. Should patients with cirrhosis have hepatitis B immune status and/or vaccination be documented in the chart? | Yes, it is important for patients with cirrhosis to have their hepatitis B immune status and vaccination documented in the chart. Hepatitis B is a viral infection that can cause liver inflammation and damage, and it is one of the leading causes of cirrhosis and liver cancer. In patients with cirrhosis, it is especially important to prevent hepatitis B infection, as it can lead to further deterioration of the liver and increase the risk of death.  The hepatitis B vaccine is a safe and effective way to prevent hepatitis B infection. The vaccine is recommended for all adults who are at risk of hepatitis B, including patients with cirrhosis. It is important for patients with cirrhosis to have their hepatitis B immune status documented in the chart, as this will help to determine whether they need to be vaccinated or not. If a patient has already been vaccinated, it is important to document this in the chart as well, as this will help to ensure that they receive booster doses as needed. | Similar | 1/2 |
| 21. What treatment should be considered for patients with untreated hepatitis C cirrhosis? | Treatment for patients with untreated hepatitis C cirrhosis typically involves the use of antiviral medications to help clear the hepatitis C virus from the body. The goal of treatment is to halt or slow the progression of liver damage and prevent complications such as liver failure or liver cancer.  There are several antiviral medications that are effective in treating hepatitis C, including:  Interferon-based therapies (such as pegylated interferon)  Direct-acting antivirals (DAAs)  Combination therapies that use a combination of interferon and DAAs  The specific treatment regimen recommended for a patient with untreated hepatitis C cirrhosis will depend on several factors, including the type of hepatitis C virus they have, the severity of their liver damage, and any other medical conditions they may have. It is important for patients to work closely with their healthcare team to determine the most appropriate treatment plan for their specific needs. | Similar | 1/2 |
| 22. What treatment should be considered for Patients with untreated hepatitis B cirrhosis? | It is generally recommended that patients with untreated hepatitis B cirrhosis receive antiviral therapy to slow the progression of liver damage and reduce the risk of developing liver cancer. The most commonly used antiviral medications for hepatitis B are nucleos(t)ide analogues, which work by inhibiting the hepatitis B virus's ability to replicate. These medications can be taken orally and are generally well-tolerated, although they may cause side effects such as headache, fatigue, and nausea. It is important for patients to take these medications exactly as prescribed and to continue treatment for the recommended duration in order to achieve the maximum benefit. In addition to antiviral therapy, patients with hepatitis B cirrhosis may also need to receive other treatments such as liver transplantation or other supportive care measures. | Similar | 1/2 |
| 23. Patients with cirrhosis should receive counseling or be referred to a substance abuse treatment program within how many months of positive screening? | It is generally recommended that patients with cirrhosis who screen positive for substance abuse receive counseling or be referred to a substance abuse treatment program as soon as possible. There is no specific timeframe within which this should occur, as the need for treatment may vary depending on the individual's specific circumstances and the severity of their substance abuse. In general, however, it is important for patients with cirrhosis to receive appropriate treatment as soon as possible to help prevent further liver damage and improve their overall health. | Similar | 2 |
| 24. Should Patients with cirrhosis who are undergoing abdominal surgery have documentation of the risk–benefit of undergoing the surgical procedure in the medical record? | Yes, it is important to document the risk-benefit of undergoing surgery in the medical record for patients with cirrhosis or any other underlying medical condition. This documentation can help to inform the decision-making process and ensure that the patient is aware of the potential risks and benefits of the procedure. It is also important to carefully consider the potential effects of the surgery on the patient's underlying condition and to have a plan in place to manage any potential complications that may arise. | Similar | 2 |
| 25. Recently discharged patients with cirrhosis should have a clinic visit with a health care provider within how many weeks of discharge? | Recently discharged patients with cirrhosis should have a clinic visit with a health care provider within 1-2 weeks of discharge. This is to ensure that the patient's condition is stable and to address any issues or concerns that may have arisen since the patient was discharged from the hospital. It is important for patients with cirrhosis to have close follow-up with their health care provider to monitor the progression of their disease and to manage any complications that may arise. | Similar | 2 |
| 26. Patients with cirrhosis should be assessed for frailty using what method? | There are several ways to assess frailty in patients with cirrhosis. One commonly used method is the Fried frailty criteria, which includes five criteria: weight loss, weakness, low physical activity, slow walking speed, and exhaustion.  Another method is the Clinical Frailty Scale (CFS), which categorizes patients into one of nine levels of frailty, ranging from very fit to very frail. The CFS takes into account factors such as mobility, cognition, nutrition, and co-morbidities.  Both the Fried frailty criteria and the CFS are simple and easy to use, and can provide useful information about a patient's frailty status and prognosis. It is important to note that frailty can be reversible, and interventions such as physical therapy and rehabilitation can improve a patient's frailty status. | Similar | 2 |

### Supplemental Table 4A. Grading of the responses from the ChatGPT Language Model for the published HCC surveillance knowledge questionnaires

#### Article title: Physician- and patient-reported barriers to hepatocellular carcinoma surveillance: A nationwide survey

| Questions | Responses 1 | Responses 2 | Reviewer Grade  1. Comprehensive  2. Correct but inadequate  3. Mixed with correct and incorrect/outdated data 4. Completely incorrect |
| --- | --- | --- | --- |
| In your opinion, which of the following patients are high-risk groups for hepatocellular carcinoma and should undergo surveillance? (Select all that apply.)  1. Patient with cirrhosis  2. All patients with chronic HBV infection  3. Patient with chronic HBV infection who fulfills indication for HCC surveillance  4. All patients with chronic HCV infection  5. Patient with chronic HCV infection who fulfills indication for HCC surveillance  6. None are at risk | Patient with cirrhosis  All patients with chronic HBV infection  All patients with chronic HCV infection  Patient with chronic HCV infection who fulfills indication for HCC surveillance  Patients with cirrhosis and those with chronic HBV or HCV infection are considered high-risk groups for hepatocellular carcinoma (HCC) and should undergo surveillance. Additionally, patients with chronic HBV or HCV infection who fulfill the indications for HCC surveillance should also undergo surveillance. | Patient with cirrhosis  All patients with chronic HBV infection  All patients with chronic HCV infection  Patient with chronic HCV infection who fulfills indication for HCC surveillance | 3 |
| What is/are the optimal tool(s) for HCC surveillance? (Choose only one answer.)  1. Upper abdominal ultrasonography only  2. Serum alpha-fetoprotein only  3. Upper abdominal ultrasonography as the main screening tool with serum alpha-fetoprotein  as an optional tool  4. Upper abdominal ultrasonography or serum alpha-fetoprotein  5. Upper abdominal ultrasonography and serum alpha-fetoprotein  6. CT or MRI of upper abdomen  7. Ultrasonography or CT or MRI of upper abdomen | The optimal tool for HCC surveillance is Upper abdominal ultrasonography and serum alpha-fetoprotein (Option 5) as they are considered to be the most effective and efficient tools for HCC surveillance. It is recommended to use both tools in combination to increase the sensitivity and specificity of HCC detection. | The optimal tool for HCC surveillance is Upper abdominal ultrasonography and serum alpha-fetoprotein (Option 5) as they are considered to be the most effective and efficient tools for HCC surveillance. It is recommended to use both tools in combination to increase the sensitivity and specificity of HCC detection. | 1/2 |
| Which is the recommended interval for HCC surveillance? (Choose only one answer.)  1. Every 3 months  2. Every 6 months  3. Every 6-12 months  4. Every 12 months  5. More than 12 months  6. Only in symptomatic patients | The recommended interval for HCC surveillance is Every 6 months (Option 2) for high-risk patients such as those with cirrhosis or chronic hepatitis B or C. This interval is considered as the most appropriate to detect HCC at an early stage and to improve the chances of survival. It is important to note that the interval may vary depending on the individual patient's risk factors and the stage of the disease. | The recommended interval for HCC surveillance is Every 6-12 months (Option 3) for high-risk patients such as those with cirrhosis or chronic hepatitis B or C. This interval is considered as the most appropriate to detect HCC at an early stage and to improve the chances of survival. It is important to note that the interval may vary depending on the individual patient's risk factors and the stage of the disease, and that regular monitoring and follow up with the treating physician is important to determine the best surveillance interval. | 1^st^ response: 2  2^nd^ response: 3 |
| What screening test should be done with patients with Compensated cirrhosis (no ascites and history of variceal bleeding)  A. Upper abdominal ultrasonography only  B. Serum alpha-fetoprotein only  C. Upper abdominal ultrasonography as the main screening tool with serum alphafetoprotein as an optional tool  D. Upper abdominal ultrasonography or serum alpha-fetoprotein  E. Both upper abdominal ultrasonography and serum alpha-fetoprotein  F. CT or MRI of upper abdomen  G. The patient has no indication for HCC surveillance. | E. Both upper abdominal ultrasonography and serum alpha-fetoprotein  Compensated cirrhosis is a stage of liver disease in which the liver is damaged and not functioning optimally, but the person does not have symptoms or complications such as ascites or variceal bleeding. It is important to monitor patients with compensated cirrhosis for the development of complications and to identify liver cancer (hepatocellular carcinoma, HCC) early, as it can be cured if caught at an early stage. Both upper abdominal ultrasonography and serum alpha-fetoprotein are recommended as screening tests for HCC in patients with compensated cirrhosis. Other imaging tests, such as CT or MRI, may also be used. | E. Both upper abdominal ultrasonography and serum alpha-fetoprotein are recommended as screening tests for patients with compensated cirrhosis. Ultrasonography is the primary screening tool, as it is noninvasive and can detect small hepatocellular carcinomas (HCCs) that may not be detectable by other means. Serum alpha-fetoprotein is a secondary screening tool that can be used to confirm the diagnosis of HCC. The combination of these two tests is the most effective way to detect HCC in patients with compensated cirrhosis. | 1/2 |
| What screening test should be done with patients with Cirrhosis with ascites  A. Upper abdominal ultrasonography only  B. Serum alpha-fetoprotein only  C. Upper abdominal ultrasonography as the main screening tool with serum alphafetoprotein as an optional tool  D. Upper abdominal ultrasonography or serum alpha-fetoprotein  E. Both upper abdominal ultrasonography and serum alpha-fetoprotein  F. CT or MRI of upper abdomen  G. The patient has no indication for HCC surveillance. | E. Both upper abdominal ultrasonography and serum alpha-fetoprotein are recommended as screening tests for patients with cirrhosis and ascites. Upper abdominal ultrasonography is a non-invasive imaging test that uses sound waves to produce an image of the organs in the upper abdomen, including the liver, gallbladder, and pancreas. It is a useful tool for detecting liver abnormalities, such as liver cancer. Serum alpha-fetoprotein (AFP) is a protein produced by the liver that can be measured in the blood. Elevated levels of AFP can be a sign of liver cancer. Therefore, both tests can be useful in detecting liver cancer in patients with cirrhosis and ascites. | The correct answer is E. Both upper abdominal ultrasonography and serum alpha-fetoprotein should be done as screening tests for patients with cirrhosis with ascites. Ultrasonography is a non-invasive, cost-effective method for detecting hepatocellular carcinoma (HCC), which is a common complication of cirrhosis. Serum alpha-fetoprotein (AFP) is a protein produced by liver cells and is elevated in the blood of patients with HCC. Both ultrasonography and AFP are recommended as part of a surveillance program for HCC in patients with cirrhosis. | 4 |
| What screening test should be done with patients with chronic HBV infection with first degree relative of HCC  A. Upper abdominal ultrasonography only  B. Serum alpha-fetoprotein only  C. Upper abdominal ultrasonography as the main screening tool with serum alphafetoprotein as an optional tool  D. Upper abdominal ultrasonography or serum alpha-fetoprotein  E. Both upper abdominal ultrasonography and serum alpha-fetoprotein  F. CT or MRI of upper abdomen  G. The patient has no indication for HCC surveillance. | The correct answer is E: Both upper abdominal ultrasonography and serum alpha-fetoprotein.  In patients with chronic hepatitis B virus (HBV) infection, the risk of developing hepatocellular carcinoma (HCC) is increased. Therefore, it is important to regularly monitor these patients for the development of HCC. Upper abdominal ultrasonography and serum alpha-fetoprotein are both recommended as screening tools for HCC in patients with chronic HBV infection. Upper abdominal ultrasonography is a non-invasive imaging test that uses sound waves to create a picture of the liver and surrounding organs. Serum alpha-fetoprotein is a blood test that measures the levels of a protein called alpha-fetoprotein in the blood. Elevated levels of alpha-fetoprotein can be a sign of liver cancer.  It is important to note that having a first-degree relative with HCC may also increase the risk of developing HCC, so it is important for these patients to undergo regular HCC surveillance as well. | The correct answer is E: Both upper abdominal ultrasonography and serum alpha-fetoprotein.  Chronic hepatitis B virus (HBV) infection is associated with an increased risk of developing hepatocellular carcinoma (HCC), which is a type of liver cancer. Therefore, patients with chronic HBV infection and a first-degree relative with HCC should undergo HCC surveillance.  Upper abdominal ultrasonography and serum alpha-fetoprotein are both commonly used for HCC surveillance in patients with chronic HBV infection. Upper abdominal ultrasonography is a non-invasive imaging test that uses high-frequency sound waves to produce images of the liver, gallbladder, and other organs in the upper abdomen. Serum alpha-fetoprotein is a protein produced by liver cells that can be detected in the blood. Elevated levels of alpha-fetoprotein may be a sign of liver damage or cancer.  CT or MRI of the upper abdomen can also be used for HCC surveillance, but they are generally not the first choice because they expose the patient to ionizing radiation and are more expensive than ultrasonography. | 1/2 |
| What screening test should be done with A 45-year-old man with chronic HBV infection without cirrhosis?  A. Upper abdominal ultrasonography only  B. Serum alpha-fetoprotein only  C. Upper abdominal ultrasonography as the main screening tool with serum alphafetoprotein as an optional tool  D. Upper abdominal ultrasonography or serum alpha-fetoprotein  E. Both upper abdominal ultrasonography and serum alpha-fetoprotein  F. CT or MRI of upper abdomen  G. The patient has no indication for HCC surveillance. | The correct answer is E. Both upper abdominal ultrasonography and serum alpha-fetoprotein should be done as screening tests for a 45-year-old man with chronic HBV infection without cirrhosis.  Upper abdominal ultrasonography is a commonly used screening tool for detecting liver cancer, including HCC (hepatocellular carcinoma). It is a non-invasive and relatively inexpensive test that uses sound waves to create an image of the liver.  Serum alpha-fetoprotein (AFP) is a protein that is produced by the liver and certain other tissues. Elevated levels of AFP in the blood can be a sign of liver cancer.  Both of these tests can help to detect HCC in its early stages, when it is more treatable. It is important for individuals with chronic HBV infection to undergo regular HCC surveillance, as they are at increased risk for developing the disease. | E. Both upper abdominal ultrasonography and serum alpha-fetoprotein. It is recommended to perform both upper abdominal ultrasonography and serum alpha-fetoprotein as screening tests for hepatocellular carcinoma (HCC) in individuals with chronic HBV infection, particularly if they do not have cirrhosis. This is because HCC can occur in individuals with chronic HBV infection even in the absence of cirrhosis. Both of these tests have good sensitivity and specificity for the detection of HCC, and their combination can further improve the accuracy of HCC diagnosis. CT or MRI of the upper abdomen may also be used as screening tests, but they are generally more expensive and may not be necessary in all cases. | 3 |
| What screening test should be done with a 45-year-old woman with chronic HBV infection without cirrhosis?  A. Upper abdominal ultrasonography only  B. Serum alpha-fetoprotein only  C. Upper abdominal ultrasonography as the main screening tool with serum alphafetoprotein as an optional tool  D. Upper abdominal ultrasonography or serum alpha-fetoprotein  E. Both upper abdominal ultrasonography and serum alpha-fetoprotein  F. CT or MRI of upper abdomen  G. The patient has no indication for HCC surveillance. | The correct answer is E. Both upper abdominal ultrasonography and serum alpha-fetoprotein should be done for HCC surveillance in a 45-year-old woman with chronic HBV infection without cirrhosis. Upper abdominal ultrasonography and serum alpha-fetoprotein are both important tools for the screening and surveillance of hepatocellular carcinoma (HCC) in patients with chronic HBV infection. CT or MRI of the upper abdomen may also be used for HCC surveillance, but ultrasonography and alpha-fetoprotein are generally the primary screening tests. | It is generally recommended to screen individuals with chronic HBV infection for the development of hepatocellular carcinoma (HCC), including with upper abdominal ultrasonography and serum alpha-fetoprotein testing. Option E (both upper abdominal ultrasonography and serum alpha-fetoprotein) would be the most appropriate choice for HCC surveillance in a 45-year-old woman with chronic HBV infection. Option C (upper abdominal ultrasonography as the main screening tool with serum alphafetoprotein as an optional tool) is also a reasonable choice. Option A (upper abdominal ultrasonography only) and option B (serum alpha-fetoprotein only) would not be as thorough as using both tests, as they would only detect HCC in certain situations. Option F (CT or MRI of upper abdomen) would also be a reasonable choice, although it may be more costly and not always necessary. Option G (the patient has no indication for HCC surveillance) is incorrect, as individuals with chronic HBV infection are at increased risk for developing HCC and should undergo regular surveillance for the disease. | 4 |

### Supplemental Table 4B. Grading of the responses from the ChatGPT Language Model for the published HCC surveillance knowledge questionnaires

#### Article title: Knowledge and Perceptions of Hepatitis B and Hepatocellular Carcinoma Screening Guidelines Among Trainees: A Tale of Three Centers

| Questions | Responses 1 | Responses 2 | Reviewer Grade  1. Comprehensive  2. Correct but inadequate  3. Mixed with correct and incorrect/outdated data 4. Completely incorrect |
| --- | --- | --- | --- |
| Should a 25-year-old Haitian man with chronic hepatitis B, on treatment with entecavir be screened for Hepatocellular carcinoma (HCC)?  1. Yes  2. No | It is generally recommended that individuals with chronic hepatitis B, especially those who are on treatment with entecavir, should be screened for hepatocellular carcinoma (HCC), also known as liver cancer. This is because individuals with chronic hepatitis B are at an increased risk of developing HCC due to the ongoing damage to the liver caused by the virus. Therefore, it is important for individuals with chronic hepatitis B, including a 25-year-old Haitian man on treatment with entecavir, to undergo regular screening for HCC in order to detect any potential liver tumors at an early stage when they may be more treatable. | Yes. It is generally recommended that individuals with chronic hepatitis B, especially those on antiviral treatment, be screened for Hepatocellular carcinoma (HCC). This is because individuals with chronic hepatitis B are at an increased risk of developing HCC, which is a type of liver cancer. Early detection of HCC can improve treatment outcomes and potentially increase the chances of survival. Therefore, it is important for individuals with chronic hepatitis B, including those on antiviral treatment, to undergo regular screenings for HCC. | 1/2 |
| Should a 40-year-old Cuban woman who was recently diagnosed with hepatitis B be screened for Hepatocellular carcinoma (HCC)?  1. Yes  2. No | Yes, it is recommended that a person with hepatitis B be screened for Hepatocellular carcinoma (HCC), especially if they have a history of liver damage or cirrhosis caused by the virus. HCC is a type of liver cancer that can develop in people with chronic hepatitis B infection, and early detection is important for improving the chances of successful treatment. Therefore, it is important for the 40-year-old Cuban woman to undergo regular HCC screening as part of her overall care plan for managing her hepatitis B infection. | It is generally recommended that individuals with chronic hepatitis B infection be screened for hepatocellular carcinoma (HCC). This is because people with chronic hepatitis B infection have an increased risk of developing HCC, which is a type of liver cancer. The risk of developing HCC increases with the duration of infection and the severity of liver damage. Therefore, it is important for individuals with chronic hepatitis B infection to undergo regular screenings to detect HCC early, when it is most treatable.  In the case of a 40-year-old Cuban woman who has recently been diagnosed with hepatitis B, it would be appropriate for her to undergo screening for HCC. This could involve various tests such as blood tests, imaging tests (such as ultrasound or CT scan), or biopsy. It is important for her to discuss the appropriate screening schedule and tests with her healthcare provider. | 4 |
| Should a 25-year-old Vietnamese man with past history of hepatitis B be screened for Hepatocellular carcinoma (HCC)?  1. Yes  2. No | Yes, a 25-year-old Vietnamese man with a past history of hepatitis B should be screened for Hepatocellular Carcinoma (HCC). It is recommended that individuals with chronic hepatitis B infection undergo regular screening for HCC, as this type of cancer is a common complication of hepatitis B. The frequency and timing of screening depend on the individual's risk factors and liver health, but it is generally recommended to start screening at an earlier age for individuals with chronic hepatitis B. It is important for individuals with hepatitis B to receive proper medical care and follow a recommended screening schedule to detect and treat any potential complications, including HCC, as early as possible. | It is generally recommended that individuals with a past history of hepatitis B be screened for Hepatocellular Carcinoma (HCC), especially if they have advanced liver disease or cirrhosis. This is because individuals with hepatitis B have an increased risk of developing HCC. It is important for individuals at high risk for HCC to undergo regular screenings to detect the cancer at an early stage, when it may be more treatable. Therefore, the answer is 1. Yes, a 25-year-old Vietnamese man with a past history of hepatitis B should be screened for HCC. | 4 |
| A 35-year-old Chinese woman with platelets of 80,000, INR of 1.1, bilirubin of 1.4 mg/dL. On ultrasound, she was noted to have mild ascites and a spleen that measures 14 cm. Should she be screened for Hepatocellular carcinoma (HCC)?  1. Yes  2. No | It is not appropriate to use a single laboratory value or ultrasound finding to determine the need for screening for hepatocellular carcinoma (HCC). HCC is a type of liver cancer that is more common in people with chronic liver diseases such as cirrhosis, hepatitis B or C, and alcohol-related liver disease. Risk factors for HCC include long-term infection with hepatitis B or C, heavy alcohol use, cirrhosis, and a family history of HCC.  A comprehensive evaluation for HCC should consider all of the patient's risk factors and any clinical or laboratory abnormalities. This may include a thorough medical history and physical examination, imaging studies (such as ultrasound, CT, or MRI), and possibly liver biopsy.  It is important to note that HCC can be difficult to diagnose in its early stages because it often does not cause symptoms until it has reached an advanced stage. Therefore, it is important for individuals at high risk for HCC to undergo regular screenings to detect the disease at an early stage, when it is more treatable. | It is not possible to determine whether the patient should be screened for Hepatocellular carcinoma (HCC) based on the information provided. HCC is a type of liver cancer that can be associated with certain risk factors such as chronic liver disease, cirrhosis, and viral hepatitis. However, the presence of mild ascites and a spleen that measures 14 cm on ultrasound alone is not sufficient to determine whether the patient is at risk for HCC. It is important for the patient to discuss her individual risk factors and concerns with her healthcare provider. | 4 |
| Should a 40-year-old non-cirrhotic woman from Laos with chronic inactive hepatitis B be screened for Hepatocellular carcinoma (HCC)?  1. Yes  2. No | It is generally recommended that individuals with chronic hepatitis B, including those who are 40 years old and not cirrhotic, be screened for hepatocellular carcinoma (HCC). This is because individuals with chronic hepatitis B have an increased risk of developing HCC compared to the general population. Regular monitoring and early detection of HCC can improve the chances of successful treatment. It is important for individuals with chronic hepatitis B to discuss the appropriate screening and monitoring recommendations with their healthcare provider. | It is generally recommended that individuals with chronic hepatitis B, regardless of their age or cirrhosis status, be screened for hepatocellular carcinoma (HCC). This is because individuals with chronic hepatitis B are at an increased risk of developing HCC, and early detection and treatment of HCC can improve outcomes. Therefore, it is appropriate for a 40-year-old non-cirrhotic woman from Laos with chronic inactive hepatitis B to be screened for HCC. | 4 |
| Should a 40 year old cirrhotic woman from Thailand with chronic inactive hepatitis B be screened for Hepatocellular carcinoma (HCC)?  1. Yes  2. No | It is generally recommended that individuals with chronic inactive hepatitis B be screened for hepatocellular carcinoma (HCC). This is especially important for those who have advanced liver disease, such as cirrhosis, as they are at higher risk for developing HCC. The frequency and method of screening should be determined by a healthcare provider, taking into account the individual's specific circumstances and risk factors. It is important for individuals with chronic inactive hepatitis B to work closely with their healthcare providers to manage their condition and monitor for any potential complications. | Yes, a 40-year-old woman with cirrhosis and chronic inactive hepatitis B should be screened for Hepatocellular carcinoma (HCC). It is recommended that individuals with cirrhosis, including those with hepatitis B, undergo regular monitoring for the development of HCC. This typically involves imaging tests such as ultrasound, CT scan, or MRI every 6 to 12 months. Early detection and treatment of HCC can significantly improve the chances of a successful outcome. It is important for this individual to discuss with their healthcare provider about the specific screening recommendations and frequency based on their individual circumstances. | 1^st^ response: 1  2^nd^ response: 3 |
| Should A 40-year-old Nigerian woman with chronic hepatitis B with normal liver function tests be screened for Hepatocellular carcinoma (HCC)?  1. Yes  2. No | Yes, it is recommended that individuals with chronic hepatitis B undergo screening for hepatocellular carcinoma (HCC). This is especially important if the individual has a high viral load or advanced liver fibrosis. Even if the individual's liver function tests are normal, it is still important to monitor for the development of HCC, as it can develop even in individuals with normal liver function tests. It is recommended that individuals with chronic hepatitis B discuss their specific screening recommendations with their healthcare provider. | It is generally recommended that individuals with chronic hepatitis B, especially those with normal liver function tests, be screened for hepatocellular carcinoma (HCC). Early detection of HCC can improve the chances of successful treatment, so it is important for individuals with chronic hepatitis B to undergo regular screenings. It is also important for individuals with chronic hepatitis B to follow a healthy lifestyle and take their medications as prescribed to help prevent the development of HCC. | 4 |
